# The isolated effect of age on the risk of COVID-19 severe outcomes: a systematic review with meta-analysis

**DOI:** 10.1101/2021.05.27.21257909

**Authors:** Karla Romero Starke, David Reissig, Gabriela Petereit-Haack, Stefanie Schmauder, Albert Nienhaus, Andreas Seidler

## Abstract

**Introduction:** Increased age has been reported to be a factor for COVID-19 severe outcomes. However, many studies do not consider the age-dependency of comorbidities, which influence the course of disease. Protection strategies often target individuals after a certain age, which may not necessarily be evidence-based. The aim of this review was to quantify the isolated effect of age on hospitalization, admission to ICU, mechanical ventilation, and death.

**Methods:** This review was based on an umbrella review, in which Pubmed, Embase, and pre-print databases were searched on December 10, 2020 for relevant reviews on COVID-19 disease severity. Two independent reviewers evaluated the primary studies using predefined inclusion and exclusion criteria. The results were extracted, and each study was assessed for risk of bias. The isolated effect of age was estimated by meta-analysis, and the quality of evidence was assessed using GRADE.

**Results:** Seventy studies met our inclusion criteria (case mortality n=14, in-hospital mortality n=44, hospitalization n=16, admission to ICU n=12, mechanical ventilation n=7). The risk of in-hospital and case mortality increased per age year by 5.7% and 7.4%, respectively (Effect Size (ES) in-hospital mortality=1.057, 95% CI:1.038-1.054; ES case mortality= 1.074, 95% CI:1.061-1.087), while the risk of hospitalization increased by 3.4% per age year (ES=1.034, 95% CI:1.021-1.048). No increased risk was observed for ICU admission and intubation by age year. There was no evidence of a specific age threshold at which the risk accelerates considerably. The confidence of evidence was high for mortality and hospitalization.

**Conclusions:** Our results show a best-possible quantification of the increase in COVID-19 disease severity due to age. Rather than implementing age thresholds, prevention programs should consider the continuous increase in risk. There is a need for continuous, high-quality research and “living” reviews to evaluate the evidence throughout the pandemic, as results may change due to varying circumstances.

**What is already known?:**

- Increasing age and comorbidities are risk factors for COVID-19 severe outcomes, such as hospitalization and mortality. However, comorbidities such as diabetes, cardiovascular disease, chronic pulmonary diseases increase with age, and the isolated effect of age on COVID-19 disease severity is not known.

**What are the new findings?:**

- The risk of COVID-19 disease severity due to the isolated effect of age increases by age year and no specific age threshold was observed.
- A best possible quantification of the increase in risk of COVID-19 severe outcomes due to age has been done.

**What do the new findings imply?:**

- Any workplace restrictions targeting a particular older age group are rather arbitrary, and may contribute to ageism in the society.
- If scores are to be built to assess an individual’s risk for COVID-19 severe outcomes in workplace settings, these should be based on per-age increases.

## 1. Introduction

Since identified in Wuhan, China in December 2019 (1), COVID-19 has caused more than 3.4 million deaths worldwide (2). Early on, the increase COVID-19 adverse outcomes in the older population was observed (3-6). Chronic diseases, which increase with age, are also established risk factors for COVID-19 disease severity (7, 8). However, age and age-related risk factors, are often not de-coupled in studies (9). Since mortality rates are often presented by age groups (10), prevention strategies targeting older age groups in the workplace were started. After the first wave, the Federal State of Lower Saxony in Germany encouraged teachers over the age of 60 years to work from home. Recently, an IKKA score was developed in Germany for workers in order to establish, among other things, age-based criteria that could decide on the admissibility of work attendance (11). The risk score was partly-based on the age group of the individual (<50 years: score=0; 50-59 years: score=4; ≥60 years: score=10). There have been discussions concluding in a consensus over the need to improve the risk score based on current evidence regarding the isolated effect of age on COVID-19 disease severity (12). In general, we should be careful to classify 60 years of age as risk persons (or any other age group starting at a particular “arbitrary” age), as this can considerably reduce the chances of older unemployed people finding a job or make them “targets” for layoffs.

In mid-2020, we published a rapid review investigating the isolated effect of age on COVID-19 disease severity - that is, the direct effect of age after accounting for important age-related risk factors such as diabetes, cardiovascular disease, and chronic pulmonary disease. The result was that the effect of age was rather small (13). This review was based mainly on Chinese studies published early in the pandemic. As more studies have been published worldwide, a different picture might emerge, and it is necessary to examine the newly acquired evidence on the isolated effect of age on COVID-19 disease severity.

We aim to gather information on published studies to answer the following:

1. What is the isolated effect of age on COVID-19 disease severity (hospitalization, admission to the intensive care unit (ICU), mechanical ventilation or death due to COVID-19), after adjusting for important age-related risk factors?
2. Is there evidence of an age threshold in which the risk of COVID-19 increases rapidly?

## 2. Methods

### Search, selection, and data extraction

This systematic review was based on an umbrella review on pre-existing health conditions and severe COVID-19 outcomes published elsewhere (14). To summarize, on December 11, 2020 a systematic search for systematic reviews was done in PubMed and Embase, with hand searches on preprint servers (search string in supplement). The included systematic reviews investigated the association between at least one chronic health condition and a severe COVID-19 outcome, such as hospitalization, ICU admission, intubation, or death. After screening the titles and abstracts, followed by full text screening, 120 systematic reviews were included (Figure S1). Then, primary studies included in these reviews were screened. They were included in the analysis if they reported at least one quantitative measure of association in persons with pre-existing health conditions and at least one age-adjusted estimate. 160 primary studies were included in the umbrella review (Figure S2) (14).

To find adequate primary studies for the purpose of our research question regarding age and COVID-19 disease severity, we excluded studies investigating special populations, such as cancer patients or persons with diabetes. Studies were included if the age effect had been adjusted for at least three of the following comorbidities: diabetes mellitus, cardiovascular disease, cancer/immunodeficiency, chronic kidney disease, chronic liver disease, and chronic pulmonary disease (see Table 1 for eligibility criteria). Two independent scientists screened the full texts of the studies. In case of disagreement, a consensus decision was sought between both scientists. If no agreement was achieved, the decision was made by a third reviewer.

**Table 1.** Eligibility criteria

|  | Inclusion criteria | Exclusion criteria |
| --- | --- | --- |
| Population | General population infected with COVID-19 (both sexes, all ages) | Special populations such as studying only cancer patients or persons with diabetes |
| Exposure | Age, in years | All other exposures which do not include age |
| Comparator/control | Age reference, in years |  |
| Outcomes | Due to infection with COVID-19: risk of hospitalization, admission to ICU, intubation, and death.<br>Risks measured as hazard ratios, risk ratios, odds ratios | Other outcomes, including outcomes of composite disease severity. |
| Study design | cross-sectional, case-control, and cohort studies | RCTs, qualitative studies, ecological studies, case reports, experiments, congress abstracts, posters |

Excluded are studies only reporting univariate (unadjusted) effect values of age. Excluded are studies in which the age effect is not adjusted at least for three comorbidities listed here: diabetes mellitus, cardiovascular disease , cancer or immunodeficiency, chronic kidney disease, chronic liver disease, and chronic pulmonary disease .

Data extraction was done by one reviewer and checked by another one. In case of missing information, study authors were contacted. The data extraction form included information on the first author, publication year, country of origin, study population, age and sex characteristics, outcome assessment, confounders, analysis methods, and results. A protocol of the review was registered a priori with the PROSPERO database of systematic reviews (https://www.crd.york.ac.uk/prospero/display_record.php?RecordID=220614).

### Risk of Bias Assessment

We evaluated the overall risk of bias for each study as “low” or “high”, and followed the procedure for assessment based on Ijaz et al. 2013 (15), with modifications (13, 16). We also considered the criteria described by SIGN (Scottish Intercollegiate Guidelines, 2012) (17) and CASP (Critical Appraisal Skills Programme) (18). We assessed eight domain: 1) recruitment procedure and follow-up 2) exposure definition and measurement, 3) outcome definition and measurement, 4) inclusion of important age-dependent risk factors, 5) analysis method, 6) chronology, 7) funding, and 8) conflict of interest. Each domain was characterized as having a low, high, or unclear risk of bias (see supplementary material for a detailed description). Domains 1-5 and 6-8 were major and minor domains, respectively. A study was evaluated as having an overall low risk of bias if all major domains had a low risk of bias.

### Data synthesis

We evaluated studies using age categories and calculated the median of each of the age categories. If lower and upper categories were open ended, we used reasonable values based on the youngest and oldest participants included. For the younger open-ended age groups, we chose a lower boundary of 18 years if the population encompassed adults only. The median value of 18 years and the upper boundary of the lower open-ended age category was estimated. For the older open-ended age groups an upper boundary of 95 years of age was chosen and a median value was similarly calculated. However, these values were also modified in a sensitivity analysis. If studies only had risks of binary categories of age, such as >60 years vs ≤60 years, they were excluded from the meta-analysis. A log-linear model and a log-cubic model was constructed weighted for variance, and the Akaike Information Criteria (AIC) for each model was evaluated. The log of the relative risk (RR) was then plotted against the age mid-points to assess linearity for the specific outcome studied, first for each individual study, and then for all studies using categorized age. We wanted to assess whether (a) there is an age threshold present at which the ln(RR) increases and (b) whether a log-linear relationship for the categorical studies would be a reasonable assumption. If a log-linear association could be assumed, a risk effect per year was obtained by using the Generalized Least Squares for trend estimation of summarized dose-response data (*glst*) (19). If the number of cases per age category was unknown, the risk estimate was obtained by variance-weighted least squares (*vwls*). If the log-linear association could be assumed, then the studies using categorical values were pooled with the ones using continuous values through a random-effects meta-analysis using the *metan* package (20). A similar procedure has been done before (15, 21). If two studies investigated the same population, the study with the highest quality was chosen. Odds ratios (OR) were converted to RRs using the formula proposed by Zhang and Yu 1998 (22) when the prevalence in the reference group was greater than 10% to avoid an overestimation of the RRs.

We compared the pooled effects of studies using continuous values for age and those using categorical values. Heterogeneity was assessed by I^2^ values, and publication bias was explored using Funnel plots and Egger’s test (*metabias*) if there were at least ten studies. Stata Version 14.2 (StatCorp, College Station, TX, USA) was used.

### 2.4 Assessment of the quality of the total body of evidence

We used the Grading of Recommendations Assessment, Development, and Evaluation (GRADE) approach for grading the quality of the total body of evidence (23), following the example of Hulshof et al. 2019 (24), with modifications (25). The quality of evidence was either high, moderate, or low. Since only observational studies were included, the starting level was set to “moderate”. The quality was downgraded based on four factors: study limitations, indirectness, inconsistency, imprecision, and publication bias. If study findings had large effect sizes, if there were dose-response relationships, or if the presence of residual confounding was suspected (increasing the confidence in the association), the quality of evidence was upgraded. For the effect size evaluation, we considered the difference in risk due to age between a person entering adulthood (18 years) and the worldwide life expectancy (72 years) (26), a difference of 54 years.

## Results

After reviewing the 160 included primary studies from the umbrella review (14), we included 70 studies (27-96) (Figure 1). Table S1 lists the excluded studies with reasons.

**Figure 1.**
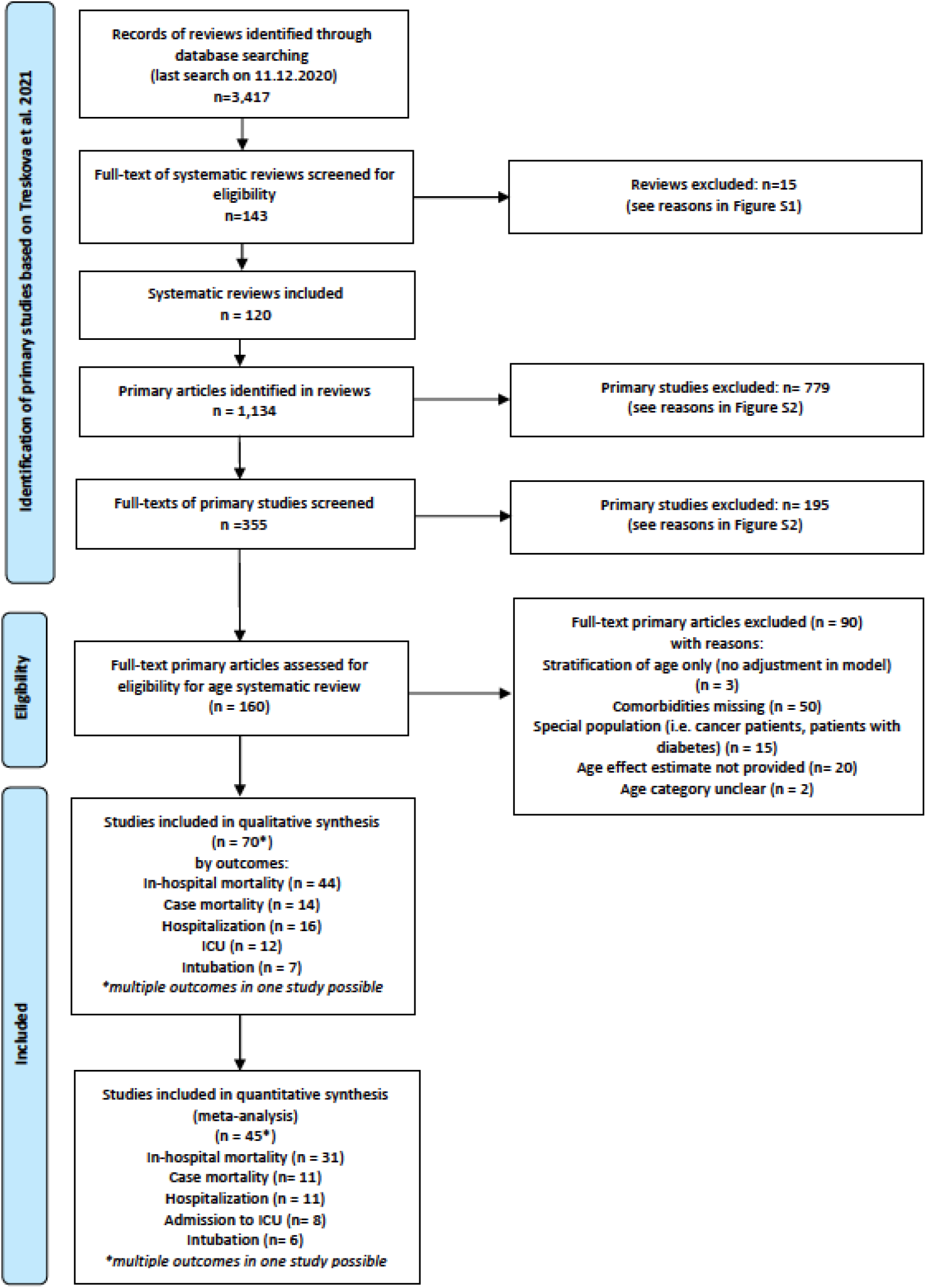
Selection process for primary studies from original Umbrella Review (Treskova et al. 2020)

Except for Gu et al. 2020 (51), a case control study, all studies were either retrospective or prospective cohort studies. Most originated from USA (34), while 14 studies came from Europe, 8 from Latin America (Mexico and Brazil), 10 from China, two from South Korea, and one each from South Africa and Israel. Details on study characteristics are in Table S2.

### In-hospital mortality

Forty-four studies investigating the risk of in-hospital mortality, meaning the risk of mortality in hospitalized patients, were included. Most studies were set in Western countries, such as USA (n=21), Europe (n=13), and Latin America (n=2). Six studies were set in China, one in South Africa, and one in South Korea. Nineteen studies reported the risk of in-hospital mortality per age year, while the others used age categories to report the effect of age on risk. The results can be seen in Table S3. From the 44 studies, eight studies (28, 30, 34, 48, 64, 72, 75, 77) were assessed as having a low risk of bias. The majority had a high risk of bias because not all six age-dependent comorbidities were included in the model, while some used rough age categories in their models (greater than 10 years or a binary age category such as >60 years vs ≤60 years) (Table S4).

Thirty-one studies were included in the meta-analysis. Six studies (27, 37, 58, 65, 67, 83) were excluded because of the use of a binary age category, while one was excluded because the risk was reported by an age z-score, making it hard to compare to the other studies (81). Four studies were excluded from the main meta-analysis because they investigated mortality only on critically-ill hospitalized patients or patients with COVID-19 cytokine storm (43, 50, 53, 72). Two studies (36, 56) investigated the same study population, but Carter and Collins et al. 2020 (36) was chosen because this study had adjusted for more age-related risk factors. Likewise, Shi et al. 2020 (84) and Zhao et al. 2020 (95) used the same population. The former study was used because it reported age as a continuous factor, rather than a very rough age category. For the studies using age as categorical variables, the age and log of RR were modeled as linear and cubic relationships and plotted (Figure S3). Although the weighted cubic model had a lower AIC value (−71.8) than the weighted linear model (−62.5), the cubic model showed no evidence of a rapid increase in the slope at a specific age. At around age 70, there was a downturn in the slope which did not follow the points, indicating a possible better fit with a fractional or higher-level polynomial relationship. We assumed a log-linear relationship for simplicity, as at least for the ranges 20 to 70 years there seemed to be no conflict with that assumption. A per-year effect was calculated for studies using age categories and they were included in the meta-analysis.

The pooled RR for the 31 studies was 1.046 (95% CI: 1.039-1.054) per age year (Table 2). The funnel plot was asymmetric (Figure S4; Egger’s test p<0.0001), but the funnel plot using age as a continuous variable differed from the one using age derived from categories (Figure S5a-b). Upon closer look, the pooled RR of studies using age as a continuous variable (16 studies, 52%) was higher than the pooled RR of studies dividing age into categories (RR_age continuous_=1.057; 95% CI 1.047-1.067; RR_age categories_=1.037; 95% CI 1.028-1.046). Since the calculated midpoints from the age categories might be inaccurate, we hypothesized that the RR of studies using a higher number of age categories would be closer to the RR_age continuous_. Indeed, studies having more than four age categories had a pooled RR of 1.039 (95% CI 1.026-1.053), whereas studies with four or less age categories had a pooled RR of 1.030 (95% CI 1.020-1.041). We divided studies by age category width (5 vs. 10 vs. 10+ years intervals) and found that the effect estimate was lowest with studies using the widest age categories (10+ years RR=1.030; 95% CI 1.021-1.039), followed by studies using 10-year categories (RR=1.046; 95% CI 1.040-1.051). Only one study used a 5-year age category, having the highest estimate (RR=1.060; 95%CI 1.051-1.070). Because of probable bias using different age categories, further analyses for in-hospital mortality focused on studies using age as a continuous variable (Table 2). The pooled RR did not differ by study quality. Studies from USA, Europe, and South Korea had similar pooled RRs, whereas as the pooled RR for the Chinese studies was lower (RR=1.041; 95% CI 0.999-1.085). The unadjusted and adjusted RRs on studies reporting both values were similar (RR_unadjusted_=1.060, 95% CI: 1.034-1.086; RR_adjusted_=1.059, 95% CI: 1.037-1.081).

**Table 2.**
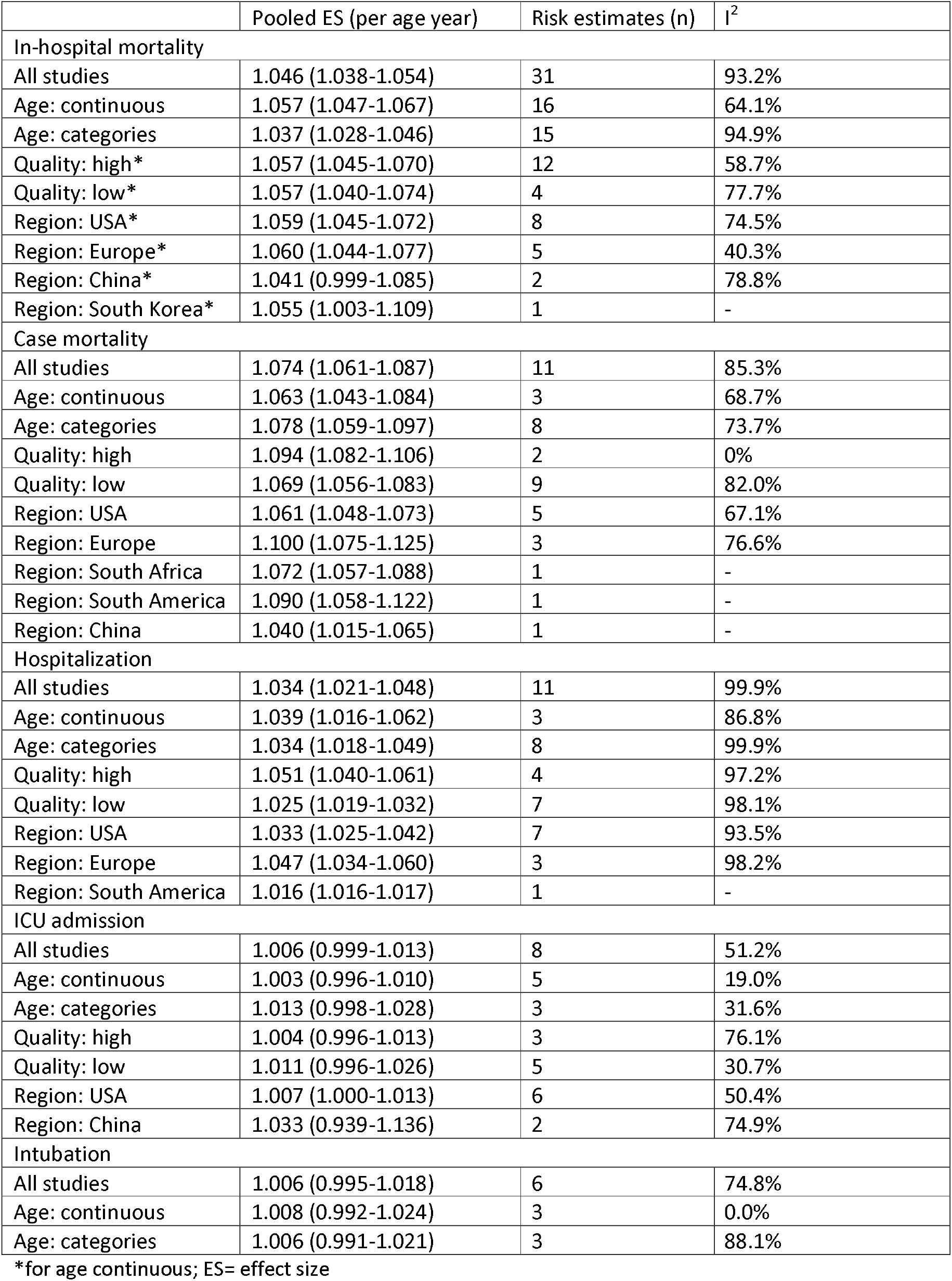
Results of pooled risks from meta-analyses

|  | Pooled ES (per age year) | Risk estimates (n) | I<sup>2</sup> |
| --- | --- | --- | --- |
| In-hospital mortality |  |  |  |
| All studies | 1.046 (1.038-1.054) | 31 | 93.2% |
| Age: continuous | 1.057 (1.047-1.067) | 16 | 64.1% |
| Age: categories | 1.037 (1.028-1.046) | 15 | 94.9% |
| Quality: high* | 1.057 (1.045-1.070) | 12 | 58.7% |
| Quality: low* | 1.057 (1.040-1.074) | 4 | 77.7% |
| Region: USA* | 1.059 (1.045-1.072) | 8 | 74.5% |
| Region: Europe* | 1.060 (1.044-1.077) | 5 | 40.3% |
| Region: China* | 1.041 (0.999-1.085) | 2 | 78.8% |
| Region: South Korea* | 1.055 (1.003-1.109) | 1 | - |
| Case mortality |  |  |  |
| All studies | 1.074 (1.061-1.087) | 11 | 85.3% |
| Age: continuous | 1.063 (1.043-1.084) | 3 | 68.7% |
| Age: categories | 1.078 (1.059-1.097) | 8 | 73.7% |
| Quality: high | 1.094 (1.082-1.106) | 2 | 0% |
| Quality: low | 1.069 (1.056-1.083) | 9 | 82.0% |
| Region: USA | 1.061 (1.048-1.073) | 5 | 67.1% |
| Region: Europe | 1.100 (1.075-1.125) | 3 | 76.6% |
| Region: South Africa | 1.072 (1.057-1.088) | 1 | - |
| Region: South America | 1.090 (1.058-1.122) | 1 | - |
| Region: China | 1.040 (1.015-1.065) | 1 | - |
| Hospitalization |  |  |  |
| All studies | 1.034 (1.021-1.048) | 11 | 99.9% |
| Age: continuous | 1.039 (1.016-1.062) | 3 | 86.8% |
| Age: categories | 1.034 (1.018-1.049) | 8 | 99.9% |
| Quality: high | 1.051 (1.040-1.061) | 4 | 97.2% |
| Quality: low | 1.025 (1.019-1.032) | 7 | 98.1% |
| Region: USA | 1.033 (1.025-1.042) | 7 | 93.5% |
| Region: Europe | 1.047 (1.034-1.060) | 3 | 98.2% |
| Region: South America | 1.016 (1.016-1.017) | 1 | - |
| ICU admission |  |  |  |
| All studies | 1.006 (0.999-1.013) | 8 | 51.2% |
| Age: continuous | 1.003 (0.996-1.010) | 5 | 19.0% |
| Age: categories | 1.013 (0.998-1.028) | 3 | 31.6% |
| Quality: high | 1.004 (0.996-1.013) | 3 | 76.1% |
| Quality: low | 1.011 (0.996-1.026) | 5 | 30.7% |
| Region: USA | 1.007 (1.000-1.013) | 6 | 50.4% |
| Region: China | 1.033 (0.939-1.136) | 2 | 74.9% |
| Intubation |  |  |  |
| All studies | 1.006 (0.995-1.018) | 6 | 74.8% |
| Age: continuous | 1.008 (0.992-1.024) | 3 | 0.0% |
| Age: categories | 1.006 (0.991-1.021) | 3 | 88.1% |

\*for age continuous; ES= effect size

### Case Mortality

Fourteen studies investigating case mortality were included, in which mortality was ascertained in individuals who had tested positive for COVID-19. Five studies were from USA (52, 54, 61, 90, 92), three studies originated from Europe (34, 78, 79), three from Latin America (35, 86, 87), and one each from China (51), South Africa (33), and South Korea (66). The results can be seen in Table S5. Three studies (21%) were rated as having a low risk of bias. Most of the studies had a high risk of bias because they did not adjust for all age-dependent risk factors, the age categories were quite wide, and the recruitment procedure indicated a possible selection bias (i.e. such as recruitment of only symptomatic people or people who presented for care at hospitals without being hospitalized, or if ICD codes used for recruitment were not necessarily related to COVID-19) (Table S6).

Eleven studies were included in the meta-analysis, of which the majority used age categories (8 of 11). One study was excluded due to the use of binary age categories (87). One study (54) reported the age year RR within age categories, and therefore it was not possible to compare to the other studies. Two studies used the same study population (35, 86), but Solis et al. 2020 (86) was chosen because the study had adjusted for more age-related factors and it had used a higher number of age categories.

The cubic model showed a relatively flat relationship between log(RR) and age from ages 40 to 85 years (Figure S6). Because both models had similar AIC values (linear: −157.4; cubic: −159.3), we assumed a linear model.

The meta-analysis resulted in a pooled RR of 1.074 (95% CI 1.061-1.087) per age year (Table 4). The respective funnel plot showed asymmetry (Figure S7), although Egger’s test was not significant (p=0.320). The pooled RR of studies using continuous (RR=1.063; 95% CI 1.043-1.084) and categorical (RR=1.078; 1.059-1.097) age variables differed. However, since the 95% CI values overlapped, there was no significant difference observed. We divided studies by age category width (5 vs. 10 vs. 10+-year intervals) and again found that the effect estimate was lowest with studies using the widest age categories (10+ years RR=1.042; 95% CI 1.028-1.056; 10 years RR=1.080; 95% CI 1.057-1.103) and highest for studies using the most narrow age of five years (RR=1.094; 95% CI 1.082-1.107).

The high-quality studies had a higher RR per age than low quality studies (RR_high quality_=1.094, 95% CI: 1.082-1.106; RR_low quality_=1.069; 95% CI: 1.056-1.083). European studies (n=3) had the highest pooled RR (RR= 1.100; 95% CI 1.075-1.125), followed by USA studies (RR= 1.061; 95% CI 1.075-1.125). Other regions had only one study, so a reliable comparison was not possible. The unadjusted and adjusted RRs on studies reporting both values were similar (RR_unadjusted_=1.079; 95% CI: 1.062-1.097; RR_adjusted_=1.072; 95% CI: 1.054-1.091).

### Hospitalization

A summary of the results of studies investigating risk of hospitalization by age can be seen in Table S7. The study population typically used were individuals who were positive for COVID-19, some of which were hospitalized due to complications related to the infection. Sixteen studies were included, of which nine originated in the USA (29, 38, 47, 52, 63, 76, 77, 91, 96), three in Europe (34, 78, 79), three in Latin America (35, 49, 85), and one in Israel (70). Four (25%) of the studies had a low risk of bias, and the reasons for high risk of bias were mainly the lack of adjustment for all pre-defined age-related risk factors and the use of binary or rough age categories (Table S8).

Of the sixteen studies, eleven were included in the meta-analysis. Ebinger et al. 2020 (47) was excluded because the outcome was complex with reference categories that could not be compared to other studies (hospitalized/non-ICU vs. ICU/non-intubated vs ICU/intubated). Carrillo-Vega et al. 2020 (35) and Giannouchos et al. 2020 (49) used the same study population, but the latter was used because the study had more age categories and adjusted for a higher number of age-related risk factors. Furthermore, two studies (70, 85) used binary age categories and one did not provide confidence intervals (63). Most studies (8 of 11) used age categories.

The results of the weighted linear and cubic models of studies using age categories are shown in Figure S8. Although the AIC values for the linear model were higher than for the cubic model (−57.2 vs −21.7, we did choose the linear model because of a lack of threshold where the rate increases with age and because the age range modeled appeared as a linear relationship.

The pooled RR for all studies was 1.034 (95% CI: 1.021-1.048). The funnel plot looked asymmetric (Figure S9), but Egger’s test was statistically not-significant (p=0.439). The RR of studies using continuous and categorical age values were very similar (Table 4), noting that most studies (63%) using age categories utilized more than four age categories. Again, using a finer age width increased the RR for studies using age categories (10+ years RR=1.024 95% CI: 1.018-1.030; 10 years RR=1.040 95%CI: 1.037-1.043; 5 years RR=1.056 95% CI:1.055.1.057). The pooled effect for high-quality studies was higher than for low quality studies (RR_high quality_ =1.051, 95% CI: 1.040-1.061; RR_low quality_=1.025; 95% CI: 1.019-1.032). Moreover, the European studies had a higher pooled risk (RR=1.047; 95% CI: 1.034-1.060) than the USA studies (RR=1.033, 95% CI: 1.025-1.042), but the difference was not statistically significant. The unadjusted RR values were slightly higher than the adjusted values (RR_unadjusted_=1.045, 95% CI: 1.026-1.065; RR_adjusted_=1.031, 95% CI: 1.009-1.053).

### Admission to ICU

Twelve studies investigated the effect of age on admission to ICU. Typically, the studies used hospitalized individuals as the study population and followed them up (retrospectively or prospectively). Eight studies originated in the USA (28, 55, 61, 64, 77, 82, 88, 96) and four were from China (32, 46, 51, 89). A summary of the results can be seen in Table S9. Four (33%) of the studies were of high quality (low risk of bias). Similar to the other outcomes, the reasons for a high risk of bias were mainly due to the confounding domain, and to a lesser extent, to the exposure domain (rough age categories) (Table S10).

Eight studies were included in the meta-analysis. Du et al. 2020 (46) was not included. Due to the nature of its complex outcome, it could not be compared to other studies. Two studies evaluated the risk of ICU on individuals who had tested positive to COVID-19 (51, 96), and one study used a binary age category (88).

The pooled RR for the studies was 1.006 (95% CI: 0.999-1.013). Five studies (63%) used age as a continuous variable, and these studies had only a slightly lower RR (1.003; 95% CI: 0.996-1.010) compared to the ones using age as a categorical value (RR=1.013; 95% CI: 0.998-1.028), but still the effect was statistically not-significant for both. The high quality studies had a slightly lower RR than the low quality studies (RR_high quality_=1.004, 95% CI: 0.996-1.013; RR_low quality_=1.011; 95% CI: 0.996-1.026), but again both estimates were not statistically-significant (Table 4).

### Mechanical ventilation (intubation)

Seven studies investigated the effect of age on the risk of intubation, using individuals hospitalized due to COVID-19. All studies were set in the USA (42, 55, 74, 82, 88, 91), and a summary of the results can be seen in Table S11. No studies had a low risk of bias (high quality), either due to the “confounding” or “exposure assessment” domain (Table S12). Half of the studies used age categories, and the effect estimate was similar for studies using a continuous and a categorical age variable. One study (88) was excluded from the meta-analysis because it used a binary category for age. The pooled RR showed a very weak statistically not-significant increase in risk per age year (RR=1.006; 95% CI 0.995-1.018) (Table 4).

### Quality of evidence

Since only observational studies were included, the initial level of evidence was set at “moderate”. After decreasing a level for unclear publication bias, and increasing two levels for dose-response effect and two levels for a large effect estimate, the overall certainty of evidence was high for both risk of in-hospital mortality and case mortality. For hospitalization, after downgrading for high inconsistency and unclear publication bias, and upgrading for dose response, effect estimate, and effect size, the overall certainty of evidence was high (Table 5).

**Table 5.** Assessment of evidence for the risk of studied outcomes based on Grades of Recommendations, Assessment, Development, and Evaluation framework (GRADE).

| Risk | Quality of study limitations: ↓ | Indirectness of evidence: ↓ | Inconsistency: ↓ | Imprecision, range confidence interval effect size >2.0: ↓ | Publication bias, yes or unclear: ↓ | Effect estimate >2.0: ↑ >5.0: ↑↑ | Dose-response effect: ↑ | Residual confounding: ↑ | Overall certainty (high, moderate, low) |
| --- | --- | --- | --- | --- | --- | --- | --- | --- | --- |
| In-hospital mortality | no (-)<sup>1</sup> | no (-) | no (-)<sup>2</sup> | no (-) | unclear ↓ | yes ↑↑<sup>3</sup> | yes ↑ | no (-) | high |
| Case mortality | no (-)<sup>1</sup> | no (-) | no (-)<sup>4</sup> | no (-) | unclear ↓ | yes ↑↑<sup>5</sup> | yes ↑ | no (-) | high |
| Hospitalization | no (-)<sup>1</sup> | no (-) | yes ↓<sup>6</sup> | no (-) | unclear ↓ | yes ↑↑<sup>7</sup> | yes ↑ | no (-) | high |

<sup>1</sup> High quality studies also reported a significant association between age and risk of outcome. <sup>2</sup> Moderate heterogeneity (I<sup>2</sup>=64.1%) was observed on studies using age as a continuous variable and on high-quality studies (I<sup>2</sup>= 58.7%) <sup>3</sup> Although per age RR is 1.057, when comparing different adult groups can have an effect size of greater than 5.0 (i.e., a 54-year difference, 18 years vs 72 years results in a RR of 20.0) <sup>4</sup> Moderate heterogeneity (I<sup>2</sup>=68.7%) was observed on studies using age as a continuous variable and on high-quality studies heterogeneity was very low (I<sup>2</sup>= 0%) <sup>5</sup> Although per age RR is 1.074, when comparing different adult groups can have an effect size of greater than 5.0 (i.e., a 54-year difference, 18 years vs 72 years results in a RR of 47.2) <sup>6</sup> High heterogeneity observed (I<sup>2</sup> > 85%) <sup>7</sup> Although per age RR is 1.034, when comparing different adult groups can have an effect size of greater than 5.0 (i.e., a 54-year difference, 18 years vs 72 years results in a RR of 6.08)

There was no statistically-significant effect observed for admission to ICU and intubation. Hence, these outcomes were not included in the assessment.

## Discussion

An increased age-related risk of COVID-19 in-hospital mortality, case mortality, and hospitalization of 5.7%, 7.4%, and 3.4% per age year, respectively, was observed, with a high quality of evidence. Further, there was no evidence of an age threshold at which the risk of disease severity increased, with the age effect appearing to be linear. The association between age and both ICU admission and mechanical ventilation was weak and not statistically-significant. Although effects varied slightly by region, there were no obvious trends observed.

Our results for case mortality are in agreement with a review which found a log-linear association between age and infection fatality rate, but which did not include comorbidities in their analysis (9). Strengths and limitations

To our knowledge, since our rapid review published (13), this is the first systematic review primarily focused on estimating the isolated effect of age on the risk of COVID-19 disease severity, including case and in-hospital mortality, hospitalization, ICU admission, and intubation. It encompasses 70 primary studies with data from more than 400,000 participants and includes only studies adjusting for important age-related factors associated with COVID-19 disease severity (14). Adjusting for comorbidities allows to explicitly “factor out” the mediating effect of comorbidities in order to get the isolated effect of age, or the risk presented by a person of a given age who has no preexisting conditions.

The results in this review should be considered in light of its limitations. For studies reporting age as a categorical value, since the age categories used in the studies could not be compared because of their heterogeneity, the median of the age category represented the age for the reported effect size. This procedure could have led to inaccuracies of the effect size. This was evident in that effect sizes increased when using finer age categories, indicating that the use of wide age categories most likely leads to an underestimation of the age effect. We reported the pooled effect sizes for studies using age as a continuous variable for the outcome in-hospital mortality. For the other outcomes we reported the pooled effect size for studies using categorical and continuous age variables together since their effect was similar.

The funnel plots were asymmetrical, which was especially obvious for in-hospital mortality, where half of the studies used age as a categorical variable. However, funnel plot asymmetry has other possible causes besides publication bias (97). In this case, we believe that the cause is due to the heterogeneity of the exposure-namely when using studies with age as categorical variables. This was indeed shown when plotting funnel plots separately for studies using continuous and categorical age variables. The studies using continuous variables did not show obvious asymmetry in the funnel plots, while the studies using categorical age variables did show asymmetry. However, they had a narrow range of standard errors, which is also a contraindication for using funnel plots (97, 98). Even though we downgraded for publication bias in our GRADE evaluation, the overall assessment of evidence was still rated as high.

The adjusted effect estimates were only slightly lower than the unadjusted estimates, or at least the difference was not as great as observed in our previous rapid review. An explanation could be a potential interaction effect between age and certain chronic diseases, or a possible selection effect. This was observed in the related umbrella review by Treskova et al. 2021 (14) and the review by Mesas et al. 2021 (99) where younger individuals who had heart failure and kidney disease had a higher risk of in-hospital mortality compared to the risk observed in older individuals. This effect was also seen for hypertension (99). It may be that older patients are more quickly admitted to the hospital with less severe concomitant diseases than younger patients, which may be reflected as an increased in-hospital mortality for younger patients, essentially due to a selection effect in the hospital admission.

There was no increased risk by age year for being admitted to the ICU or for mechanical ventilation, but this result is hard to reconcile, given the strength of association between age and mortality. The included studies mostly occurred during the first and second waves, when ICU capacity was limited. How this lack of association is due to the possibility of triage in some scenarios is unknown, but worth considering.

Lastly most of the primary studies included were done in the first or second wave of the pandemic, and new mutations which may affect the association between age and disease severity were not considered.

### Methodological quality of the included studies

Although high-quality studies were present for every outcome except mechanical ventilation, low-quality studies dominated. High-quality studies had the same effect estimate as low-quality studies for in-hospital mortality, but for case mortality and hospitalization, low-quality studies underestimated the isolated effect of age. The main reason for a low risk of bias was the lack of adjustment for all age-related risk factors in the domain “confounding”. Moreover, considering the “recruitment” domain, some studies included participants who used clinical symptoms as a COVID-19 case, which is less accurate than a RT-PCR test. It should be mentioned that the recruitment domain could have potentially been high risk for most studies, as in the early stages of the pandemic, RT-PCR testing was relatively scarce, and only individuals with symptoms or who had direct contact with a positive person were tested in some countries. Furthermore, some studies used rough age categories which will lead to an inaccurate risk effect. Lastly, some studies corrected for biomarkers or factors (such as fever, d-Dimer level, lymphocyte count) that reflect disease severity. When studying associations between disease severity and age and chronic factors, using such markers in the model should be avoided to avoid overadjustment.

### Implications for practice, policy, and future research

During the COVID-19 pandemic, it is necessary to accurately define risk groups for COVID-19 disease severity when implementing contact or work restrictions. This review shows that increased age year is associated with mortality and hospitalization, and an age threshold for which there is a marked increase in risk was not apparent. Therefore, targetting persons above a certain (arbitrary) age threshold is not recommended in the workplace, as it may lead to ageism, permeating in the community and in the workplace (100). If workers older than 60 years are not allowed to perform a work task because of perceived higher risk of disease severity starting at that age, it is difficult to argue why a person can still perform that activity at age 59, but must stop it or needs special protection on their 60^th^ birthday. Rather, individualized risk profiles should be made by considering a continuous increase in risk by age-year.

Contrary to workplace restrictions, the use of an isolated, per-age year increase when prioritizing vaccinations is not recommended. Government agencies often divide the population in age groups- but this categorization is not purely about age-related effects. Rather, the high-age groups always include a higher proportion of individuals with pre-exisiting conditions. In this respect, our result of an approximately linear increase in isolated age risk cannot be used as an argument against the definition of vaccination prioritization groups.

This comprehensive review contradicts our previous rapid review (13), which encompassed mainly Chinese studies and indicated a weak influence of age on COVID-19 disease severity. This highlights the need for continuous, “living” reviews of the available evidence throughout the pandemic, as results may change due to more available evidence available or to new variants affecting the association between age and disease severity. Further, as there are hints of possible effect modifications between age and certain chronic diseases (14), future studies should focus on quantifying these interactions.

## Conclusions

A best-possible quantification of the increase in COVID-19 disease severity due to age was achieved. Age-related workplace prevention programs should consider the continuous increase in risk, rather than implementing age thresholds. There is a need for continuous, high-quality research to characterize age associations with disease severity in light of new variants.

## Data Availability

All data used is included in text and supplementary materials.

## Acknowledgments

Our heartfelt thanks to Dr. Thomas Harder and Dr. Marina Treskova from the Robert Koch Institute for their recommendations and for giving us access to their study database.

## Patient and Public Involvement

Patients or the public were not involved in the design, or conduct, or reporting, or dissemination plans of our research.

