## Supplementary Material for "The isolated effect of age on the risk of COVID-19 severe outcomes: a systematic review with meta-analysis"

**Search strategy of Umbrella Review**

Search Syntax PubMed 1:

("Severe Acute Respiratory Syndrome Coronavirus 2" [Supplementary Concept] OR "COVID-19" [Supplementary Concept] OR "covid 19 diagnostic testing" [Supplementary Concept] OR "covid 19 drug treatment" [Supplementary Concept] OR "covid 19 serotherapy"[Supplementary Concept] OR "covid 19 vaccine" [Supplementary Concept] OR "Severe Acute Respiratory Syndrome Coronavirus 2"[tiab] OR ncov*[tiab] OR covid*[tiab] OR sars-cov-2[tiab] OR "sars cov 2"[tiab] OR "SARS Coronavirus 2"[tiab] OR "Severe Acute Respiratory Syndrome CoV 2"[tiab] OR "Wuhan coronavirus"[tiab] OR "Wuhan seafood market pneumonia virus"[tiab] OR "SARS2"[tiab] OR "2019-nCoV"[tiab] OR "hcov-19"[tiab] OR „novel 2019 coronavirus“[tiab] OR "2019 novel coronavirus*"[tiab] OR „novel coronavirus 2019*“[tiab] OR "2019 novel human coronavirus*"[tiab] OR „human coronavirus 2019“[tiab] OR "coronavirus disease-19"[tiab] OR "corona virus disease-19"[tiab] OR "coronavirus disease 2019"[tiab] OR "corona virus disease 2019"[tiab] OR "2019 coronavirus disease"[tiab] OR "2019 corona virus disease"[tiab] OR „novel coronavirus disease 2019“[tiab] OR „novel coronavirus infection 2019“[tiab] OR "new coronavirus*"[tiab] OR "coronavirus outbreak"[tiab] OR "coronavirus epidemic"[tiab] OR "coronavirus pandemic"[tiab] OR "pandemic of coronavirus"[tiab]) AND ("2019/12/01"[PDAT] : "2099/12/31"[PDAT])

Search Syntax PubMed 2:

("wuhan"[tiab] or china[tiab] or hubei[tiab]) AND ("Severe Acute Respiratory Syndrome Coronavirus 2"[Supplementary Concept] OR "COVID-19" [Supplementary Concept] OR "covid 19 diagnostic testing"[Supplementary Concept] OR "covid 19 drug treatment"[Supplementary Concept] OR "covid 19 serotherapy"[Supplementary Concept] OR "covid 19 vaccine"[Supplementary Concept] OR "coronavirus*"[tiab] OR "corona virus*"[tiab] OR ncov[tiab] OR covid*[tiab] OR sars*[tiab])

Search Syntax Embase 1:

('severe acute respiratory syndrome coronavirus 2':ti,ab OR 'severe acute respiratory syndrome coronavirus 2'/exp OR 'covid 19'/exp OR ncov*:ti,ab OR covid*:ti,ab OR 'sars cov 2':ti,ab OR 'sars-cov-2':ti,ab OR 'sars coronavirus 2':ti,ab OR 'sars coronavirus 2'/exp OR 'severe acute respiratory syndrome cov 2':ti,ab OR 'wuhan coronavirus':ti,ab OR 'wuhan seafood market pneumonia virus':ti,ab OR sars2:ti,ab OR '2019-ncov':ti,ab OR 'hcov-19':ti,ab OR 'novel 2019 coronavirus':ti,ab OR '2019 novel coronavirus*':ti,ab OR 'novel coronavirus 2019'/exp OR '2019 novel human coronavirus*':ti,ab OR 'human coronavirus 2019':ti,ab OR 'coronavirus disease-19':ti,ab OR 'corona virus disease-19':ti,ab OR 'coronavirus disease 2019':ti,ab OR 'coronavirus disease 2019'/exp OR 'corona virus disease 2019':ti,ab OR '2019 coronavirus disease':ti,ab OR 'novel coronavirus 2019*':ti,ab OR 'novel coronavirus disease 2019':ti,ab OR 'novel coronavirus infection 2019':ti,ab OR '2019 corona virus disease':ti,ab OR 'new coronavirus*':ti,ab OR 'coronavirus outbreak':ti,ab OR 'coronavirus epidemic':ti,ab OR 'coronavirus pandemic':ti,ab OR 'pandemic of coronavirus':ti,ab OR 'severe acute respiratory syndrome coronavirus 2 vaccine'/exp OR 'covid 19 vaccine'/exp) AND 2020:py

Search Syntax Embase 2:

(wuhan:ti,ab OR china:ti,ab OR hubei:ti,ab) AND ('severe acute respiratory syndrome coronavirus 2':ti,ab OR 'severe acute respiratory syndrome coronavirus 2'/exp OR 'severe acute respiratory syndrome coronavirus 2' OR 'covid*':ti,ab OR 'covid 19'/exp OR 'covid 19' OR coronavirus*:ti,ab OR 'corona virus*':ti,ab OR ncov:ti,ab OR covid*:ti,ab OR sars*:ti,ab OR 'sars coronavirus 2'/exp)

These search terms were combined with “systematic review OR meta-analysis”[all fields] to identify eligible systematic reviews.

Records identified through database searching (last search on 11.12.2020)

n=3,417

Identification

Unique systematic reviews screened after duplicates removed

n=3,335

Screening

Reviews excluded: n=15

5 pre-print duplication

4 no citations for included primary studies

2 citations do not match given references for primary studies

1 estimates of prevalence

1 full-text was not retrievable

Full-text screened for eligibility

n=143

Eligibility

Systematic reviews included

n = 120

Included

**Figure S1. Selection of systematic reviews of Umbrella Review (Treskova et al. 2021)**

Titles identified through extraction the records from 120 reviews, after removing duplications

n =1,134

Identification

Records excluded: n = 778

653 no risk estimates for pre-existing conditions

54 case reports without risk estimates

25 report risk estimates for any undefined comorbidity

2	withdrawn papers

45 full-text was not retrievable

Records screened for risk estimates for pre-existing conditions

n =1,133

Screening

Full-text screened for eligibility based on adjustment for age confounding effect

n = 355

Records excluded by adjustment: n = 84

71 not age-adjusted for pre-existing conditions

12 not reported

1	duplication in content

Records excluded: n = 111

*by outcome*: n = 92

16 incidence or positive test result

9	composite outcome: ICU or death

35 composite outcome: severe disease

32 other outcomes

*by risk factor or population*: n = 19

9	estimates irrespective of SARS-CoV2 status

2	simulated population

1	patients undergoing surgery

2	risk: blood glucose analysis

1	risk: treatment with Tocilizumab

2	risk: visceral fat

1 separate outcome for men and women

1	CRP and CD4 in HIV patients

Full-text screened

n = 271

Eligibility

Studies included for data extraction

n = 160

Included

**Figure 2. Selection of primary studies in umbrella review (Treskova et al. 2021)**

**Risk of Bias Instrument and Explanation**

| **Major risk of bias domains** | **Risk** | **Criteria** | **Hints/ notes** |
| --- | --- | --- | --- |
| **1. Recruitment procedure and follow-up (in cohort studies)**  ***For ecological studies or other analytical studies^†^***  *HINT: We are looking for selection bias:*  *Examples of possible selection bias:*   - *If positive COVID-19 participants are recruited, positivity tested by clinical evaluation, such as typical symptoms. Ideally, participants were tested with RT-PCR laboratory tests to have a low risk of bias.*   *Examples of baseline difference:*   - *Protocols for recruitment or inclusion/exclusion criteria were applied differently across study groups* - *Study participants/populations were recruited at different times* - *Study participants/populations were* *recruited at different times* - *Study participants were recruited from different populations and proportions of participants from each population in each study group are not uniform (consider study base principle)* - *Participation rates were inadequate or not comparable across study groups* | low | ☐ There were no baseline differences among study groups or adjustment techniques were used to correct for the differences, OR there is insufficient information about participant selection, but there is indirect evidence that suggests that participant recruitment and inclusion/exclusion criteria was consistent, as described by the criteria for a judgment of low risk of bias.  ☐ For cohort studies: Loss to follow-up is below 20% in total and not different between the two groups (up to 10% difference). * |  |
|  | high | There were baseline differences among study groups and no adjustment was used to correct for differences, such as (select all that apply):  ☐ Protocols for recruitment or inclusion/exclusion criteria were applied differently across study groups.  ☐ Study participants/populations were recruited at different times.  ☐ Study participants were recruited from different populations and proportions of participants from each population in each study group are not uniform (study base principle not considered).  ☐ Participation rates were inadequate or not comparable across study groups.  ☐ For cohort studies: total loss to follow-up is larger than acceptable (20% or more)* OR drop out differs between the groups by more than 10%* OR the reasons for drop out considerably differ between exposed and non-exposed groups.* |  |
|  | unclear | ☐ There is insufficient information about participant selection to permit a judgment. |  |
| **2. Exposure definition and measurement** | low | ☐ There is high confidence in the accuracy of the exposure assessment^#^ or less established/ less direct exposure measurements are validated against well-established of direct methods^†^ AND  ☐ Exposure measured per year or by 5- or 10-year categories. |  |
|  | high | ☐ Exposure was not accurately measured. ^#^  ☐ Less established or less direct exposure measurements are not validated and are suspected to introduce bias, which may affect the outcome assessment. ^†^  ☐ Exposure used “large/rough” categories (i.e., ≥60 vs. <60 years) |  |
|  | unclear | ☐ Not reported. |  |
| **3. Outcome “hospitalization, ICU admission, mechanical ventilation, case mortality, deaths/mortality”**  **Source and validation** | low | ☐ Outcome was objectively ascertained to minimize bias through objective sources (hospital/medical records). ^#^  ☐ Measurement methods were similar in the different groups. ^#^ |  |
|  | high | ☐ Outcome was not objectively ascertained. ^#^  ☐ Measurement methods were different in the groups. ^#^ |  |
|  | unclear | ☐ Not reported. |  |
| **4. Confounding** | low | ☐ Major age-related risk factors were assessed and accounted for (diabetes, cardiovascular disease, immunosuppression, kidney disease, liver disease, chronic pulmonary disease). | Confounders not assessed: |
|  | high | ☐ Major age-related risk factors were not assessed nor were they accounted for. |  |
|  | unclear | ☐ Not reported. |  |
| **5. Analysis method: methods to reduce research specific bias** | low | ☐ Authors used adequate statistical models to reduce bias (e.g., standardization, adjustment in multivariate model, propensity scoring). |  |
|  | high | ☐ Authors did not use adequate statistical models to reduce bias. |  |
|  | unclear | ☐ Not reported. |  |

| **Minor risk of bias domains*** | **Risk** | **Criteria** | Hints/ notes |
| --- | --- | --- | --- |
| **6. Chronology** | low | ☐ Temporal relation may be established (age-related risk factors precede the outcome). |  |
|  | high | ☐ Temporal relation cannot be established. (laboratory values or other covariates reflecting disease severity are used in model) | Please specify: |
|  | unclear | ☐ Not reported. |  |
| **7. Funding** | low | ☐ Grant/ non-profit-organizations*  ☐ Study was clearly not affected by sponsors. * |  |
|  | high | ☐ Sponsoring organization participated in data analysis.  ☐ Study was probably affected by sponsors. |  |
|  | unclear | ☐ Industry, combined industry+grant*, unclear if study was affected by sponsors.  ☐ Not reported. |  |
| **8. Conflict of interest** | low | ☐ Reported not having conflict of interest or clear from report/communication that study was not affected by author(s) affiliation.* |  |
|  | high | ☐ Conflict of interest exists (at least one author).* |  |
|  | unclear | ☐ Not reported. |  |

| **Overall risk of bias assessment:**  General rule for rating domains: if at least one box in domain is marked for “high risk”, the domain is high risk. | | | **Low Risk** | **High Risk** | **Unclear Risk** |
| --- | --- | --- | --- | --- | --- |
| Major domains | 1. Recruitment procedure and follow-up (in cohort studies) | |  |  |  |
|  | 2. Exposure assessment | |  |  |  |
|  | 3. Outcome source and validation | |  |  |  |
|  | 4. Confounding | |  |  |  |
|  | 5. Analysis method: methods to reduce research specific bias | |  |  |  |
| Minor domains | 6. Chronology | |  |  |  |
|  | 7. Funding | |  |  |  |
|  | 8. Conflict of interest | |  |  |  |
| **General rule for overall rating: Low risk of bias:** low risk in all major domains  **High risk of bias:** if not low risk | | **Overall assessment:** |  |  |  |

**Description of Risk of Bias Domains (1-8)**

1. **Recruitment procedure and follow-up**

A low-risk study should have minimized selection bias by ensuring that there were no baseline differences among the study groups, or if there were, adjustment techniques were used to correct for the difference. If there was indirect evidence suggesting that participant recruitment and that the inclusion and exclusion criteria were consistent, the domain was assessed as having a low risk of bias. In cohort studies, the loss to follow-up should have been less than 20% and this loss should not be different between the groups. If the studies recruited COVID-19 positive participants, preferably a real-time reverse transcription polymerase chain reaction (RT-PCR) test would have been used to test for COVID-19 positivity. If COVID-19 ascertainment methods were unreliable, such as relying on clinical symptoms, the domain was also considered as high risk. If there were baseline differences among the study groups, such as using different inclusion/exclusion criteria, inadequate participation rates or rates that were not comparable across the group, or if the study participants were recruited from different populations and the proportions of participants from each population in each study were not uniform, the domain was considered as high risk.

2. **Exposure definition and measurement**

If age was accurately measured and finely categorized, such as in per year, or in five or ten-year categories, this domain was considered to have a low risk of bias. If age was measured in rough or large categories, greater than in 10-year categories or such as >60 years vs ≤60 years, the domain was considered as having a high risk of bias.

3. **Outcome source and validation**

If the outcome was objectively measured to minimize bias, such as through hospital or medical records, and the assessment was similar for the comparison groups, this domain was assessed as having a low risk of bias.

4. **Age-dependent risk factors**

In order for studies to be considered as having a low risk of bias, the following age-dependent factors should have been considered, based on the current evidence (7, 8): (1) diabetes, (2) cardiovascular disease, (3) immunosuppression/cancer, (4) chronic respiratory diseases, such as chronic obstructive pulmonary disease (COPD), (5) chronic kidney disease, and (6) chronic liver disease. Risk factors for COVID-19 disease severity with no or little age-dependency (such as sex, obesity, smoking) were not considered age-dependent risk factors.

5. **Analysis methods**

If adequate statistical models were used to reduce bias and control for confounding, this domain was considered as having a low risk of bias.

6. **Chronology**

If temporal relation may be established, especially considering that the age-related risk factors used in the model precede the outcome, this domain was assessed as having a low risk of bias. If there were covariates in the model reflecting disease severity (such as fever, lymphocite counts, D-dimer level, etc,) the domain was considered as having high risk.

7. **Funding**

Funding was assessed in two areas, namely the funding source and its involvement in the research. If a study was funded by non-profit organization(s) and it was not affected by sponsors, the domain was considered low risk. If the funding organization participated in the data analysis or the study was probably affected by the sponsors, the domain had a high risk of bias.

8. **Conflict of interest**

If the authors reported no conflicts of interest, this domain was low risk. If one author had a conflict of interest, this domain was assessed as having a high risk of bias.

**Overall Risk of Bias**

From the above domains, domains 1-5 were set as major domains, while domains 6-8 were minor domains. A study could obtain an overall low risk of bias if all major domains were assessed as having low risk of bias.

**Table S1. Excluded studies and reasons for exclusion**

| **Reference** | **(st: Stratification of age only (no adjustment in model); co: Comorbidities missing; p: Special population (i.e. cancer patients, patients with diabetes); a: Age effect estimate not provided; ag: age category unclear; o: Other)** | **Reason for exclusion** | | | | | |
| --- | --- | --- | --- | --- | --- | --- | --- |
|  |  | **st** | **co** | **p** | **a** | **ag** | **o** |
| Al‐Sabah, S., Al‐Haddad, M., Al‐Youha, S., Jamal, M., & Almazeedi, S. (2020). COVID‐19: Impact of obesity and diabetes on disease severity. Clinical Obesity, 10(6), e12414. | |  | x |  |  |  |  |
| Al‐Salameh, A., Lanoix, J. P., Bennis, Y., Andrejak, C., Brochot, E., Deschasse, G., ... & Lalau, J. D. (2020). Characteristics and outcomes of COVID‐19 in hospitalized patients with and without diabetes. Diabetes/Metabolism Research and Reviews, e3388. | |  | x |  |  |  |  |
| Amit, M., Sorkin, A., Chen, J., Cohen, B., Karol, D., Tsur, A. M., ... & Benov, A. (2020). Clinical course and outcomes of severe Covid-19: a national scale study. Journal of clinical medicine, 9(7), 2282. | |  | x |  |  |  |  |
| Bellan, M., Patti, G., Hayden, E., Azzolina, D., Pirisi, M., Acquaviva, A., ... & Sainaghi, P. P. (2020). Fatality rate and predictors of mortality in an Italian cohort of hospitalized COVID-19 patients. Scientific reports, 10(1), 1-10. | |  | x |  |  |  |  |
| Berenguer, J., Ryan, P., Rodríguez-Baño, J., Jarrín, I., Carratalà, J., Pachón, J., ... & Jofre, C. S. (2020). Characteristics and predictors of death among 4035 consecutively hospitalized patients with COVID-19 in Spain. Clinical Microbiology and Infection, 26(11), 1525-1536. | |  | x |  |  |  |  |
| Bezzio, C., Saibeni, S., Variola, A., Allocca, M., Massari, A., Gerardi, V., ... & Fiorino, G. (2020). Outcomes of COVID-19 in 79 patients with IBD in Italy: an IG-IBD study. Gut, 69(7), 1213-1217. | |  |  | x |  |  |  |
| Bianchetti, A., Rozzini, R., Guerini, F., Boffelli, S., Ranieri, P., Minelli, G., ... & Trabucchi, M. (2020). Clinical presentation of COVID19 in dementia patients. The journal of nutrition, health & aging, 24, 560-562. | |  | x |  |  |  |  |
| Borghesi, A., Zigliani, A., Golemi, S., Carapella, N., Maculotti, P., Farina, D., & Maroldi, R. (2020). Chest X-ray severity index as a predictor of in-hospital mortality in coronavirus disease 2019: A study of 302 patients from Italy. International Journal of Infectious Diseases, 96, 291-293. | |  | x |  |  |  |  |
| Busetto, L., Bettini, S., Fabris, R., Serra, R., Dal Pra, C., Maffei, P., ... & Vettor, R. (2020). Obesity and COVID‐19: an Italian snapshot. Obesity, 28(9), 1600-1605. | |  | x |  |  |  |  |
| Cariou, B., Hadjadj, S., Wargny, M., Pichelin, M., Al-Salameh, A., Allix, I., ... & Gourdy, P. (2020). Phenotypic characteristics and prognosis of inpatients with COVID-19 and diabetes: the CORONADO study. Diabetologia, 63(8), 1500-1515. | |  |  | x |  |  |  |
| Caussy, C., Pattou, F., Wallet, F., Simon, C., Chalopin, S., Telliam, C., ... & Disse, E. (2020). Prevalence of obesity among adult inpatients with COVID-19 in France. The Lancet Diabetes & Endocrinology, 8(7), 562-564. | |  |  |  | x |  |  |
| Chen, F., Sun, W., Sun, S., Li, Z., Wang, Z., & Yu, L. (2020). Clinical characteristics and risk factors for mortality among inpatients with COVID‐19 in Wuhan, China. Clinical and translational medicine. | |  | x |  |  |  |  |
| Chen, R., Liang, W., Jiang, M., Guan, W., Zhan, C., Wang, T., ... & for COVID, M. T. E. G. (2020). Risk factors of fatal outcome in hospitalized subjects with coronavirus disease 2019 from a nationwide analysis in China. Chest, 158(1), 97-105. | |  | x |  |  |  |  |
| Ciceri, F., Castagna, A., Rovere-Querini, P., De Cobelli, F., Ruggeri, A., Galli, L., ... & Zangrillo, A. (2020). Early predictors of clinical outcomes of COVID-19 outbreak in Milan, Italy. Clinical Immunology, 217, 108509. | |  | x |  |  |  |  |
| Crouse, A. B., Grimes, T., Li, P., Might, M., Ovalle, F., & Shalev, A. (2021). Metformin use is associated with reduced mortality in a diverse population with COVID-19 and diabetes. Frontiers in Endocrinology, 11, 1081. | |  | x |  |  |  |  |
| D’Silva, K. M., Serling-Boyd, N., Wallwork, R., Hsu, T., Fu, X., Gravallese, E. M., ... & Wallace, Z. S. (2020). Clinical characteristics and outcomes of patients with coronavirus disease 2019 (COVID-19) and rheumatic disease: a comparative cohort study from a US ‘hot spot’. Annals of the rheumatic diseases, 79(9), 1156-1162. | |  |  |  | x |  |  |
| Dai, M., Liu, D., Liu, M., Zhou, F., Li, G., Chen, Z., ... & Cai, H. (2020). Patients with cancer appear more vulnerable to SARS-CoV-2: a multicenter study during the COVID-19 outbreak. Cancer discovery, 10(6), 783-791. | |  | x |  |  |  |  |
| Denova‐Gutiérrez, E., Lopez‐Gatell, H., Alomia‐Zegarra, J. L., López‐Ridaura, R., Zaragoza‐Jimenez, C. A., Dyer‐Leal, D. D., ... & Barquera, S. (2020). The association of obesity, type 2 Diabetes, and hypertension with severe coronavirus disease 2019 on admission among Mexican patients. Obesity, 28(10), 1826-1832. | |  | x |  |  |  |  |
| Du, R. H., Liang, L. R., Yang, C. Q., Wang, W., Cao, T. Z., Li, M., ... & Shi, H. Z. (2020). Predictors of mortality for patients with COVID-19 pneumonia caused by SARS-CoV-2: a prospective cohort study. European Respiratory Journal, 55(5). | |  | x |  |  |  |  |
| Escalera-Antezana, J. P., Lizon-Ferrufino, N. F., Maldonado-Alanoca, A., Alarcon-De-la-Vega, G., Alvarado-Arnez, L. E., Balderrama-Saavedra, M. A., ... & Rodriguez-Morales, A. J. (2020). Risk factors for mortality in patients with Coronavirus Disease 2019 (COVID-19) in Bolivia: An analysis of the first 107 confirmed cases. Infez Med, 28(2), 238-242. | |  | x |  |  |  |  |
| Galloway, J. B., Norton, S., Barker, R. D., Brookes, A., Carey, I., Clarke, B. D., ... & Cantle, F. (2020). A clinical risk score to identify patients with COVID-19 at high risk of critical care admission or death: an observational cohort study. Journal of Infection, 81(2), 282-288. | | x |  |  |  |  |  |
| Gao, C., Cai, Y., Zhang, K., Zhou, L., Zhang, Y., Zhang, X., ... & Li, F. (2020). Association of hypertension and antihypertensive treatment with COVID-19 mortality: a retrospective observational study. European heart journal, 41(22), 2058-2066. | |  | x |  |  |  |  |
| Garassino, M. C., Whisenant, J. G., Huang, L. C., Trama, A., Torri, V., Agustoni, F., ... & Horn, L. (2020). COVID-19 in patients with thoracic malignancies (TERAVOLT): first results of an international, registry-based, cohort study. The Lancet Oncology, 21(7), 914-922. | |  | x |  |  |  |  |
| Garcia, P. D. W., Fumeaux, T., Guerci, P., Heuberger, D. M., Montomoli, J., Roche-Campo, F., ... & RISC-19-ICU Investigators. (2020). Prognostic factors associated with mortality risk and disease progression in 639 critically ill patients with COVID-19 in Europe: Initial report of the international RISC-19-ICU prospective observational cohort. EClinicalMedicine, 25, 100449. | | x |  |  |  |  |  |
| Gayam, V., Chobufo, M. D., Merghani, M. A., Lamichhane, S., Garlapati, P. R., & Adler, M. K. (2021). Clinical characteristics and predictors of mortality in African‐Americans with COVID‐19 from an inner‐city community teaching hospital in New York. Journal of medical virology, 93(2), 812-819. | |  | x |  |  |  |  |
| Geretti, A. M., Stockdale, A. J., Kelly, S. H., Cevik, M., Collins, S., Waters, L., ... & Semple, M. G. (2020). Outcomes of COVID-19 related hospitalization among people with HIV in the ISARIC WHO Clinical Characterization Protocol (UK): a prospective observational study. Clinical Infectious Diseases. | |  |  | x |  |  |  |
| Goicoechea, M., Cámara, L. A. S., Macías, N., de Morales, A. M., Rojas, Á. G., Bascuñana, A., ... & Aragoncillo, I. (2020). COVID-19: clinical course and outcomes of 36 hemodialysis patients in Spain. Kidney international, 98(1), 27-34. | |  | x |  |  |  |  |
| Goyal, P., Ringel, J. B., Rajan, M., Choi, J. J., Pinheiro, L. C., Li, H. A., ... & Safford, M. M. (2020). Obesity and COVID-19 in New York City: a retrospective cohort study. Annals of Internal Medicine, 173(10), 855-858. | |  |  |  | x |  |  |
| Halasz, G., Leoni, M. L., Villani, G. Q., Nolli, M., & Villani, M. (2020). Obesity, overweight and survival in critically ill patients with SARS-CoV-2 pneumonia: is there an obesity paradox? Preliminary results from Italy. European Journal of Preventive Cardiology, 2047487320939675. | |  |  |  | x |  |  |
| Harmouch, F., Shah, K., Hippen, J. T., Kumar, A., & Goel, H. (2021). Is it all in the heart? Myocardial injury as major predictor of mortality among hospitalized COVID‐19 patients. Journal of medical virology, 93(2), 973-982. | |  | x |  |  |  |  |
| Huang, S., Wang, J., Liu, F., Liu, J., Cao, G., Yang, C., ... & Xiong, B. (2020). COVID-19 patients with hypertension have more severe disease: a multicenter retrospective observational study. Hypertension Research, 43(8), 824-831. | |  | x |  |  |  |  |
| Hur, K., Price, C. P., Gray, E. L., Gulati, R. K., Maksimoski, M., Racette, S. D., ... & Khanwalkar, A. R. (2020). <? covid19?> Factors Associated With Intubation and Prolonged Intubation in Hospitalized Patients With COVID-19. Otolaryngology–Head and Neck Surgery, 163(1), 170-178. | |  | x |  |  |  |  |
| Kim, M. K., Jeon, J. H., Kim, S. W., Moon, J. S., Cho, N. H., Han, E., ... & Lee, J. H. (2020). The clinical characteristics and outcomes of patients with moderate-to-severe coronavirus disease 2019 infection and diabetes in Daegu, South Korea. Diabetes & metabolism journal, 44(4), 602. | |  |  |  | x |  |  |
| Kuderer, N. M., Choueiri, T. K., Shah, D. P., Shyr, Y., Rubinstein, S. M., Rivera, D. R., ... & Loaiza-Bonilla, A. (2020). Clinical impact of COVID-19 on patients with cancer (CCC19): a cohort study. The Lancet, 395(10241), 1907-1918. | |  | x |  |  |  |  |
| Lee, L. Y., Cazier, J. B., Starkey, T., Turnbull, C. D., Team, U. C. C. M. P., Kerr, R., & Middleton, G. (2020). COVID-19 mortality in patients with cancer on chemotherapy or other anticancer treatments: a prospective cohort study. The Lancet, 395(10241), 1919-1926. | |  |  | x |  |  |  |
| Lee, L. Y., Cazier, J. B., Starkey, T., Briggs, S. E., Arnold, R., Bisht, V., ... & Wyatt, S. (2020). COVID-19 prevalence and mortality in patients with cancer and the effect of primary tumour subtype and patient demographics: a prospective cohort study. The lancet oncology, 21(10), 1309-1316. | |  |  |  | x |  |  |
| Li, Q., Chen, L., Li, Q., He, W., Yu, J., Chen, L., ... & Hu, Y. (2020). Cancer increases risk of in-hospital death from COVID-19 in persons< 65 years and those not in complete remission. Leukemia, 34(9), 2384-2391. | |  | x |  |  |  |  |
| Lieberman-Cribbin, W., Rapp, J., Alpert, N., Tuminello, S., & Taioli, E. (2020). The impact of asthma on mortality in patients with COVID-19. Chest, 158(6), 2290-2291. | |  | x |  |  |  |  |
| Mahdavinia, M., Foster, K. J., Jauregui, E., Moore, D., Adnan, D., Andy-Nweye, A. B., ... & Bishehsari, F. (2020). Asthma prolongs intubation in COVID-19. The Journal of Allergy and Clinical Immunology: In Practice, 8(7), 2388-2391. | |  |  |  | x |  |  |
| Mato, A. R., Roeker, L. E., Lamanna, N., Allan, J. N., Leslie, L., Pagel, J. M., ... & Eyre, T. A. (2020). Outcomes of COVID-19 in patients with CLL: a multicenter international experience. Blood, The Journal of the American Society of Hematology, 136(10), 1134-1143. | |  |  | x |  |  |  |
| McCarty, T. R., Hathorn, K. E., Redd, W. D., Rodriguez, N. J., Zhou, J. C., Bazarbashi, A. N., ... & Chan, W. W. (2020). How do presenting symptoms and outcomes differ by race/ethnicity among hospitalized patients with COVID-19 infection? Experience in Massachusetts. Clinical Infectious Diseases. | |  |  |  |  | x |  |
| Mehta, V., Goel, S., Kabarriti, R., Cole, D., Goldfinger, M., Acuna-Villaorduna, A., ... & Verma, A. (2020). Case fatality rate of cancer patients with COVID-19 in a New York hospital system. Cancer discovery, 10(7), 935-941. | |  |  | x |  |  |  |
| Mendy, A., Apewokin, S., Wells, A. A., & Morrow, A. L. (2020). Factors associated with hospitalization and disease severity in a racially and ethnically diverse population of COVID-19 patients. MedRxiv. | |  | x |  |  |  |  |
| Mikami, T., Miyashita, H., Yamada, T., Harrington, M., Steinberg, D., Dunn, A., & Siau, E. (2021). Risk factors for mortality in patients with COVID-19 in New York City. Journal of general internal medicine, 36(1), 17-26. | |  | x |  |  |  |  |
| Miyashita, H., & Kuno, T. (2020). Prognosis of coronavirus disease 2019 (COVID‐19) in patients with HIV infection in New York City. HIV medicine. | |  | x |  |  |  |  |
| Miyashita, H., Mikami, T., Chopra, N., Yamada, T., Chernyavsky, S., Rizk, D., & Cruz, C. (2020). Do patients with cancer have a poorer prognosis of COVID-19? An experience in New York City. Annals of Oncology. | |  | x |  |  |  |  |
| Miyashita, S., Yamada, T., Mikami, T., Miyashita, H., Chopra, N., & Rizk, D. (2020). Impact of dementia on clinical outcomes in elderly patients with coronavirus 2019 (COVID‐19): an experience in New York. Geriatrics & Gerontology International, 20(7), 732. | |  |  |  | x |  |  |
| Moon, S. S., Lee, K., Park, J., Yun, S., Lee, Y. S., & Lee, D. S. (2020). Clinical characteristics and mortality predictors of COVID-19 patients hospitalized at nationally-designated treatment hospitals. Journal of Korean Medical Science, 35(36). | |  | x |  |  |  |  |
| Nakeshbandi, M., Maini, R., Daniel, P., Rosengarten, S., Parmar, P., Wilson, C., ... & Breitman, I. (2020). The impact of obesity on COVID-19 complications: a retrospective cohort study. International Journal of Obesity, 44(9), 1832-1837. | |  | x |  |  |  |  |
| Okoh, A. K., Sossou, C., Dangayach, N. S., Meledathu, S., Phillips, O., Raczek, C., ... & Grewal, H. S. (2020). Coronavirus disease 19 in minority populations of Newark, New Jersey. International journal for equity in health, 19(1), 1-8. | |  | x |  |  |  |  |
| Passamonti, F., Cattaneo, C., Arcaini, L., Bruna, R., Cavo, M., Merli, F., ... & ITA-HEMA-COV Investigators. (2020). Clinical characteristics and risk factors associated with COVID-19 severity in patients with haematological malignancies in Italy: a retrospective, multicentre, cohort study. The Lancet Haematology, 7(10), e737-e745. | |  |  | x |  |  |  |
| Pettit, N. N., MacKenzie, E. L., Ridgway, J. P., Pursell, K., Ash, D., Patel, B., & Pho, M. T. (2020). Obesity is associated with increased risk for mortality among hospitalized patients with COVID‐19. Obesity, 28(10), 1806-1810. | |  |  |  |  | x |  |
| Pinato, D. J., Zambelli, A., Bower, M., Sng, C. C., Salazar, R., Bertuzzi, A., ... & Gennari, A. (2020). Clinical portrait of the SARS-CoV-2 epidemic in European patients with cancer. Cancer discovery, 10(10), 1465-1474. | |  |  | x |  |  |  |
| Rastad, H., Karim, H., Ejtahed, H. S., Tajbakhsh, R., Noorisepehr, M., Babaei, M., ... & Qorbani, M. (2020). Risk and predictors of in-hospital mortality from COVID-19 in patients with diabetes and cardiovascular disease. Diabetology & metabolic syndrome, 12(1), 1-11. | |  |  | x |  |  |  |
| Regina, J., Papadimitriou-Olivgeris, M., Burger, R., Le Pogam, M. A., Niemi, T., Filippidis, P., ... & Lhopitallier, L. (2020). Epidemiology, risk factors and clinical course of SARS-CoV-2 infected patients in a Swiss university hospital: an observational retrospective study. Plos one, 15(11), e0240781. | |  | x |  |  |  |  |
| Rivera-Izquierdo, M., del Carmen Valero-Ubierna, M., R-delAmo, J. L., Fernández-García, M. Á., Martínez-Diz, S., Tahery-Mahmoud, A., ... & Jiménez-Mejías, E. (2020). Sociodemographic, clinical and laboratory factors on admission associated with COVID-19 mortality in hospitalized patients: A retrospective observational study. PLoS One, 15(6), e0235107. | |  | x |  |  |  |  |
| Robilotti, E. V., Babady, N. E., Mead, P. A., Rolling, T., Perez-Johnston, R., Bernardes, M., ... & Kamboj, M. (2020). Determinants of COVID-19 disease severity in patients with cancer. Nature medicine, 26(8), 1218-1223. | |  |  | x |  |  |  |
| Rossi, A. P., Gottin, L., Donadello, K., Schweiger, V., Nocini, R., Taiana, M., ... & Polati, E. (2020). Obesity as a risk factor for unfavourable outcomes in critically ill patients affected by Covid-19 related respiratory failure: clinical relevance and potential pathophysiological mechanism. | |  |  |  | x |  |  |
| Rottoli, M., Bernante, P., Belvedere, A., Balsamo, F., Garelli, S., Giannella, M., ... & Bartoletti, M. (2020). How important is obesity as a risk factor for respiratory failure, intensive care admission and death in hospitalised COVID-19 patients? Results from a single Italian centre. European Journal of Endocrinology, 183(4), 389-397. | |  | x |  |  |  |  |
| Russell, B., Moss, C., Papa, S., Irshad, S., Ross, P., Spicer, J., ... & Van Hemelrijck, M. (2020). Factors affecting COVID-19 outcomes in cancer patients: a first report from Guy's Cancer Center in London. Frontiers in oncology, 10, 1279. | |  |  | x |  |  |  |
| Sanchez‐Pina, J. M., Rodríguez Rodriguez, M., Castro Quismondo, N., Gil Manso, R., Colmenares, R., Gil Alos, D., ... & Calbacho, M. (2020). Clinical course and risk factors for mortality from COVID‐19 in patients with haematological malignancies. European journal of haematology, 105(5), 597-607. | |  |  | x |  |  |  |
| Shah, V., Ko Ko, T., Zuckerman, M., Vidler, J., Sharif, S., Mehra, V., ... & Kulasekararaj, A. G. (2020). Poor outcome and prolonged persistence of SARS‐CoV‐2 RNA in COVID‐19 patients with haematological malignancies; King's College Hospital experience. British journal of haematology, 190(5), e279-e282. | |  |  |  | x |  |  |
| Shang, Y., Liu, T., Wei, Y., Li, J., Shao, L., Liu, M., ... & Zhou, F. (2020). Scoring systems for predicting mortality for severe patients with COVID-19. EClinicalMedicine, 24, 100426. | |  | x |  |  |  |  |
| Shi, Q., Zhang, X., Jiang, F., Zhang, X., Hu, N., Bimu, C., ... & Wang, W. (2020). Clinical characteristics and risk factors for mortality of COVID-19 patients with diabetes in Wuhan, China: a two-center, retrospective study. Diabetes care, 43(7), 1382-1391. | |  |  | x |  |  |  |
| Sigel, K., Swartz, T., Golden, E., Paranjpe, I., Somani, S., Richter, F., ... & Glicksberg, B. S. (2020). Covid-19 and people with HIV infection: outcomes for hospitalized patients in New York City. Clinical Infectious Diseases. | |  | x |  |  |  |  |
| Simonnet, A., Chetboun, M., Poissy, J., Raverdy, V., Noulette, J., Duhamel, A., ... & Verkindt, H. (2020). High prevalence of obesity in severe acute respiratory syndrome coronavirus‐2 (SARS‐CoV‐2) requiring invasive mechanical ventilation. Obesity, 28(7), 1195-1199. | |  | x |  |  |  |  |
| Singh, S., Bilal, M., Khan, A., Chowdhry, M., Sánchez-Luna, S. A., Kochhar, G. S., ... & Thompson, C. C. (2020). Outcomes of COVID-19 in Patients with Obesity in United States: A Large Research Network Study. Available at SSRN 3611983. | |  |  |  | x |  |  |
| Singh, S., & Khan, A. (2020). Clinical characteristics and outcomes of coronavirus disease 2019 among patients with preexisting liver disease in the United States: a multicenter research network study. Gastroenterology, 159(2), 768-771. | |  |  |  | x |  |  |
| Singh, S., Khan, A., Chowdhry, M., Bilal, M., Kochhar, G. S., & Clarke, K. (2020). Risk of severe coronavirus disease 2019 in patients with inflammatory bowel disease in the United States: a multicenter research network study. Gastroenterology, 159(4), 1575-1578. | |  |  |  | x |  |  |
| Sisó-Almirall, A., Kostov, B., Mas-Heredia, M., Vilanova-Rotllan, S., Sequeira-Aymar, E., Sans-Corrales, M., ... & Benavent-Àreu, J. (2020). Prognostic factors in Spanish COVID-19 patients: A case series from Barcelona. PloS one, 15(8), e0237960. | | x |  |  |  |  |  |
| Smith, A. A., Fridling, J., Ibhrahim, D., & Porter Jr, P. S. (2020). Identifying patients at greatest risk of mortality due to COVID-19: A New England perspective. Western Journal of Emergency Medicine, 21(4), 785. | |  | x |  |  |  |  |
| Sun, H., Ning, R., Tao, Y., Yu, C., Deng, X., Zhao, C., ... & Xu, D. (2020). Risk factors for mortality in 244 older adults with COVID‐19 in Wuhan, China: a retrospective study. Journal of the American Geriatrics Society, 68(6), E19-E23. | |  | x |  |  |  |  |
| Sy, K. T. L., Haw, N. J. L., & Uy, J. (2020). Previous and active tuberculosis increases risk of death and prolongs recovery in patients with COVID-19. Infectious Diseases, 52(12), 902-907. | |  |  |  | x |  |  |
| Wang, D., Yin, Y., Hu, C., Liu, X., Zhang, X., Zhou, S., ... & Peng, Z. (2020). Clinical course and outcome of 107 patients infected with the novel coronavirus, SARS-CoV-2, discharged from two hospitals in Wuhan, China. Critical Care, 24, 1-9. | |  | x |  |  |  |  |
| Wang, K., Zuo, P., Liu, Y., Zhang, M., Zhao, X., Xie, S., ... & Liu, C. (2020). Clinical and Laboratory Predictors of In-hospital Mortality in Patients With Coronavirus Disease-2019: A Cohort Study in Wuhan, China. Clinical infectious diseases, 71(16), 2079-2088. | |  | x |  |  |  |  |
| Xie, J., Covassin, N., Fan, Z., Singh, P., Gao, W., Li, G., ... & Somers, V. K. (2020, June). Association between hypoxemia and mortality in patients with COVID-19. In Mayo Clinic Proceedings (Vol. 95, No. 6, pp. 1138-1147). Elsevier. | |  | x |  |  |  |  |
| Xu, J., Yang, X., Yang, L., Zou, X., Wang, Y., Wu, Y., ... & Shang, Y. (2020). Clinical course and predictors of 60-day mortality in 239 critically ill patients with COVID-19: a multicenter retrospective study from Wuhan, China. Critical Care, 24(1), 1-11. | |  | x |  |  |  |  |
| Yan, Y., Yang, Y., Wang, F., Ren, H., Zhang, S., Shi, X., ... & Dong, K. (2020). Clinical characteristics and outcomes of patients with severe covid-19 with diabetes. BMJ open diabetes research and care, 8(1), e001343. | |  |  |  | x |  |  |
| Yang, Q., Zhou, Y., Wang, X., Gao, S., Xiao, Y., Zhang, W., ... & Wang, Y. (2020). Effect of hypertension on outcomes of adult inpatients with COVID-19 in Wuhan, China: a propensity score–matching analysis. Respiratory research, 21(1), 1-9. | |  |  |  | x |  |  |
| Yarza, R., Bover, M., Paredes, D., López-López, F., Jara-Casas, D., Castelo-Loureiro, A., ... & Gómez-Martín, C. (2020). SARS-CoV-2 infection in cancer patients undergoing active treatment: analysis of clinical features and predictive factors for severe respiratory failure and death. European journal of cancer, 135, 242-250. | |  |  | x |  |  |  |
| Yu, C., Lei, Q., Li, W., Wang, X., Liu, W., Fan, X., & Li, W. (2020). Clinical characteristics, associated factors, and predicting COVID-19 mortality risk: a retrospective study in Wuhan, China. American journal of preventive medicine, 59(2), 168-175. | |  | x |  |  |  |  |
| Yun, F., Yun, L., Tao, B., Yusang, X., Jie, H., Jian, L., ... & Hongzhou, L. (2020). COVID-19 with Different Severity: A Multi-center Study of Clinical Features. | |  | x |  |  |  |  |
| Zhang, F., Xiong, Y., Wei, Y., Hu, Y., Wang, F., Li, G., ... & Zhu, W. (2020). Obesity predisposes to the risk of higher mortality in young COVID‐19 patients. Journal of medical virology, 92(11), 2536-2542. | |  | x |  |  |  |  |
| Zhang, H., Wang, L., Chen, Y., Wu, Q., Chen, G., Shen, X., ... & Xie, C. (2020). Outcomes of novel coronavirus disease 2019 (COVID‐19) infection in 107 patients with cancer from Wuhan, China. Cancer, 126(17), 4023-4031. | |  |  | x |  |  |  |
| Zhang, P., Zhu, L., Cai, J., Lei, F., Qin, J. J., Xie, J., ... & Li, H. (2020). Association of inpatient use of angiotensin-converting enzyme inhibitors and angiotensin II receptor blockers with mortality among patients with hypertension hospitalized with COVID-19. Circulation research, 126(12), 1671-1681. | |  |  |  | x |  |  |
| Zhang, X., Guo, W., Hua, J., Luo, Z., Gao, S., Ran, L., ... & Wang, F. (2020). The incidence, risk factors and clinical outcomes of acute kidney injury in critically ill patients with COVID-19: A multicenter study. | |  |  |  | x |  |  |
| Zhang, Y., Cui, Y., Shen, M., Zhang, J., Liu, B., Dai, M., ... & Pan, P. (2020). Association of diabetes mellitus with disease severity and prognosis in COVID-19: a retrospective cohort study. Diabetes research and clinical practice, 165, 108227. | |  |  |  | x |  |  |
| Zhou, F., Yu, T., Du, R., Fan, G., Liu, Y., Liu, Z., ... & Cao, B. (2020). Clinical course and risk factors for mortality of adult inpatients with COVID-19 in Wuhan, China: a retrospective cohort study. The lancet, 395(10229), 1054-1062. | |  | x |  |  |  |  |
| Zhu, L., She, Z. G., Cheng, X., Qin, J. J., Zhang, X. J., Cai, J., ... & Li, H. (2020). Association of blood glucose control and outcomes in patients with COVID-19 and pre-existing type 2 diabetes. Cell metabolism, 31(6), 1068-1077. | |  |  |  | x |  |  |
| Zimmerman, P., Stroever, S., Burton, T., Hester, K., Kim, M., Fahy, R., ... & Nicastro, J. (2020). Mortality associated with intubation and mechanical ventilation in patients with COVID-19. MedRxiv. | |  | x |  |  |  |  |

**Table S2. Characteristics of the included studies**

| **Author, Year**  **Country**  **(Ref)** | **Study**  **Design** | **Population**  **Sampling**  **Participants** | **Age/Sex Characteristics** | **Time period of study** | **Age categories used** | **Outcome**  **Outcome ascertainment** |
| --- | --- | --- | --- | --- | --- | --- |
| Antwi-Amoabeng  2020  USA  (165) | Retrospective cohort | Patients in Renown Regional Medical Center (a referral hospital serving most of Northern Nevada) and affiliated satellite hospitals.  All people who had a positive nasopharyngeal RT-PCR test for SARS-COV-2.  N= 172 | Mean age:  53 yrs.  Range: 33.5-68 yrs.  Male: 96 (55.8%)  Female: 76 (44.2%) | March 12- May 8, 2020 | ≤61 yrs. vs.  >61 yrs. | Mortality (in hospital)  Data extracted from electronic health records |
| Argenziano  2020  USA  (166) | Prospective cohort | First 1000 consecutive adult COVID-19 patients at New York-Presbyterian/ Columbia University Irving Medical Center (NYP/CUIMC) registered in the emergency department or as inpatients.  Laboratory-confirmed COVID-19 infection confirmed by SARS-CoV-2 RT-PCR test  N=1000 | Median age:  63.0 yrs.  IQR: 50.0-75.0 yrs.  Male: 596 (59.6%)  Female: 404 (41.4%) | March 11-April 6^th^, 2020 followed up until April 30, 2020 | Per year | ICU admission  In-hospital mortality  Hospital records |
| Azar  2020  USA  (167) | Retrospective cohort | Sutter health system, delivering comprehensive medical services in ambulatory clinics and acute care hospitals in North California.  Patients aged 18 or older with a positive test result in the HER laboratory records or patients who had a documented ICD-10 diagnosis of confirmed COVID-19 in the electronic health record without a positive test result.  N= 1052 | Mean age:  53.0 yrs.  95% CI: 51.8-54.1 yrs.  Male: 518 (49.2%)  Female: 534 (50.8%) | COVID-19 diagnosis January 1 – April 8, 2020 | 18-39 years  40-49 yrs.  50-59 yrs.  60-69 yrs.  70-79 yrs.  80+ yrs. | Hospitalization  Data extracted from the Sutter electronic health records |
| Baqui  2020  Brazil  (168) | Retrospective cohort | Hospitalized patients in North Region and Central-south region in Brazil with a positive RT-PCR test for severe acute respiratory syndrome coronavirus 2 and who had ethnicity information in the data set SIVEP-Grip (Sistema de Informaçāo de Vigilância Epidemiológica da Gripe).  N= 6882 | Mean age (SD)  North region:  Survivors: 46.9 yrs. (19.3)  Non-survivors: 65.3 yrs. (16.0)  Central-south region:  Survivors: 52.2 yrs. (16.6)  Non-survivors: 67.9 yrs. (15.8)  Male: 58.2%  Female: 42.8% | February 27- May 4, 2020 | <40 yrs.  40-50 yrs.  50-60 yrs.  60-70 yrs.  ≥70 yrs. | Mortality (in-hospital)  Information on death through SIVEP-Grip |
| Bello-Chavolla  2020  Mexico  (170) | Retrospective cohort | Individuals with confirmed cases of COVID-19 from the General Directorate of Epidemiology of the Mexican Ministry of Health.  N= 51, 633 | Mean age (SD)  46.65 yrs. (15.83)  Male: 29,803 (57.7%)  Female:21,830 (42.3%) | Until May 18, 2020 | <40 yrs.  ≥40yrs. - ≤65 yrs.  >65 yrs. | 30-day mortality (population based)  Data from Mexican Ministry of Health |
| Bi, Hong, and Meng  2020  China  (173) | Prospective cohort | Patients at Shenzhen Third People’s Hospital, the designated hospital to treat all patients diagnosed with COVID-19 in Shenzhen, China, regardless of clinical severity and symptom profile.  N= 420 | Mean age (IQR)  45 yrs. (34.60)  Male: 200 (47.6%)  Female: 220 (52.4%) | January 11- March 10, 2020 | 0-39 yrs.  40-59 yrs.  60+ | ICU Admission (hospital based)  Hospital records |
| Boulle  2020  South Africa  (176) | Prospective cohort | Public sector patients aged ≥20 years with documented sex and not known to have died before March 1, 2020 (before first diagnosed COVID-19 case) in the Western Cape Provincial Health Data Centre (WCPHDC).  COVID-19 diagnosis based on a positive SARS-CoV-2 PCR test, available to all until June 1, 2020 and then restricted to patients requiring admission or aged >55 years or with comorbidities.  All public-sector SARS-Cov-2 cases diagnosed before June 1, 2020  N= 15,203  Hospitalized public-sector SARS-CoV-2 cases  N= 2978 | Age  All public-sector SARS-Cov-2 cases diagnosed before June 1, 2020  Survivors  20-39 yrs: 8,653 (59%)  40-49 yrs.: 2,991 (20%)  50-59 yrs.: 1,881 (13%)  60-69 yrs.: 739 (5%)  ≥70 yrs.: 429 (3%)  Non-survivors  20-39 yrs: 35 (7%)  40-49 yrs.: 51 (10%)  50-59 yrs.: 137 (27%)  60-69 yrs.: 150 (29%)  ≥ 70 yrs.: 137 (27%) | Deaths 100 days since March 1, 2020 | 20-39 yrs.  40-49 yrs.  50-59 yrs.  60-69 yrs.  ≥70 yrs. | Mortality by 100 days for:  -all adult cases diagnosed before June 1, 2020 (population-based mortality)  and  -hospitalized adult COVID-19 cases (in-hospital mortality)  Ascertainment through WCPHDC |
| Burn  2020  Spain  (177) | Retrospective cohort | Individuals recorded in the Information System for Research in Primary Care database (SIDIAP) in Catalonia, Spain, with at least one year of prior history available, no prior diagnosis for COVID-19 and who were not hospitalized on March 1, 2020 included. SIDIAP captures patient records from 80% of the Catalan population, and is representative in geography, age, and gender.  Linkage was made at an individual level to COVID-19 PT-PCR testing data, hospital data, and regional mortality data.  General population: N= 5,586,521  Diagnosed with COVID-19: N= 102,002  Hospitalized with COVID-19: N= 16,901 | Median age  Whole study population  44 yrs., IQR: 25-60 yrs.  Diagnosed with COVID-19  47 yrs., IQR: 36-61  Hospitalized after diagnosis  61 yrs. (50-75 yrs.)  Died after being hospitalized  81 yrs. (73-87 yrs.)  Sex  Diagnosed with COVID-19  Female: 59 %  Male: 41%  Hospitalized after diagnosis  Female: 4,357 (46.2%)  Male: 5080 (53.8%) | March 1, 2020 to May 6, 2020 | 5-year age categories starting with 20 years to 95 years | Hospitalization with COVID-19 (population based)  Hospitalization with COVID-19 (diagnosed cases)  Mortality (from diagnosed to death/population based)  Mortality (hospital-based)  Outcome ascertainment through SIDIAP. |
| Carrillo-Vega*  2020  Mexico  (180) | Retrospective cohort | All records of positive cases of COVID-19 from the Epidemiological Surveillance System for Viral Respiratory Diseases of the Mexican Ministry of Health  N= 9946 | Mean age (SD)  48.15 yrs. (14.35)  Sex  Male: 5753 (57.8%)  Female: 4193 (42.2%) | Until April 23, 2020 | 25-49 yrs.  50-74 yrs.  ≥75 yrs. | Hospitalization by individuals testing positive (population-based)  Mortality (population-based)  Records from Mexican Ministry of Health |
| Carter and Collins**  2020  UK and Italy  (181) | Prospective cohort | Network of clinical teams with an interest in frailty from ten UK sites and one Italian site as part of the COVID-19 in Older People (COPE) study.  All consecutive patients admitted to hospitals aged ≥18 years with diagnosis of COVID-19. Diagnosis through swabs or by clinical diagnosis.  N= 1564  UK: 1410 (90.2%)  Italy: 154 (9.8%) | Median age: 74 yrs.  IQR: 61-83  Male: 903 (57.7%)  Female: 661 (42.3%) | Admitted between February 27- April 28, 2020 | <65 yrs.  65-79 yrs.  >80 yrs. | Mortality from the date of admission or date of diagnosis (in-hospital mortality)  Day 7 mortality (in-hospital)  Hospital records |
| Chen and Bai  2020  China  (184) | Retrospective cohort | Hospitalized patients in three branches of Tongji Hospital with confirmed COVID-19  N= 3309 | Median age: 62 yrs.  IQR: 49-69 yrs.  Male: 1642 (49.6%)  Female: 1667 (50.4%) | January 18- March 27, 2020 | ≤ 45 yrs.  > 45 yrs. | Mortality (in-hospital)  Hospital records |
| Cchiba and Patel‡  2020  USA  (186) | Retrospective cohort | Patients of all ages (two <18 years) with confirmed diagnosis of COVID-19 with RT-PCR in 10 hospitals affiliated with Northwestern Medicine (Chicago and surrounding Illinois suburbs)  N= 1526 | <40 yrs.: 414 (27.1%)  40-69 yrs.: 844 (55.3%)  ≥70 yrs.: 268 (17.6%)  Male: 718 (47%)  Female: 808 (53%) | March 1- April 15, 2020; mortality determined up to April 30, 2020 | <40 yrs.  40-69 yrs.  ≥70 yrs. | Hospitalization  Ascertainment by electronic records |
| Chilimuri  2020  USA  (187) | Retrospective cohort | Consecutive patients 18 years and above diagnosed with COVID-19 by RT-PCR test and admitted to the Bronxcare Health System (BCHS) hospital in the South Bronx, New York City.  N= 375 | Median age: 63 yrs.  IQR: 52-72  Male: 236 (63%)  Female: 139 (37%) | March 9 – April 9, 2020 | Per year | Mortality (in-hospital)  Hospital and electronic health records |
| Ciardullo  2020  Italy  (188) | Retrospective cohort | Patients aged 18 years or older admitted to Policlinico di Monza hospital in Monza, Italy diagnosed with COVID-19 through RT-PCR tests and/or clinically by the presence of typical symptoms, exposures to known affected individuals and radiographic findings of interstitial pneumonia.  N=371 (339 for analysis due to missing data) | Mean age: 72 yrs.  SD= 14  Male: 210 (65.4%)  Female: 129 (34.6%) | February 22-May 15, 2020 | Per year | Mortality (in-hospital)  Hospital and electronic records |
| Conversano and Melillo  2020  Italy  (190) | Retrospective cohort | All patients admitted to San Raffaele Hospital in Milan with a confirmed diagnosis of SARS-CoV-2 pneumonia by chest radiograph or computed tomographic scan and real-time polymerase chain reaction.  N=191 | Mean age: 63.4 yrs.  SD: 14.9  Male: 131 (68.6%)  Female: 60 (31.4%) | February 27- March 17, 2020 followed up until April 8, 2020 | Per year | Mortality (in-hospital)  Hospital electronic records |
| Costa Monteiro  2020  USA  (191) | Retrospective cohort | First 112 hospitalized patients 18 years or older with a positive PCR test or mini-bronchoalveolar lavage test at the University of California, Los Angeles (UCLA) hospital system (Ronald Reagan UCLA Medical Center and Santa Monica UCLA Medical Center  N=112 | Median age: 61 yrs.  IQR: 45-74  Male: 74 (66%)  Female: 38 (34%) | March 12- April 16, 2020 | Per year | Mechanical ventilation  Electronic health records/ hospital records |
| Cummings  2020  USA  (193) | Prospective cohort | Adult patients (aged 18 years or older) admitted to one of two New York-Presbyterian hospitals affiliated with Columbia University Irving Medical Center in northern Manhattan, New York City and diagnosed with laboratory-confirmed (RT-PCR) COVID-19 and were critically ill with acute hypoxaemic respiratory failure.  N= 257 | Median age: 62 yrs.  IQR: 51-72 | March 2- April 1, 2020 followed up until April 28, 2020 | Per 10-year increase | In-hospital mortality  among critically ill patients  Medical/hospital records |
| Di Castelnuovo  2020  Italy  (197) | Retrospective cohort | Hospitalized Patients in 30 Italian clinical centres.  Laboratory-confirmed SARS-CoV-2 infection confirmed by polymerase chain reaction on a nasopharyngeal swab.  N=3894 | Median age:  67 yrs.  IQR: 55-79 yrs.  Male: 2403 (61.7%)  Female: 1491 (38.3%) | February 19-May 23, 2020  assessed on May 29, 2020 | 18-44 yrs.  45-64 yrs.  65-74 yrs.  75-84 yrs.  85+ yrs. | In-hospital mortality or palliative discharge  Clinical data were abstracted from electronic medical records or charts |
| Docherty  2020  United Kingdom  (198) | Prospective cohort | Admitted patients to one of 208 acute care hospitals in England, Scotland, and Wales who tested positive for SARS-CoV-2.  RT-PCT was the only mode of testing available during the period of study. The decision to test was at the discretion of the clinician attending the patient, and not defined by protocol. Only patients who tested positive for Covid-19 were eligible for enrolment.  N= 20,133 (34% of covid-19 admissions in the 3 countries) | Median age:  73 yrs.  IQR: 58-82 yrs.  Range: 0-104 yrs.  Male: 12,068 (59.9%)  Female: 8065 (40.1%) | February 6 - April 19, 2020  followed up until May 3, 2020 | <50 yrs.  50-59 yrs.  60-69 yrs.  70-79 yrs.  ≥80 yrs. | In-hospital mortality  Hospital records |
| Du  2020  China  (199) | Prospective cohort | Hospitalized patients in Wuhan Pulmonary Hospital (Wuhan, Hubei Province, China).  All patients with confirmed or suspected COVID-19 pneumonia enrolled in this study were tested by RT-PCR test.  N=179 | Mean age:  53.0 yrs.  SD: ±13.7 yrs.  Median age:  57.6 yrs.  IQR: 49-68 yrs.  Range: 18-87 yrs.  Male: 97 (54.2%)  Female: 82 (45.8%) | December 25, 2019 - February 7, 2020  followed up until February 14, 2020 | <60 yrs.  60-69 yrs.  ≥70 yrs. | In-hospital risk of severity at admission  (critically ill patients having one of respiratory failure and requirement for mechanical ventilation, or shock or combined with other organ system failure which warrant ICU admission)  Data were collected from the electronic medical records using a piloted case report form from the time of hospital admission. |
| Ebinger  2020  USA  (201) | Retrospective cohort | Patients who had a laboratory-confirmed diagnosis of SARS-CoV-2 via RT-PCR while being evaluated or treated for signs or symptoms at the Cedars-Sinai Health System Los Angeles (California). It includes Cedars-Sinai Medical Center (CSMC), Marina Del Rey Hospital (MDRH), and affiliated clinics.  N=442 | Mean age:  52.72 yrs.  SD: 19.65 yrs.  Male: 256 (57.9%)  Female: 186 (42.1%) | March 8 - March 21, 2020 | Per 10 years | Illness severity in 3 categories:  Hospitalized/non-ICU, ICU/Non-intubated, ICU/intubated  Electronic health records |
| Giacomelli  2020  Italy  (209) | Prospective cohort | Hospitalized adult Covid-19 patients to Luigi Sacco Hospital in Milan (Italy).  All of the study patients had Covid-19 confirmed by a positive RT-PCR test on a nasopharyngeal swab.  N=233 | Median age:  61 yrs.  IQR: 50-72 yrs.  Male: 72 (30.9%)  Female: 161 (69.1%) | February 21 -March 19, 2020  followed up until April 20, 2020 | Per 10 years | In-hospital mortality  Data were extracted from the patients' clinical charts on a daily basis.  The life status of the patients discharged before the censoring date was ascertained by means of telephone calls made by two physicians on 20 April, 2020. |
| Giannouchos*  2020  Mexico  (210) | Retrospective cohort | Patients initially classified as “suspected cases of viral respiratory disease” during point of service at medical facilities in Mexico, from a dataset released by the Mexican Health Ministry and compiled by the General Bureau of Epidemiology through the System of Epidemiological Surveillance of Viral Respiratory Diseases (comprising of 475 monitoring units).  Patients included had laboratory-confirmed diagnosis by RT-PCR if they had serious symptoms.  N=89,756 | Mean age:  46.2 yrs.  SD: 16.0 yrs.  Male: 56.4%  Female: 43.6% | Dataset released on May 31, 2020 | 0-17 yrs.  18-44 yrs.  45-64 yrs.  ≥65 yrs. | Population-based hospitalization,  Electronic/medical records |
| Grasselli  2020  Italy  (213) | Retrospective cohort | All 4209 consecutive patients with confirmed SARS-CoV-2 infection were admitted to one of the COVID-19 Lombardy Network’s ICUs. All the critically ill patients requiring ICU admission in Lombardy have been referred to the Regional Coordinating Center.  Laboratory-confirmed diagnosis of SARS-CoV-2 infection by RT-PCR test.  N = 3988 | Median age:  63 yrs.  IQR: 56-69 yrs.  Male: 3188 (79.9%)  Female: 800 (20.1%) | February 20 - April 22, 2020  followed up until May 30, 2020 | Per 10 years | In-hospital mortality of ICU patients  The staff of the Regional Coordinating Center contacted  each ICU of the Network daily by telephone and recorded on  an electronic worksheet the demographic and clinical patient data. |
| Gu  2020  China  (214) | Nested case-control | Source data from 12,973 publicly reported laboratory-confirmed patients with Covid-19 in 31 province-level regions outside of Hubei Province in mainland China by the National Health Committee during the study period. Deaths included as cases and each case was matched with up to three controls on gender and age +/- 1 year old. Asymptomatic people not included.  N=275 | Median age:  68.0 yrs.  IQR: 22 yrs.  Male: 173 (62.9%)  Female: 102 (37.1%) | December 18, 2019 - March 8, 2020 | Per year | Population-based mortality  National/provincial/municipal health commission websites, the official COVID-19 data reporting websites in China. |
| Gu  2020  USA  (215) | Retrospective cohort | Patients who were tested at University of Michigan Health System (MM), transfer patients from other hospitals, and patients who were tested elsewhere but treated at MM.  Diagnostic tests: RT-PCR (88.6%), commercial antibody test, COVID-19 nasopharynx or oropharynx PCR tests. Prioritized testing  N=1139 (Positive tests)  N= 523 (hospitalized) | Median age:  53 yrs.  IQR: 39-66 yrs.  Male: 531 (46.6%)  Female: 608 (53.4%) | March 10 - April 22, 2020  followed up until July 28, 2020 | Per 10 yrs.  <18 yrs.  18- <35 yrs.  35- <50 yrs.  50- <65 yrs.  65- <80 yrs.  ≥80 yrs. | Hospitalization  ICU (population-based)  Mortality (population-based)  Electronic health records |
| Gupta  2020  USA  (216) | Prospective cohort | Adult patients (≥18 years of age) with Covid-19 to participating admited to ICUs in 65 hospitals in the USA.  Laboratory-confirmed diagnosis by nasopharyngeal or oropharyngeal swab.  N= 2215 | Mean age:  60.5 yrs.  SD: 14.5 yrs.  Male: 1436 (64.8%)  Female: 779 (35.2%) | March 4 - April 4, 2020  followed up until June 4, 2020 | 18-39 yrs.  40-49 yrs.  50-59 yrs.  60-69 yrs.  70-79 yrs.  ≥80 yrs. | 28-day in-hospital mortality for ICU patients  Mechanical ventilation for ICU patients  Electronic medical records. |
| Harrison  2020  USA  (219) | Retrospective cohort | Patients 18 years or older with Covid-19 from 24 participating healthcare organizations (inpatient and outpatient care settings), distributed between the four large Census Bureau–designated regions of the US (Northeast/Midwest/South/West).  Identified based on a positive laboratory test (not specified) in their records and ICD-classification based on Centers for Disease Control and Prevention (CDC) coding guidelines. False positives excluded.  N=31,461 | Median age:  50 yrs.  IQR: 35-63 yrs.  Male: 14,306 (45.5%)  Female: 17,155 (54.5%) | January 20 - May 26, 2020 | Per year | Population-based mortality  Electronic medical records |
| Hashemi  2020  USA  (220) | Retrospective cohort | Adult patients hospitalized to nine hospitals (two large tertiary centres and seven community  hospitals) in a single healthcare system in Massachusetts.  Laboratory-confirmed diagnosis of SARS-CoV-2 infection via polymerase chain reaction (PCR) nasopharyngeal swab or tracheal aspirate.  N=363 | Mean age:63.4 yrs.  SD: 16.5 yrs.  Male: 201 (55.4%)  Female: 162 (44.6%) | March 11 – April 2, 2020 | Per year | In-hospital mechanical ventilation  In-hospital ICU admission  In-hospital mortality  Electronic medical records. |
| Hewitt**  2020  United Kingdom and Italy  (221) | Prospective cohort | Adult patients (≥18 years) admitted with Covid-19 to ten participating hospitals in the UK and one in Italy, based on the COVID-19 in Older People (COPE) study.  Inclusion criteria were complete hospital records and laboratory-confirmed SARS-CoV-2-positive swabs or a clinical diagnosis made by the parent clinical team and based on signs, symptoms, or radiology consistent with Covid-19.  N=1564 | Median age:  74 yrs.  IQR: 61-83 yrs.  Male: 903 (57.7%)  Female: 661 (42.3%) | February 27 - April 28, 2020 | <65 yrs.  65-79 yrs.  ≥80 yrs. | In-hospital mortality  Day 7 ortality  Hospital/medical records |
| Hwang  2020  South Korea  (224) | Retrospective cohort | Included two cohorts of adult inpatients (≥ 18 years old) with near severe stage of Covid-19 from Kyungpook National University Hospital and Kyungpook National University Chilgok Hospital in Daegu.  Confirmed diagnosis of SARS-CoV-2 infection by RT-PCR  N=103 | Mean age:  67.62 yrs.  SD: 15.32 yrs.  Range: 24-97 yrs.  Male: 52 (50%)  Female: 51 (50%) | February 1 - March 25, 2020 | Per year | In-hospital mortality (among moderate/severely diseased)  Electronic medical records. |
| Imam  2020  USA  (225) | Retrospective cohort | Hospitalized patients in eight hospitals of Beaumont Health, the largest healthcare system in  Southeast Michigan caring for over a third of patients in Detroit.  Laboratory-confirmed diagnosis of SARS-CoV-2 infection by using RT-PCR on nasopharyngeal swab.  N=1305 | Mean age:  61 yrs.  SD: 16.3 yrs.  Range: 24-97 yrs.  Male: 702 (53.8%)  Female: 603 (46.2%) | March 1 - April 1, 2020  Assessed on April 17, 2020 | >60 yrs. vs.  <60 yrs. | In-hospital mortality  Electronic medical records. Manual chart review toconfirm mortality to ensure accuracy and completeness. |
| Jun  2020  USA  (226) | Prospective cohort | Admitted patients with Covid-19 to five Mount Sinai hospitals in New York City: Three in Manhattan; 1 in Brooklyn; 1in Queens, through the emergency department  Nucleic acid-based test to detect SARS-CoV-2 infection in nasopharyngeal or oropharyngeal swab specimens.  N=3086 | Median age:  66 yrs.  IQR: 56-77 yrs.  Male: 1825 (59.1%)  Female: 1261 (40.9%) | Admitted on or before April 13, 2020  Followed through June 2, 2020 | Per year | In-hospital mortality  Electronic health records |
| Kabarriti  2020  USA  (227) | Retrospective cohort | Patients with Covid-19 who received care at the Montefiore Medical Center, a large academic medical center located in the Bronx (New York), whether or not they were admitted as patients.  SARS-CoV-2 positive status was determined based on reverse transcription quantitative polymerase chain reaction assay.  N=5902 | Median age:  58 yrs.  IQR: 44-71 yrs.  Male: 2773 (47%)  Female: 3129 (53%) | March 14 - April 15, 2020  final data collection on April 27, 2020 | ≤ 40 yrs.  41-60 yrs.  61-80 yrs.  >80 yrs. | Mortality (population-based)  Data were obtained from electronic medical records. |
| Kalligeros  2020  USA  (228) | Retrospective cohort | Patients 18 years or older admitted with Covid-19 to Rhode Island Hospital, The Miriam Hospital, or Newport Hospital, in Rhode Island (USA).  Laboratory-confirmed diagnosis of SARS-CoV-2 infection by using a reverse transcriptase-polymerase chain reaction assay.  N=103 | Median age:  60 yrs.  IQR: 52-70 yrs.  Male: 63 (61.17%)  Female: 67 (38.83%) | February 17 - April 5, 2020 | Per year | ICU admission within 10 days of hospital admission  Mechanical ventilation within 10 days of hospital admission  Electronic chart review |
| Khalil  2020  UK  (229) | Prospective cohort | Adult patients (≥18 years old) with laboratory-confirmed RT-PCR test COVID-19 admitted to Chelsea & Westminster Hospital, a central London teaching hospital.  N= 220 | Mean age:  66.9 years  95% CI: 64.7–69.2 years  Male: 130 (59.1%)  Female: 90 (40.9%) | March 7 -April 7, 2020  Followed up until May 8, 2020 | Per year | In-hospital mortality at 28 days  Medical records, death certificates |
| Killerby  2020  USA  (230) | Retrospective cohort | Hospitalized patients selected sequentially from hospital-provided lists of patients aged ≥18 years with laboratory (RT-PCR) confirmed COVID-19  and non-hospitalized patients aged ≥18 years with laboratory (RT-PCR) confirmed COVID-19-evaluated at outpatient clinics or an emergency department and not admitted. Data from six acute care hospitals and associated outpatient clinics in metropolitan Atlanta, Georgia.  N=531 (220 hospitalized, 311 non-hospitalized) | Non-hospitalized  Mean age: 45 years  IQR: 33.0-58.0 years  Hospitalized  Mean age: 61 years  IQR: 45.0-70.0 years  Non-hospitalized  Male: 114 (36.7%)  Female: 197 (63.3%)  Hospitalized  Male: 114 (51.8%)  Female: 106 (48.2%) | Hospitalized:  March 1 – March 30, 2020  Non-hospitalized:  April 8-May 1, 2020 | 18-44 yrs.  45-64 yrs.  ≤65 yrs. | Hospitalization (included stays for observation and deaths that occurred in an emergency department (ED))  Medical records |
| Kim  2020  USA  (231) | Retrospective cohort | Patients 18 years or older hospitalized with laboratory-confirmed COVID-19  Identified through the Coronavirus Disease 2019-Associated Hospitalization Surveillance Network (COVID-NET) covering 154 acute-care hospital in 74 counties and 13 states in the USA and who had complete chart review.  N= 2491 | Median age: 62 yrs.  IQR: 50-75 yrs.  Male: 1326 (53.2%)  Female: 1165 (46.8%) | March 1 – May 2, 2020  Assessed on May 2, 2020 | 18-39 yrs.  40-49 yrs.  50-64 yrs.  65-74 yrs.  75-84 yrs.  ≥85 yrs. | Intensive care unit (ICU)  In-hospital mortality  COVID-NET surveillance |
| Klang  2020  USA  (233) | Retrospective cohort | Hospitalized adult patients in a large academic hospital system in New York City (Mount Sinai, 5 hospitals) with positive RT-PCR test for COVID-19  and had definite outcomes (discharged or died). Patients with missing BMI data were excluded.  N= 3406 | Age ≤50 years: 572 (16.8%)  Age >50 years: 2834 (83.2%)  Age ≤50 years  Male: 397 (69.4%)  Female: 175 (30.6%)  Age >50 years  Male: 1564 (55.1%)  Female: 1270 (44.9%) | March 1- May 17, 2020 | Per decile, stratified by ≤50 years and  >50 years | In-hospital mortality  Electronic medical records |
| Lee  2020  South Korea  (235) | Retrospective cohort | Patients hospitalized or isolated with laboratory-confirmed SARS-CoV2-infection via RT-PCR  Data from Korea Centers for Disease Control and Prevention (KCDC)  N= 8,266 | Mean age: 44.36 years  SD: 19.13 years  Male: 3181 (39.48%)  Female: 5085 (61.52 %) | January 19 -March 16, 2020 assessed on March 24, 2020 | Per year | Mortality within 60 days (hospitalized or isolated)  Electronic health records from the National Health Information Databas |
| Li  2020  China  (238) | Retrospective cohort | Patients with laboratory-confirmed SARS-CoV-2 infection via RT-PCR at Wuhan Seventh People’s Hospital, Wuhan, China.  N= 596 | Median age: 58 years  IQR: 47–68 years  Male: 280 (47.0 %)  Female: 316 (53.0 %) | January 23 - March 14, 2020 | <65 years  ≥ 65 years | In-hospital mortality  Electronic health records |
| Magleby  2020  USA  (241) | Retrospective cohort | Patients with coronavirus disease confirmed via RT-PCR tests hospitalized at two hospitals in New York City (New York Presbyterian Hospital/Weill Cornell Medical Center and affiliated Lower Manhattan Hospital).  N= 678 | Median age (IQR)  high viral load:  72 yrs. (60-81 yrs.)  medium viral load:  69 yrs. (58-79 yrs.)  low viral load:  63 yrs. (50-73 yrs.)  high viral load  Male: 139 (63.2%)  Female: 81 (36.8%)  Medium viral load  Male: 132 (61.1%)  Female: 84 (38.9%)  Low viral load  Male: 143 (59.1%)  Female: 99 (40.9%) | March 30 - April 30, 2020 | Per year | In-hospital mortality  Electronic medical records |
| Meng  2020  China  (248) | Retrospective cohort‡ | Patients with pathogen-confirmed COVID-19 via RT-PCR who were hospitalized at Tongji Hospital in Wuhan, China diagnosed with moderate to severe pathogen-confirmed COVID-19  N= 2556 | Age^1^:  ≤49 years: 707 (26.53%)  50-64 years: 907 (34.93%)  65-79 years: 868 (32.57%)  ≥80 years: 185 (06.94%)  Male: 1328 (49.83%)  Female: 1337 (50.17%) | January 18 - March 27, 2020 | ≤49 Years  50-64 years  65-79 years  ≥80 years | In-hospital mortality  Medical records |
| Merzon  2020  Israel  (249) | Retrospective cohort | Population-based epidemiological study utilizing data from the Leumit Health Services (LHS) database. The study population included the 14,000 members of LHS tested for COVID-19 infection by PCR test.  N= 782 people with positive PCR test | Mean age: 35.58 years  (95% CI: 34.49-36.67)  Male: 385 (49.23%)  Female: 397 (50.77%) | February 1 - April 30, 2020 | ≤50 years  >50 years | Hospitalization |
| Murillo-Zamora and Hernandez-Suarez††  2020  Mexico  (255) | Retrospective cohort | Hospitalized adult patients (18 and older) with a laboratory-confirmed case of COVID-19 within 14 days before symptoms onset identified from epidemiological surveillance of viral respiratory diseases, belonging to the Mexican Institute of Social Security (IMSS). IMSS provides health care services to more than 1/3 of population of Mexico.  N= 66,123 | Age (years)  20-29: 2264 (3.4%)  30-34: 10,616 (16.1%)  45-59: 22,415 (33.9%)  60+: 30,828 (46.6%)  Male: 40,124 (60.7%)  Female: 25,999 (39.3%) | March 4-August 15, 2020 | 20-29 yrs.  30-34 yrs.  45-59 yrs.  60+ yrs. | In-hospital mortality  Medical records and death certificates |
| Narain  2020  USA  (257) | Retrospective cohort | Patients admitted to the 12 hospitals and emergency departments within the Northwell Health system, the largest private nonprofit health system in New York State, who were diagnosed with COVID-19 by RT-PCR tests and who were older than 18 years, and meeting COVID-19 cytokine storm (CCS) criteria.  N= 5,776 | For standard of care group:  Median age: 64.6 yrs.  IQR: 53.5-76.4  Male: 3702 (64.1%)  Female: 2075 (35.9%) | March 1- April 24, 2020 | Per year | In-hospital mortality  among CCS patients  Electronic health records |
| Palaiodimos  2020  USA  (259) | Retrospective cohort | First 200 patients who presented to the emergency room and were admitted to the inpatient medicine service or the intensive care unit at the Montefiore Medical Center in the Bronx, New York with laboratory-confirmed COVID-19.  N= 200 | Median age: 64 years  IQR: 50-73.5  Male: 98 (49%)  Female: 102 (51%) | Admission between March 09- March 22, 2020 followed up until April 12, 2020 | Per year and per quartiles (results shown for per year only) | In-hospital mortality  Medical records |
| Patel‡  2020  USA  (261) | Retrospective cohort | Adult patients 18 years or older hospitalized with COVID-19 in one of ten hospitals affiliated with Northwestern Medicine in Chicagoland, with or without anticoagulation treatment. They were discharged or died during study period.  N=1716 | Age distribution  18-44 yrs.: 22.1%  45- 59 yrs.: 29.6%  60-69 yrs.: 21.6%  70-79 yrs.: 14.1%  >80 yrs.: 12.6%  Male: 935 (54.5%)  Female: 781 (45.5%) | March 9- June 26, 2020 | 18-44 yrs.  45- 59 yrs.  60-69 yrs.  70-79 yrs.  >80 yrs. | In-hospital mortality  Mechanical ventilation  Death in critical illness  Data extracted from Northwestern University Enterprise Data Warehouse (NUEDW) (medical records) |
| Perez-Guzman  2020  UK  (262) | Retrospective cohort | All admission with RT-PCR-positive SARS-CoV-2 infection to the Imperial College Healthcare National Health Service Trust (ICHNT), one of the largest hospital trusts in England serving North West London.  N= 559 with completed outcomes (death or discharge) until May 1, 2020 | Median age: 69 years  IQR: 54-79 years  Male: 382 (62%)  Female: 232 (38%) | February 25- April 5, 2020 followed up until May 1, 2020 | Per year | In-hospital mortality  Hospital records |
| Petrilli  2020  USA  (263) | Prospective cohort | Patients with all data available (besides age and sex) with positive RT-PCT test for SARS-CoV-2, containing four acute care hospitals (two in Manhattan, one in Brooklyn, and one in Long Island).  N= 5279 (positive test)  N= 2741 (hospitalized) | Median age: 54 years  IQR: 38-66 years  Men: 2615 (49.5%)  Women: 2674 (50.5%) | March 1-April 8, 2020, followed up until May 5, 2020 | 19-44 yrs.  45-54 yrs.  55-64 yrs.  65-74 yrs.  ≥75 yrs. | Hospital admission  In-hospital mortality (discharge to hospice or death)  Electronic health records. |
| Price-Haywood  2020  USA  (266) | Retrospective cohort | Patients seen at an Ochsner Health facility in Louisiana, who tested positive for SARS-CoV-2 on PCR assay and identified themselves as white non-hispanic or as black non-hispanic.  Excluded were those with any other ethnicity.  N= 3481 (for hospitalization)  N = 1382 (for in-hospital mortality) | Mean age (SD)  White non-hispanic  55.5 yrs (18.5)  Black non-hispanic  53.6 yrs. (16.1)  White non-hispanic  Male: 471 (45.7%)  Female: 559 (54.3%)  Black non-hispanic  Male: 923 (37.7%)  Female: 1528 (62.3%) | March 1- April 11, 2020 followed through May 7, 2020 | Age, per 5-year units | Hospitalization  ICU  In-hospital mortality  Electronic medical record system. |
| Reilev  2020  Denmark  (269) | Retrospective cohort | People in the Danish population who tested positive for SARS-CoV-2 via PCR tests, whose results were in the Danish Microbiology Database. A unique personal identifier assigned to all Danish citizens was used to link to health registries for information.  N= 11,122 (positive cases, for hospitalization, death within 30 days)  N= 2254 hospitalized (for ICU, in-hospital mortality) | Positive cases:  Median age: 48 yrs.  IQR: 33-62 yrs.  Men: 4692 (42%)  Women: 6430 (58%) | February 27- May 19, 2020 | 0-9 yrs.  10-19 yrs.  20-29 yrs.  30-39 yrs.  40-49 yrs.  50-59 yrs.  60-69 yrs.  70-79 yrs.  80-89 yrs.  90+ yrs. | Hospitalization  Mortality (population-based)  In-hospital mortality  Danish administrative and health registries. |
| Rentsch  2020  USA  (270) | Retrospective cohort | Members of the VA Birth Cohort aged 54 to 75 years with a laboratory confirmed positive SARS-CoV-2/Covid-19 nasopharyngeal swab (1% were from other sources) result.  N=3789  N=585 (positive cases, for hospitalization and ICU) | Median age: 66.1 yrs.  IQR: 60.4-71 yrs.  Male: 27 (4.6 %)  Female: 558 (95.4 %) | February 8 - March 30, 2020 | Age, per 5-year increase | Hospitalization  ICU  Electronic health record data  from Veterans Affairs Healthcare System/Veterans Health Administration |
| Rossi  2020  Italy  (274) | Retrospective cohort | All symptomatic patients who tested positive with PCR in the province of Reggio Emilia registered in a special dedicated SARS-CoV-2 database. Asymptomatic cases excluded.  N= 2653 | Median age: 63.2 yrs.  Male: 1328 (50,1 %)  Female: 1325 (49,9 %) | February 27-April 2, 2020- assessed until April 3, 2020. | <51 yrs.  51-60 yrs.  61-70 yrs.  71-80 yrs.  ≥81 yrs. | Hospitalization  Death (population-based)  Data obtained through routinely available administrative databases of the Health Authority |
| Salacup  2020  USA  (277) | Retrospective cohort | All patients older than 18 years old with a confirmed diagnosis of COVID-19 via RT-PCR diagnosis presenting at the Einstein Medical Center in Philadelphia. Included data for patients with definite outcome (discharge or death)  N= 242 | Median age: 66 yrs.  IQR: 58-76 yrs.  Male: 93 (51%)  Female: 119 (49%) | March 1- April 24, 2020 | Per year | In-hospital mortality |
| Sapey  2020  England  (279) | Retrospective cohort | All patients with a confirmed positive SARS-CoV-2 swab (nasopharyngeal and oropharyngeal) result admitted to the University Hospitals Birmingham NHS Foundation Trust at the time or up to 2 weeks following their first positive test. Not specified if lateral flow test or PCR test.  N=2169 | Median age: 73 yrs.  IQR: 58-84 yrs.  Male: 1290 (58.2%)  Female: 927 (41.8%) | March 10- April 17, 2020 assessed on May 12, 2020 | z-score | In-hospital mortality  Electronic health records |
| Seiglie  2020  USA  (280) | Prospective cohort | Patients presenting to care with COVID-19 symptoms and who were subsequently hospitalized at Massachusetts General Hospital (MGH) with PCR confirmed SARS-CoV-2 infection.  N=450 | Mean age (SD)  No diabetes: 61.1 yrs. (18.8)  Diabetes: 66.7 yrs. (14.2)  Male: 259 (57.6%)  Female: 191 (42.4%) | March 11- April 30, 2020 followed up for 14 days from date of initial presentation | <50 yrs.  50-59 yrs.  60-69 yrs.  ≥70 yrs. | 14-day ICU admission  14-day mechanical ventilation  14-day in-hospital mortality  COVID-19 data registry |
| Shah  2020  USA  (281) | Retrospective cohort | All hospitalized patients in Phoebe Putney Health System in Southwest Georgia, with confirmed COVID-19 through a PCR test and a definite outcome  N= 522 | Median age: 63 yrs.  IQR: 50-72 yrs.  Male: 218 (41.8%)  Female: 304 (58.2%) | March 2- May 6, 2020  Assessed on May 6, 2020 | <65 yrs.  ≥65 yrs. | In-hospital mortality  Electronic medical records |
| Shi†  2020  China  (285) | Retrospective cohort study | Consecutive patients admitted to Renmin Hospital of Wuhan University in Wuhan with laboratory-confirmed COVID-19 using RT-PCR tests. Patients were excluded if they did not have cardiac biomarkers (N= 229 excluded).  N= 416 | Median age: 64 yrs.  Range: 21-95 yrs,  Male: 205 (49.3%)  Female: 211 (50.7%) | January 20- February 10, 2020  followed up February 15 2020 (mortality) | Per year | In-hospital mortality  Electronic medical records |
| Soares  2020  Brazil  (293) | Retrospective cohort | All patients diagnosed with a RT-PCR or serological test, or if they were a suspected case that previously came into close contact or resided with someone with a confirmed laboratory diagnosis in the state of Espíritu Santo, with complete data for predictive variables.  N= 10,713 | Age distribution  <60 yrs.: 8676 (81.0%)  ≥60 yrs. 2037 (19.0%) | February 29-June 11, 2020  Assessed on June 11, 2020 | <60 yrs.  ≥60 yrs. | Hospitalization due to COVID-19  Health Secretariat of Espíritu Santo state |
| Solís  2020  Mexico  (294) | Retrospective cohort | 7497 patients who tested positive for COVID-19 in Mexico, a subset of a larger dataset of 49167 patients with COVID-19 related symptoms who sought attention in medical units and were tested with positive, negative and pending results.  N=7497 | Median age: 46 yrs.  Male: 4341 (57.9%)  Female: 3156 (42.1%) | Until April 18, 2020 | 0-24 yrs.  25-29 yrs.  30-34 yrs.  35-39 yrs.  40-44 yrs.  45-49 yrs.  50-54 yrs.  55-59 yrs.  60-64 yrs.  65-69 yrs.  70+ yrs. | Population-based mortality at 35 days  Information about patients is reported on a daily basis to the Secretaría de Salud and released by the Mexican Ministry of Health. |
| Sousa  2020  Brazil  (295) | Retrospective cohort | People that presented flu-like syndrome in Fortaleza (Ceará’s capital city) and tested positive for Covid-19. Only people that needed medical assistance and went to healthcare services were tested; from those, moderate and severe cases were hospitalized and received health assistance.  N=2070 | Median age: 44 yrs.  IQR: 34-59 yrs.  Male: 1017 (49.1%)  Female: 1053 (50.9%) | Until April 14, 2020 (endpoint) | <60 yrs.  ≥60 yrs. | Population-based mortality  Data were obtained by using IntegraSUS data, a free-access website that holds daily  updated data and indicators of Covid-19 in the Ceará State.  The cause of death was determined by the physician given positive laboratory tests to COVID-19 and the clinical manifestation of the disease. |
| Suleyman  2020  USA  (296) | Retrospective cohort | Adult patients hospitalized at Henry Ford Health System (HFHS), a 5-hospital system that serves metropolitan Detroit, and diagnosed with COVID-19.  Laboratory-confirmed SARS-CoV-2 infection by PCR test.  N=355 hospital admission | Mean age (SD):  61.4 yrs. (15.4 yrs.)  Male: 165 (46.5%)  Female: 190 (53.5%) | March 9 - March 27, 2020, clinical outcomes monitored for 30 days. | <60 yrs.  >60 yrs. | ICU  Mechanical ventilation  Data were obtained from electronic health records on a standardized data collection form. |
| Tai  2020  China  (299) | Retrospective cohort | Admitted patients to Wuchang temporary hospital with laboratory-confirmed COVID-19, but having mild COVID-19. Critical patients were transferred to other hospitals for ICU care.  (RT-PCR) test.  N= 332 | Median age: 51 yrs.  IQR: 40-59 yrs.  Male: 132 (39.8%)  Female: 200 (61.2%) | February 5 - March 10, 2020  followed until March 25, 2020 | Per year | ICU (for mild COVID-19)  Electronic medical records. |
| Tartof  2020  USA  (300) | Retrospective cohort | All Kaiser Permanente Southern California (KPSC) members with 6-month continuous membership and diagnosed with COVID-19 by  diagnostic codes or positive laboratory test results (82%). Pregnant women were excluded.  ):KPSC is anintegrated health care organization located throughout 9 counties in Southern California with more than 4.7 million members.  N=6916 | Mean age (SD):  49.1 yrs. (16.6 yrs.)  Median age: 49 yrs.  IQR: 36-60 yrs.  Male: 3111 (44.98%)  Female: 3805 (55.02%) | February 13 - May 2, 2020, last date patient was enrolled was set to 21 days before study date end | 0-40 yrs.  41-50 yrs.  51-60 yrs.  61-70 yrs.  71-80 yrs.  >80 yrs. | Population-based mortality (within 21 days)  Electronic health records. |
| van Gerwen  2020  USA  (301) | Retrospective cohort | All patients 18 years or older with laboratory-confirmed diagnosis (hospitalized and ambulatory) identified via a large New York City health system with sufficient clinical documentation available or accessible, including confidential patient records.  Laboratory‐confirmed diagnosis of COVID‐19 by a RT-PCR test  N=3703 (all patients)  N= 2015 (hospitalized) | Mean age (SD):  56.8 yrs. (18.2 yrs.)  Male: 2049 (55.3%)  Female: 1654 (44.7%) | March 1 2020 – April 1, 2020 followed until May 13, 2020 (6 weeks) | 18-40 yrs.  40-60 yrs.  >60 yrs. | Hospitalization  Mechanical ventilation  In-hospital mortality  Electronic medical records. |
| Wang  2020  USA  (302) | Retrospective cohort | All confirmed COVID-19 patients within the Mount Sinai Health System (MSHS) in New York City. MSHS is the largest integrated healthcare delivery system in the New York City metropolitan region serving 3499000 million outpatients and 152520 inpatients annually.  Confirmed diagnosis of SARS-CoV-2 by lab tests (not specified).  N=7592 | Age, without comorbidity N (%):  <18 yrs: 39 (76%)  18-39 yrs: 1315 (85%)  40-49 yrs: 709 (75%)  50-59 yrs: 841 (64%)  60-69 yrs: 830 (52%)  70-79 yrs: 510 (45%)  80+ yrs: 472 (46%)  Age, with comorbidity N (%):  <18 yrs: 12 (24%)  18-39 yrs: 224 (15%)  40-49 yrs: 234 (25%)  50-59 yrs: 465 (36%)  60-69 yrs: 753 (48%)  70-79 yrs: 627 (55%)  80+ yrs: 564 (54%)  Male: 4165 (54.9%)  Female: 3427 (45.1%) | Until April 15, 2020 | <40 yrs.  40-49 yrs.  50-59 yrs.  60-69 yrs.  70-79 yrs.  80+ yrs. | Population-based mortality  Electronic health records. |
| Wang  2020  China  (305) | Retrospective cohort | All confirmed cases of COVID-19 over 60 years old admitted at isolation ward of Renmin Hospital of Wuhan University.  Laboratory‐confirmed SARS-CoV-2 nucleic acid by  RT-PCR test.  N=339 | Median age: 69 yrs.  IQR: 65-76 yrs.  Male: 166 (49%)  Female: 173 (51%) | January 1 – February 6, 2020  followed until March 5, 2020 | Per year | In-hospital mortality in patients over 60 yrs. old  Data were obtained from patient`s medical records. |
| Yehia  2020  USA  (312) | Retrospective cohort | Adults (18 years or older) with laboratory-confirmed SARS-CoV-2 infection by PCR hospitalized at one of 92 Ascension hospitals (located in 12 states).  N=7139 | Median age: 68 yrs.  IQR: 56-79 yrs.  Male: 3664 (51.3%)  Female: 3470 (48.6%) | February 19 - May 31, 2020  followed until June 25, 2020 | 18-49 yrs.  50-64 yrs.  65-84 yrs.  ≥85 yrs. | In-hospital mortality  Electronic health records and administrative data. |
| Zhao†  2020  China  (319) | Retrospective cohort | COVID-19 patients admitted to both the Shouyi and East districts of Renmin Hospital of Wuhan University.  Laboratory-confirmed diagnosis of SARS-CoV-2 by positive nucleic acid test and sequencing of viral genes, highly homologous to known SARS-CoV-2.  N=1000 | Median age: 61 yrs.  IQR: 46-70 yrs.  Male: 466 (46.6%)  Female: 534 (53.4%) | January 1 - February 14, 2020 | <60 yrs.  60-75 yrs.  ≥75 yrs. | In-hospital mortality  Hospital's medical record system. |
| †, ††, ‡, *, ** Studies marked with same symbol have the same population or a sub-group of the same population  IQR: interquartile range; yrs.: years; reverse-transcriptase polymerase chain reaction: RT-PCR; ICU: intensive care unit; COPD: chronic obstructive pulmonary disease; BMI: body mass index; yrs.: years; SOFA score: sequential organ failure assessment score; OR: odds ratio; HR: hazards ratio; CI: confidence intervals; NA: not available; IQR: inter-quartile range | | | | | | |

**Table S3. Results of studies investigating in-hospital mortality**

| **Author,**  **Year**  **(Ref)** | **Confounders and**  **age-dependent risk factors used in the model** | **Type of analysis**  **# cases / # non-cases** | **N in analysis**  **Total deaths**  **In age group: # cases/total** | **Results**  **Unadjusted effect estimates (if available)** | **Results**  **Adjusted effect estimates** |
| --- | --- | --- | --- | --- | --- |
| Antwi-Amoabeng  2020  (165) | Sex, ethnicity (Hispanic), diabetes, hypertension, obesity, chronic kidney disease, COPD, ICU stay | Logistic regression | N= 172  Deaths: 18 (10.5%)  Age ≤61 yrs.: NA  Age>61 yrs.: NA | NA | Mortality (in hospital) OR (95% CI)  Age ≤61 yrs.: Reference  Age>61 yrs.: 5.63 (0.41-77.51) |
| Argenziano  2020  (166) | Sex, BMI, smoking, coronary artery disease, congestive heart failure, history of stroke, diabetes mellitus, hypertension, cirrhosis, HIV, inflammatory bowel disease, pulmonary disease, renal disease, viral hepatitis, active malignancy, transplant history, rheumatological disease, immunosuppressed state, no comorbidities | Cox proportional hazards analysis only with complete data (n=841) | N= 1000  Deaths: 211 (21%) | NA | In-hospital mortality HR (95% CI)  Age (per year): 1.07 (1.05-1.08) |
| Baqui  2020  (168) | Sex, ethnicity (black, Pardo, East Asian), neurological disease, obesity, pulmonary disease, renal disease, diabetes, immunosuppression, liver disease, cardiovascular disease, asthma | Mixed-effects Cox regression | N= 6882  Deaths: 3254  <40 yrs.: NA  40-50 yrs.: NA  50-60 yrs.: NA  60-70 yrs.: NA  ≥70 yrs.: NA | NA | In-hospital mortality HR (95% CI)  <40 yrs.: Reference  40-50 yrs.: 1.19 (1.00-1.43)  50-60 yrs.: 1.59 (1.34-1.87)  60-70 yrs.: 2.06 (1.76-2.41)  ≥70 yrs.: 3.02 (2.59-3.52) |
| Boulle  2020  (176) | Sex, diabetes, hypertension, chronic kidney disease, chronic pulmonary disease/asthma, tuberculosis, HIV, antiretroviral therapy, CD4 count during COVID-19 | Cox proportional hazards model | Hospitalized public-sector SARS-CoV-2 cases  N= 2,978  Deaths: 550  20-39 yrs.: 45/849  40-49 yrs.: 56/528  50-59 yrs.: 142/665  60-69 yrs.: 158/518  ≥70 yrs.: 149/418 | NA | In-hospital mortality HR (95% CI)  Hospitalized public-sector SARS-CoV-2 cases (in-hospital mortality)  20-39 yrs.: Reference  40-49 yrs.: 1.83 (1.23-2.72)  50-59 yrs.: 3.81 (2.68-5.42)  60-69 yrs.: 6.11 (4.27-8.75)  ≥70 yrs.: 7.53 (5.23-10.84) |
| Burn  2020  (177) | Autoimmune condition, chronic kidney disease, COPD, dementia, heart disease, hyperlipidemia, hypertension, malignant neoplasm, obesity, Type 2 diabetes, Charlson Index | Cox proportional hazards model using a non-linear relationship with age | N= 16,901  Deaths: 2692  >18 yrs.: <5/74  18–39 yrs.: 11/1276  40-59 yrs.: 142/5217  60-69 yrs.: 292/3231  70-79 yrs.: 822/3764  ≥80 yrs.: 1424/3339 | In-hospital mortality HR (95% CIs)  Age only model:  20-24 yrs.: 0.052 (0.02 to 0.14)  25-29 yrs.: 0.066 (0.03 to 0.15)  30-34 yrs.: 0.084 (0.045 to 0.16)  35-39 yrs.: 0.11 (0.068 to 0.17)  40-44 yrs.: 0.14 (0.099 to 0.19)  45-49 yrs.: 0.18 (0.14 to 0.23)  50-54 yrs.: 0.25 (0.20 to 0.31)  55-59 yrs.: 0.37 (0.30 to 0.44)  60-64 yrs.: 0.59 (0.54 to 0.65)  65-69 yrs.: Reference  70-74 yrs.: 1.69 (1.61 to 1.78)  75-79 yrs.: 2.75 (2.47 to 3.06)  80-84 yrs.: 4.16 (3.64 to 4.75)  85-89 yrs.: 5.93 (5.23 to 6.73)  90-94 yrs.: 8.28 (7.34 to 9.34)  95+ yrs.: 11.53 (10.10 to 13.17) | In-hospital mortality HR (95% CIs)  Age and comorbidity model:  20-24 yrs.: 0.072 (0.028 to 0.19)  25-29 yrs.: 0.088 (0.041 to 0.19)  30-34 yrs.: 0.11 (0.059 to 0.20)  35-39 yrs.: 0.13 (0.086 to 0.21)  40-44 yrs.: 0.17 (0.12 to 0.23)  45-49 yrs.: 0.21 (0.16 to 0.27)  50-54 yrs.: 0.28 (0.22 to 0.35)  55-59 yrs.: 0.40 (0.34 to 0.49)  60-64 yrs.: 0.63 (0.58 to 0.69)  65-69 yrs.: Reference  70-74 yrs.: 1.54 (1.46 to 1.62)  75-79 yrs.: 2.25 (2.01 to 2.52)  80-84 yrs.: 3.15 (2.74 to 3.62)  85-89 yrs.: 4.25 (3.71 to 4.88)  90-94 yrs.: 5.68 (4.97 to 6.50)  95+ yrs.: 7.58 (6.53 to 8.81) |
| Carter and Collins*  2020  (181) | Location infection acquired, sex, smoking status, C-reactive protein, diabetes, coronary artery diseases, hypertension, reduced renal function, clinical frailty scale | Mortality:  mixed-effects multivariable Cox proportion baseline hazards regression | N= 1564  Deaths: 425 (27.2%)  <65 yrs.: 55/488  65-79 yrs.: 168/535  >80 yrs.: 202/541 | In-hospital mortality HR (95% CI)  <65 yrs.: Reference  65-79 yrs.: 3.30 (2.40-4.55)  >80 yrs.: 4.05 (2.95-5.57) | In-hospital mortality HR (95% CI)  <65 yrs.: Reference  65-79 yrs.: 2.70 (1.91-3.81)  >80 yrs.: 3.30 (2.28-4.78) |
| Chen and Bai  2020  (184) | Sex, hypertension (for overall analysis and for male only stratified analysis), cardiovascular disease, cerebrovascular disease, malignancy, chronic kidney disease, COPD, days from onset to admission | Logistic regression | N= 3309  Deaths: 307 (9.3%)  ≤45 yrs.: NA  >45 yrs.: NA | In-hospital mortality OR (95% CI)  Overall  ≤ 45 yrs.: Reference  > 45 yrs.: 8.37 (4.43–15.81)  Women  ≤ 45 yrs.: 0.32 (0.15-0.70)  > 45 yrs.: Reference  Men  ≤ 45 yrs.: 0.04 (0.01-0.38)  > 45 yrs.: Reference | In-hospital mortality OR (95% CI)  Overall  ≤ 45 yrs.: Reference  > 45 yrs.: 9.08 (4.44-18.59)  Women  ≤ 45 yrs.: 0.29 (0.12-0.67)  > 45 yrs.: Reference  Men  ≤ 45 yrs.: 0.04 (0.01-0.16)  > 45 yrs.: Reference |
| Chilimuri  2020  (187) | Sex, hypertension, diabetes, cardiovascular disease, chronic kidney disease, absolute neutrophil count, absolute lymphocyte count, lactate dehydrogenase, C-reactive protein, D-dimer, ferritin | Multiple regression analysis (not specified) | N= 375  Deaths: 160 (43%) | In-hospital mortality  Per year: OR= 1.05 (95% CI: 1.03-1.73) | In-hospital mortality OR (95% CI)  Per year: 1.04 (1.01-1.06) |
| Ciardullo  2020  (188) | Sex, diabetes, hypertension, chronic kidney disease, cardiovascular disease, COPD | Binomial logistic regression | N=371 (339 for analysis due to missing data)  Deaths: 142 (38.1%) | In-hospital mortality | In-hospital mortality OR (95% CI)  Per year: 1.065 (1.029-1.086) |
| Conversano and Melillo  2020  (190) | COPD, cancer, chronic kidney disease, beta blocker | Cox regression analysis | N=191  Deaths: 42 (22%) | In-hospital mortality  Per year: HR = 1.1 (1.0–1.2) | In-hospital mortality HR (95% CI)  Per year: 1.1 (1.0-1.1) |
| Cummings  2020  (193) | Sex, symptom duration before hospital presentation, hypertension, chronic cardiac disease, COPD or interstitial lung disease, diabetes, interleukin-6, D-dimer | Cox proportional hazards model | N=257  Deaths: 101 (39%) | In-hospital mortality among critically ill patients HR (95% CI)  Per 10-year increase: 1.49 (1.29–1.73)  Per yr. increase (own calculations):  1.04 (1.03-1.06) | In-hospital mortality among critically ill patients HR (95% CI)  Per 10-year increase: 1.31 (1.09-1.57)  Per yr. increase (own calculations):  1.03 (1.01-1.05) |
| Di Castelnuovo  2020  (197) | Gender, hypertension, diabetes, myocardial infarction, heart failure, cancer, lung disease, obesity, smoking, CRP, eGRF/chronic kidney disease stage | Cox proportional hazard regression models with multiple imputation.  Several sensitivity analyses on multiple imputations and accounting for clustering | N=3894  Deaths: 712 (18.3%)  18-44 yrs.: 6/348  45-64 yrs.: 75/1413  65-74 yrs.: 145/808  75-84 yrs.: 266/849  85+ yrs.: 220/476 | In-hospital mortality (main analysis)  HR (95% CI)  18-44 yrs.: Reference  45-64 yrs.: 2.40 (1.04-5.51)  65-74 yrs.: 7.13 (3.14-16.14)  75-84 yrs.: 13.56 (6.03-30.52)  85+ yrs.: 21.65 (9.60-48.82) | In-hospital mortality (main analysis)  HR (95% CI)  18-44 yrs.: Reference  45-64 yrs.: 1.77 (0.84-3.71)  65-74 yrs.: 3.87 (2.03-7.37)  75-84 yrs.: 6.05 (3.17-11.55)  85+ yrs.: 8.24 (4.61-14.74) |
| Docherty  2020  (198) | Sex at birth, chronic cardiac disease, chronic pulmonary disease, chronic kidney disease, diabetes, obesity, chronic neurological disorder, dementia, malignancy, moderate/severe liver disease | Cox proportional hazards models including geographical region as random intercept | N= 20,133  Deaths: 5165 (25.7%)  <50 yrs.: 121/2795  50-59 yrs.: 274/2708  60-69 yrs.: 662/3296  70-79 yrs.: 1506/4692  ≥80 yrs.: 2602/6642 | In-hospital mortality HR (95% CI)  <50 yrs.: Reference  50-59 yrs.: 2.55 (2.06-3.17)  60-69 yrs.: 5.45 (4.48-6.63)  70-79 yrs.: 9.76 (8.09-11.77)  ≥80 yrs.: 13.47 (11.20-16.20) | In-hospital mortality HR (95% CI)  <50 yrs.: Reference  50-59 yrs.: 2.63 (2.06-3.35)  60-69 yrs.: 4.99 (3.99-6.25)  70-79 yrs.: 8.51 (6.85-10.57)  ≥80 yrs.: 11.09 (8.93-13.77) |
| Giacomelli  2020  (209) | Gender, age unadjusted Charlson comorbidity index, obesity, being treated with at least one anti-hypertensive agent, disease severity, presence of anaemia, lymphocyte count, D-dimer, C-reactive protein, creatinine, creatinine kinase | Cox proportional hazards | N=233  Deaths: 48 (20.6%) | In-hospital mortality HR (95% CI)  Age (per 10 years): 1.81 (1.44-2.28)  Age (per year, own calculations): 1.06 (1.04-1.09) | In-hospital mortality HR (95% CI)  Age (per 10 years): 2.08 (1.48-2.92)  Age (per year, own calculations): 1.08 (1.04-1.11) |
| Grasselli  2020  (213) | Sex, respiratory support, hypertension, hypercholesterolemia, heart disease, type 2 diabetes, malignancy, COPD, ACE inhibitor therapy, ARB therapy, statin, diuretic, PEEP at admission, FiO2 at admission, PaO2/FiO2 at admission | Cox proportional hazards regression | N = 3988  Deaths: 1926 (48.29%)  <56 yrs.: 245/997  56-63 yrs.: 416/997  64-69 yrs.: 562/997  >69 yrs.: 703/997 | In-hospital mortality HR (95% CI)  <56 yrs.: Reference  56-63 yrs.: 1.91 (1.63-2.24)  64-69 yrs.: 2.98 (2.56-3.46)  >69 yrs.: 4.25 (3.68-4.92) | In-hospital mortality HR (95% CI)  Age (per 10 years): 1.75 (1.60-1.92)  Age (per year, own calculations): 1.06 (1.05-1.07) |
| Gupta  2020  (216) | Sex, race, hypertension, diabetes, BMI, coronary artery disease, congestive heart failure, chronic obstructive pulmonary disease, current smoker, active cancer, ≤3 d from symptom onset to ICU day 1, lymphocyte count <1000/µL on ICU day 1, invasive mechanical ventilation support/O2 level, shock on ICU day 1, coagulation component of SOFA score, liver component of SOFA score, renal component of SOFA score, no. of ICU beds | Multilevel logistic regression modelling to account for hospitals  Cox regression | N= 2215  Deaths: 784 (35.4%)  Mechanical ventilation: 1494 (67.44%)  28-day in-hospital mortality  18-39 yrs.: 31/209  40-49 yrs.: 64/282  50-59 yrs.: 124/487  60-69 yrs.: 239/614  70-79 yrs.: 201/424  ≥80 yrs.: 125/199 | NA | 28-day in-hospital mortality for ICU patients  OR (95% CI)  Multilevel logistic regression  18-39 yrs.: Reference  40-49 yrs.: 1.65 (0.97-2.80)  50-59 yrs.: 1.71 (1.05-2.80)  60-69 yrs.: 3.18 (1.95-5.18)  70-79 yrs.: 5.36 (3.20-9.00)  ≥80 yrs.: 11.15 (6.19-20.06)  Cox regression model (time-to-event analyses)  HR (95% CI)  18-39 yrs.: Reference  40-49 yrs.: 1.29 (0.85-1.96)  50-59 yrs.: 1.39 (0.94-2.04)  60-69 yrs.: 2.09 (1.43-3.04)  70-79 yrs.: 2.70 (1.83-3.98)  ≥80 yrs.: 4.55 (3.00-6.92)  28-day mortality in mechanically ventilated patients OR (95% CI)  Multilevel logistic regression  18-39 yrs.: Reference  40-49 yrs.: 1.28 (0.70-2.33)  50-59 yrs.: 1.47 (0.84-2.58)  60-69 yrs.: 2.39 (1.37-4.18)  70-79 yrs.: 3.39 (1.86-6.20)  ≥80 yrs.: 9.20 (4.55-18.60) |
| Hashemi  2020  (220) | Chronic liver disease, obesity, sex, cardiac diseases, hypertension, diabetes, hyperlipidaemia, pulmonary disorders | Logistic regression | N=363  Deaths: NA | NA | In-hospital mortality OR (95% CI)  Age (per year): 1.08 (1.05-1.12) |
| Hewitt*  2020  (221) | Sex, smoking status, increased C-reactive  protein, diabetes, coronary artery disease, hypertension, impaired renal function, clinical frailty scale | Cox proportional baseline hazards model (Mortality)  Logistic regression (Day-7 Mortality) | N=1564  Deaths: 425 (27.2%)  <65 yrs.: 55/488  65-79 yrs.: 168/535  ≥80 yrs.: 202/541 | In-hospital mortality HR (95% CI)  <65 yrs.: Reference  65-79 yrs.: 3.30 (2.40-4.55)  ≥80 yrs.: 4.05 (2.95-5.57)  Day-7 mortality OR (95% CI)  <65 yrs.: Reference  65-79 yrs.: 4.62 (2.89-7.39)  ≥80 yrs.: 6.68 (4.18-10.67) | In-hospital mortality HR (95% CI)  <65 yrs.: Reference  65-79 yrs.: 2.58 (1.82-3.64)  ≥80 yrs.: 2.92 (2.02-4.22)  Day-7 mortality OR (95% CI)  <65 yrs.: Reference  65-79 yrs.: 2.79 (1.66-4.72)  ≥80 yrs.: 3.51 (2.01-6.15) |
| Hwang  2020  (224) | Diabetes mellitus, chronic lung disease, cardiovascular disease, Alzheimer’s dementia, stroke | Cox proportional hazards regression | N=103  Deaths: 26 (25%) | NA | In-hospital mortality HR (95% CI)  Age (per year): 1.055 (1.003-1.109) |
| Imam  2020  (225) | Charlson Comorbidity Index, non-steroidal anti-inflammatory medication, angiotensin converting enzyme-inhibitor/angiotensin receptor blocker | Logistic regression | N=1305  Deaths: 200 (15.3%)  Age<60: NA  Age>60: NA | In-hospital mortality OR (95%CI)  Age<60: Reference  Age>60: 3.66 (2.57-5.20) | In-hospital mortality OR (95%CI)  Age<60: Reference  Age>60: 1.93 (1.26-2.94) |
| Jun  2020  (226) | Sex, race, Manhattan facility, hypertension, diabetes, coronary artery disease, heart failure, atrial fibrillation, chronic kidney disease, COPD/asthma, obesity, cancer, oxygen saturation | Logistic regression | N=3086  Deaths: 885 (28.7%) | In-hospital mortality OR (95% CI)  Age (per year): 1.06 (1.05-1.06) | In-hospital mortality OR (95% CI)  Age (per year): 1.06 (1.05-1.07) |
| Khalil  2020  (229) | 11 in total, not all specified  Sex, comorbidities (2-3/4+, including asthma, COPD, cardiovascular disease, hypertension, hyperlipidemia, diabetes, kidney disease, cerebrovascular accident, dementia, malignancy, liver disease, other comorbidities), respiratory rate, systolic blood pressure, lymphocyte count, platelet count, CRP, urea, creatinine, albumin | Cox proportional hazards regression | N= 220  Deaths: 58 (26,4%) | NA | 28-days in-hospital mortality HR (95% CI)  Age (per year): 1.04 (1.01-1.07) |
| Kim  2020  (231) | Sex, race, ethnicity, smoker, hypertension, obesity, diabetes, chronic lung disease, cardiovascular disease, neurologic, renal, immunosuppression, gastrointestinal or liver, hematologic, rheumatologic or autoimmune, outpatient ACE-inhibitor use, angiotensin receptor blocker use prior to hospitalization | Log-linked Poisson generalized estimating equation regressions with an exchangeable correlation matrix | N= 2490  Deaths: 420 (16.9%)  18-39 yrs.: 6/302  40-49 yrs.: 10/318  50-64 yrs.: 74/744  65-74 yrs.: 103/478  75-84 yrs.: 120/397  ≥85 yrs.: 107/251 | In-hospital mortality RR (95% CI)  18-39 yrs.: Reference  40-49 yrs.: 1.51 (0.59-3.85)  50-64 yrs.: 4.62 (2.1-10.18)  65-74 yrs. : 9.88 (4.28-22.85)  75-84 yrs.: 13.89 (6.12-31.52)  ≥85 yrs.: 19.46 (9.39-40.35) | In-hospital-mortality RR (95% CI)  18-39 yrs.: Reference  40-49 yrs.: 1.23 (0.51-2.99)  50-64 yrs.: 3.11 (1.50-6.46)  65-74 yrs.: 5.77 (2.64-12.64)  75-84 yrs.: 7.67 (3.35-17.59)  ≥85 yrs.: 10.98 (5.09 -23.69) |
| Klang  2020  (233) | Sex, cancer, coronary artery disease, congestive heart failure, hypertension, diabetes, hyperlipidemia, chronic kidney disease, smoking, BMI, race | Multivariable logistic regression | N= 3406  Deaths: 1136 (33.3%)  Age ≤50 yrs.: 60/572  Age >50 yrs.: 1076/2834 | NA | In-hospital mortality OR (95% CIs)  Age ≤50yrs.  Per decile: 3.0 (1.9-4.8)  Age >50 yrs.  Per decile: 1.7 (1.6-1.8) |
| Li  2020  (238) | Sex, diabetes, malignancy, hypertension, coronary heart disease, arrhythmia, cerebrovascular disease | Cox regression analysis | N= 596  Deaths: 54 (9.1%)  <65 yrs.: NA  ≥65 yrs.: NA | In-hospital mortality HR (95% CI)  Age <65 years: Reference  Age ≥65 years: 4.284 (2.385–7.694) | In-hospital mortality HR (95% CI)  Age <65 years: Reference  Age ≥65 years: 3.007 (1.634–5.533) |
| Magleby  2020  (241) | Sex, race, coronary artery disease, congestive heart failure, cerebrovascular disease, hypertension, COPD, days of symptoms prior to admission, fever, cough, headache, myalgias, nausea/vomiting, altered mental status, ageusia, highest level of supplemental oxygen, X-ray findings, viral load | Logistic regression | N= 678  Deaths: 127 (18.7%) | NA | In-hospital mortality OR (95% CI)  Per year: 1.10 (1.07-1.13) |
| Meng  2020  (248) | Sex, hypertension, coronary heart disease, diabetes, COPD, chronic kidney disease, cerebrovascular disease, hepatitis, tuberculosis, tumor | Logistic regression | N= 2556 (non-cancer patients)  N= 2665 (cancer and non-cancer patients)  Deaths: 293 (11.0%)  ≤49 yrs.: 14/707  50-64 yrs.: 80/907  65-79 yrs.: 142/866  ≥80 yrs.: 57/185 | In-hospital mortality OR (95% CI)  ≤49 years : Reference  50-64 years: 4.79 (2.69-8.52)  65-79 years: 9.70 (5.55-16.96)  ≥80 years : 22.03 (11.92-40.71) | In-hospital mortality OR (95% CI)  ≤49 years : Reference  50-64 years: 5.06 (2.82-9.06)  65-79 years: 9.97 (5.64-17.63)  ≥80 years : 20.33 (10.76-38.44) |
| Murillo-Zamora and Hernandez-Suarez  2020  (255) | Sex, tobacco use, obesity, asthma, COPD, diabetes, hypertension, immunosuppression, chronic kidney disease, clinically diagnosed pneumonia at hospital | Cox proportional hazards model | N= 66,123  Deaths: 32,039 (48,5%)  20-29 yrs.: 367/2,264  30-44 yrs.: 2,788/10,616  45-59 yrs.: 9,367/22,415  60+ yrs.: 19,517/30,828 | NA (Bivariate analyses) | In-hospital mortality HR (95% CI)  20-29 yrs.: Reference  30-44 yrs.: 1.60 (1.44-1.79)  45-59 yrs.: 2.37 (2.13-2.63)  60+ yrs.: 3.55 (3.20-3.94) |
| Narain  2020  (257) | CCS treatment group, sex, race, ethnicity, insurance, smoking status, mechanical ventilation, vasopressors, eosinophils, platelets, hemoglobin, eGFR, AST, ALT, sodium, ferritin, CRP, D-dimer, LDH, NLR, hospital, Charlson index, asthma, COPD, chronic liver disorder, diabetes, autoimmune disorder, cardiovascular disease, hypertension, interstitial lung disease, kidney disease, hemodialysis, BMI | Cox regression model | N= 5,776  Deaths: NA | NA | In-hospital mortality among CCS patients  Per year increase: HR= 1.03 (95% CI: 1.02-1.04) |
| Palaiodimos  2020  (259) | Sex, BMI, heart failure, coronary artery disease, diabetes, chronic kidney disease or end stage renal disease, COPD, smoking status | Logistic regression, forward stepwise approach | N= 200  Deaths: 48 (24.0%) | In-hospital mortality OR (95%CI)  Per year increase: 1.03 (1.01–1.06) | In-hospital mortality OR (95%CI)  Per year increase: 1.03 (1.00-1.07) |
| Patel  2020  (261) | Therapeutic anticoagulation, home antiplatelet, race, BMI, Charlson score, glucose level | Poisson regression model and logistic regression | N=1716  Deaths= 125 (7.3%)  Deaths in critical: 125 (7.3%)  18-44 yrs.: NA  45- 59 yrs.: NA  60-69 yrs.: NA  70-79 yrs.: NA  >80 yrs.: NA | NA | In-hospital mortality  18-44 yrs.: IRR= 1.36 (95% CI: 0.57-3.28)  45-59 yrs.: Reference  60-69 yrs.: IRR= 1.30 (95% CI: 0.69-2.46)  70-79 yrs.: IRR= 1.31 (95% CI: 0.65-2.65)  >80 yrs.: IRR= 2.36 (95% CI: 1.12-2.47)  Death in critical  18-44 yrs.: OR= 1.43 (95% CI: 0.49-4.17)  45- 59 yrs.: Reference  60-69 yrs.: OR= 1.82 (95% CI: 0.83-3.97)  70-79 yrs.: OR= 1.36 (95% CI: 0.55-3.37)  >80 yrs.: OR= 2.74 (95% CI: 1.03-7.32) |
| Perez-Guzman  2020  (262) | Race, sex, Eixhauser index | Logistic regression | N= 559  Deaths: 178 (32%) | NA | In-hospital mortality:  Per year: OR= 1.05 (1.03-1.06) |
| Petrilli  2020  (263) | Week, race/ethnicity, smoking status, BMI, coronary artery disease, heart failure, hyperlipidemia, hypertension, diabetes, asthma or COPD, chronic kidney disease, cancer, sex | Competing risk model | N= 5279 (positive test)  N= 2741 (hospitalized)  Deaths:  665 (24.3%)  19-44 yrs.: NA  45-54 yrs.: NA  55-64 yrs.: NA  65-74 yrs.: NA  ≥75 yrs.: NA | NA | In-hospital death or discharge to hospice: HR (95% CIs)  Adjusted  19-44 yrs.: Reference  45-54 yrs.: 2.59 (1.66-4.32)  55-64 yrs.: 4.40 (2.73-7.11)  65-74 yrs.: 6.99 (4.34-11.27)  ≥75 yrs.: 10.34 (6.37-16.79) |
| Price-Haywood  2020  (266) | Race, sex, Charlson Comorbidity Index score, low-income residency, obesity | Cox proportional-hazards models  For variables for which less than 25% of data missing, values were imputed | N = 1382  Deaths: 326 (23.6%) | In-hospital mortality: HR (95% CI)  Adjusted for race and sex:  Per 5-yr. unit increase:  1.18 (1.13-1.23)  Per 1-yr unit increase (own calculations): 1.03 (1.02-1.04) | In-hospital mortality: HR (95% CI)  Fully adjusted:  Per 5-yr. unit increase: 1.19 (1.13-1.24)  Per 1-yr unit increase (own calculations): 1.04 (1.02-1.04) |
| Reilev  2020  (269) | Number of comorbidities (0,1,2,3,4+ of chronic lung disease, hypertension, ischaemic heart disease, heart failure, atrial fibrillation, stroke, diabetes, dementia, cancer, chronic liver disease, hospital-diagnosed kidney disease, alcohol abuse, substance abuse, major psychiatric disorders, organ transplantation, medical overweight and obesity, rheumatoid arthritis/connective-tissue disease), sex | Logistic regression | N= 2254 hospitalized (for ICU, in-hospital mortality)  Deaths: NA  0-9 yrs.: NA/11  10-19 yrs.: NA/14  20-29 yrs.: NA/55  30-39 yrs.: NA/90  40-49 yrs.: NA/192  50-59 yrs.: NA/337  60-69 yrs.: NA/374  70-79 yrs.: NA/578  80-89 yrs.: NA/476  90+ yrs.: NA/127 | In-hospital mortality OR (95% CI)  Unadjusted  0-9 yrs.: NA  10-19 yrs.: NA  20-29 yrs.: NA  30-39 yrs.: NA  40-49 yrs.: NA  50-59 yrs.: Reference  60-69 yrs.: 3.5 (1.9-6.4)  70-79 yrs.: 7.2 (4.1-12.8)  80-89 yrs.: 13.3 (7.5-23.4)  90+ yrs.: 32.2 (17.0-61.1) | In-hospital mortality OR (95% CI)  Adjusted  0-9 yrs.: NA  10-19 yrs.: NA  20-29 yrs.: NA  30-39 yrs.: NA  40-49 yrs.: NA  50-59 yrs.: Reference  60-69 yrs.: 2.9 (1.6-5.4)  70-79 yrs.: 5.2 (2.9-9.2)  80-89 yrs.: 10.2 (5.7-18.2)  90+ yrs.: 29.1 (15.0-56.5) |
| Salacup  2020  (277) | Sex, BMI, ethnicity, COPD and asthma, diabetes, hypertension, cirrhosis, heart failure, chronic kidney disease | Logistic regression | N= 242  Deaths: 52 (21,5%) | NA | In hospital mortality OR (95% CI)  Per year: 1.056 (1.023-1.090) |
| Sapey  2020  (279) | Ethnicity, sex, deprivation, comorbidity number group 0, 1-2, 3+ (hypertension, cerebrovascular disease, atrial fibrillation, ischaemic heart disease/angina/myocardial infarct, diabetes (type 1 and 2), asthma, COPD, interstitial lung disease, chronic kidney disease, any active malignancy, dementia, obesity) | Cox proportional hazard regression | N=2169  Deaths: 769 (34.6%) | NA | In-hospital mortality HR (95% CI)  Age (z-score): 2.4 (1.8-3.2)  Age (z-score) x sex(male): 1.2 (1.0-1.5)  Age (z-score) x comorbidity group 1-2: 0.9 (0.7-1.2)  Age (z-score) x comorbidity group 3+: 0.7 (0.5-0.9) |
| Seiglie  2020  (280) | BMI, sex, race/ethnicity, diabetes, coronary artery disease or myocardial infarction, chronic heart failure, hypertension, COPD/asthma, active cancer, liver disease, renal disease | Logistic regression | N=450  Deaths: 49 (10.9%)  <50 yrs.: NA  50-59 yrs.: NA  60-69 yrs.: NA  ≥70 yrs.: NA | 14-day in-hospital death OR (95% CIs)  <50 yrs.: Reference  50-59 yrs.: 7.05 (0.81-61.7)  60-69 yrs.: 8.35 (0.98-70.8)  ≥70 yrs.: 27.03 (3.65-200.2) | 14-day in-hospital death OR (95% CI)  <50 yrs.: Reference  50-59 yrs.: 3.74 (0.40-34.54)  60-69 yrs.: 4.57 (0.50-41.9)  ≥70 yrs.: 12.66 (1.50-106.56) |
| Shah  2020  (281) | BMI, sex, ethnicity, hypertension, coronary artery disease, congestive heart failure, COPD, asthma, chronic kidney disease, diabetes, immunosuppression, chronic liver disease, cancer, tobacco smoking | Logistic regression | N= 522  Deaths: 92 (17.6%)  <65 yrs: NA  ≥65 yrs.: NA  ------  <20 yrs.: 0  20-29 yrs.: 1/19  30-39 yrs.: 1/34  40-49 yrs.: 8/58  50-59 yrs.: 7/111  60-69 yrs.: 29/137  70-79 yrs.: 27/104  80-89 yrs.: 13/42  90-99 yrs.: 6/17 | NA | In-hospital mortality OR (95% CI)  <65 yrs: Reference  ≥ 65 yrs.: 3.1 (1.7-5.6) |
| Shi  2020  (285) | Cardiovascular disease, cerebrovascular disease, diabetes, COPD, renal failure, cancer, acute respiratory distress syndrome, cardiac injury, creatinine, N-terminal pro-B-type natriuretic peptide | Cox proportional hazard regression model | N= 416  Deaths: 57 (13.7%) | NA | In-hospital mortality  From symptom onset HR (95% CI)  Age (per year): 1.02 (0.99-1.05)  From admission HR (95% CI)  Age (per year): 1.02 (0.99-1.04) |
| van Gerwen  2020  (301) | Sex, race, smoking, BMI, hypertension, coronary artery disease, atrial fibrillation, congestive heart failure, peripheral vascular disease, cerebrovascular accident/transient ischemic attack, dementia, diabetes, hypothyroidism, chronic kidney disease, malignancy, asthma, COPD, prior venous thromboembolism | Logistic regression | N=3703 (all patients)  N=2015 (hospitalized)  Deaths: 616 (30.6% of hospitalized)  18-40 yrs.: 8/207  40-60 yrs.: 101/539  >60 yrs.: 507/1269 | In-hospital mortality OR (95% CI)  18-40 yrs.: Reference  40-60 yrs.: 5.74 (2.74‐12.01)  >60 yrs.: 16.55 (8.09‐33.85) | In-hospital mortality OR (95% CI)  18-40 yrs.: Reference  40-60 yrs.: 5.29 (2.51‐11.15)  >60 yrs.: 13.04 (6.25‐27.24) |
| Wang  2020  (305) | Model 1:  Cardiovascular disease, cerebrovascular disease, COPD | Cox regression | N=7592  Deaths: 65 (19.2%) | In-hospital mortality in patients over 60 years old HR (95% CI)  Age (per year): 1.084 (1.055-1.114) | In-hospital mortality in patients over 60 years old HR (95% CI)  Age (per year): 1.064 (1.034-1.096) |
| Yehia  2020  (312) | Model 2:  Race, sex, insurance, neighborhood deprivation index, Elixhauser Comorbidity Index (ECI score)  Model 3:  Race, sex, insurance, neighborhood deprivation index, ECI score, asthma, cancer, chronic kidney disease, COPD, congestive heart failure, coronary artery disease, diabetes, obesity | Cox proportional hazards regression | N=7139  Deaths: 1446 (20.3%)  18-49 yrs.: NA  50-64 yrs.: NA  65-84 yrs.: NA  ≥85 yrs.: NA | In-hospital mortality HR (95% CI)  Model 1 (unadjusted)  18-49 yrs.: Reference  50-64 yrs.: 1.54 (1.17-2.03)  65-84 yrs.: 3.36 (2.61-4.34)  ≥85 yrs.: 5.65 (4.32-7.38) | In-hospital mortality HR (95% CI)  Model 2 (adjusted)  18-49 yrs.: Reference  50-64 yrs.: 1.40 (1.13-1.73)  65-84 yrs.: 2.44 (1.79-3.33)  ≥85 yrs.: 4.00 (2.95-5.47)  Model 3 (adjusted)  18-49 yrs.: Reference  50-64 yrs.: 1.36 (1.11-1.67)  65-84 yrs.: 2.38 (1.73-3.26)  ≥85 yrs.: 3.96 (2.82-5.55) |
| Zhao  2020  (319) | Sex, comorbidities (hypertension, diabetes, coronary heart disease, COPD, asthma, cerebrovascular disease, chronic renal disease, chronic liver disease, malignancy, autoimmune disease, organ transplantation) | Cox proportional hazard regression | N=1000  Deaths: 119 (11.9%)  <60 yrs.: 24/473  60-75 yrs.: 42/359  ≥75 yrs.: 53/168 | NA | In-hospital mortality HR (95% CI)  Model 1 (cumulative death risk after admission):  <60 yrs.: Reference  60-75 yrs.: 1.944 (1.156-3.271)  ≥75 yrs.: 4.777 (2.850-8.008)  Model 2 (cumulative death risk after disease onset):  <60 yrs.: Reference  60-75 yrs.: 1.849 (1.1-3.108)  ≥75 yrs.: 4.770 (2.841-8.008) |
| *: same study population  ICU: intensive care unit; COPD: chronic obstructive pulmonary disease; BMI: body mass index; yrs.: years; SOFA score: sequential organ failure assessment score; OR: odds ratio; HR: hazards ratio; CI: confidence intervals; NA: not available | | | | | |

**Table S4. Risk of bias summary for studies investigating in-hospital mortality**

| **Study** | **Major domains** | | | | | **Minor domains** | | | **OVERALL** |
| --- | --- | --- | --- | --- | --- | --- | --- | --- | --- |
|  | **Recruitment procedure** | **Exposure assessment** | **Outcome source and validation** | **Confounding** | **Analysis method** | **Chronology** | **Funding** | **Conflict of interest** |  |
| Antwi-Amoabeng et al. 2020  (165) | 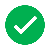 | 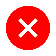 | 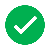 | 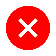 | 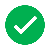 | 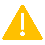 | 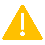 | 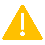 | 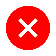 |
| Argenziano et al. 2020  (166) | 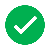 | 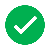 | 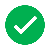 | 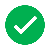 | 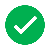 | 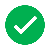 | 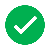 | 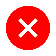 | 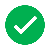 |
| Baqui et al. 2020  (168) | 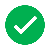 | 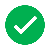 | 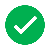 | 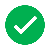 | 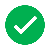 | 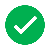 | 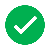 | 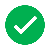 | 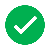 |
| Boulle et al. 2020  (176) | 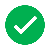 | 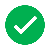 | 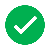 | 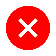 | 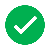 | 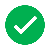 | 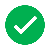 | 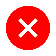 | 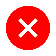 |
| Burn et al. 2020  (177) | 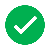 | 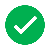 | 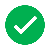 | 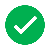 | 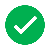 | 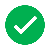 | 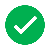 | 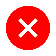 | 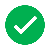 |
| Carter et al. 2020  (181) | 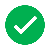 | 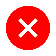 | 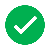 | 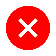 | 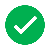 | 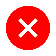 | 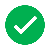 | 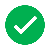 | 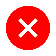 |
| Chen et al. 2020  (184) | 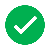 | 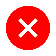 | 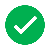 | 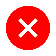 | 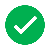 | 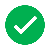 | 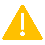 | 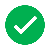 | 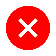 |
| Chilimuri et al. 2020  (187) | 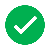 | 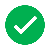 | 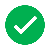 | 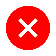 | 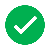 | 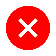 | 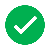 | 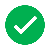 | 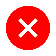 |
| Ciardullo et al. 2020  (188) | 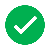 | 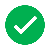 | 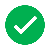 | 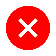 | 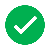 | 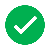 | 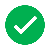 | 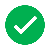 | 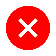 |
| Conversano et al. 2020  (190) | 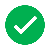 | 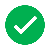 | 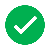 | 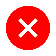 | 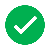 | 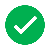 | 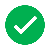 | 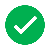 | 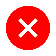 |
| Cummings et al. 2020  (193) | 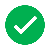 | 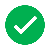 | 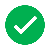 | 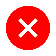 | 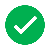 | 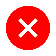 | 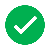 | 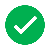 | 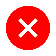 |
| Di Castelnuovo et al. 2020  (197) | 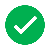 |  |  |  |  |  |  |  |  |
| Docherty et al. 2020  (198) |  |  |  |  |  |  |  |  |  |
| Giacomelli et al. 2020  (209) |  |  |  |  |  |  |  |  |  |
| Grasselli et al. 2020  (213) |  |  |  |  |  |  |  |  |  |
| Gupta et al. 2020  (216) |  |  |  |  |  |  |  |  |  |
| Hashemi et al. 2020  (220) |  |  |  |  |  |  |  |  |  |
| Hewitt et al. 2020  (221) |  |  |  |  |  |  |  |  |  |
| Hwang et al. 2020  (224) |  |  |  |  |  |  |  |  |  |
| Imam et al. 2020  (225) |  |  |  |  |  |  |  |  |  |
| Jun et al. 2020  (226) |  |  |  |  |  |  |  |  |  |
| Khalil et al. 2020  (229) |  |  |  |  |  |  |  |  |  |
| Kim et al. 2020  (231) |  |  |  |  |  |  |  |  |  |
| Klang et al. 2020  (233) |  |  |  |  |  |  |  |  |  |
| Li et al. 2020  (238) |  |  |  |  |  |  |  |  |  |
| Magleby et al. 2020  (241) |  |  |  |  |  |  |  |  |  |
| Meng et al. 2020  (248) |  |  |  |  |  |  |  |  |  |
| Murillo-Zamora et al. 2020  (255) |  |  |  |  |  |  |  |  |  |
| Narain et al. 2020  (257) |  |  |  |  |  |  |  |  |  |
| Palaiodimos et al. 2020  (259) |  |  |  |  |  |  |  |  |  |
| Patel et al. 2020  (261) |  |  |  |  |  |  |  |  |  |
| Perez-Guzman et al. 2020  (262) |  |  |  |  |  |  |  |  |  |
| Petrilli et al. 2020  (263) |  |  |  |  |  |  |  |  |  |
| Price-Haywood et al. 2020  (266) |  |  |  |  |  |  |  |  |  |
| Reilev et al. 2020  (269) |  |  |  |  |  |  |  |  |  |
| Salacup et al. 2020  (277) |  |  |  |  |  |  |  |  |  |
| Sapey et al. 2020  (279) |  |  |  |  |  |  |  |  |  |
| Seiglie et al. 2020  (280) |  |  |  |  |  |  |  |  |  |
| Shah et al. 2020  (281) |  |  |  |  |  |  |  |  |  |
| Shi et al. 2020  (285) |  |  |  |  |  |  |  |  |  |
| van Gerwen et al. 2020  (301) |  |  |  |  |  |  |  |  |  |
| Wang et al. 2020  (305) |  |  |  |  |  |  |  |  |  |
| Yehia et al. 2020  (312) |  |  |  |  |  |  |  |  |  |
| Zhao et al. 2020  (319) |  |  |  |  |  |  |  |  |  |
| = low risk of bias = unclear risk of bias = high risk of bias | | | | | | | | | |

**Figure S3. Relationship between median age and log of relative risk among studies using age as a categorical value (linear and cubic models, variance weighted): in-hospital mortality**

**Figure S4. Funnel plot of studies investigating in-hospital mortality**

**Figures S5a-b. Funnel plot of studies investigating in-hospital mortality with age as continuous variable (a) and as categorical value (b)**

**(a)	(b)**

**Table S5. Results of studies investigating case mortality**

| **Author,**  **Year**  **(Ref)** | **Confounders and**  **age-dependent risk factors used in the model** | **Type of analysis**  **# cases / # non-cases** | **N in analysis**  **Total deaths**  **In age group: # cases/total** | **Results**  **Unadjusted effect estimates (if available)** | **Results**  **Adjusted effect estimates** |
| --- | --- | --- | --- | --- | --- |
| Bello-Chavolla†  2020  (170) | Diabetes, diabetes (interaction with age<40 yrs.), smoking, immunosuppression, COPD, asthma, CVD, chronic kidney disease, hypertension, sex, obesity | Cox proportional risk regression | N= 51,633  Deaths: 5332 (10.3%)  <40 yrs.: NA  ≥40yrs. - ≤65 yrs.: NA  >65 yrs.: NA |  | 30-day mortality (population based)  Results available in figures. |
| Boulle  2020  (176) | Sex, diabetes, hypertension, chronic kidney disease, chronic pulmonary disease/asthma, tuberculosis, HIV, antiretroviral therapy, CD4 count during COVID-19 | Cox proportional hazards model | All public-sector SARS-CoV-2 cases diagnosed before June 1, 2020  N= 15,203  Deaths: 510  20-39 yrs.: 35/8688  40-49 yrs.: 51/3042  50-59 yrs.: 137/2018  60-69 yrs.: 150/889  ≥70 yrs.: 137/566 | NA | Mortality HR (95% CI)  All public-sector SARS-CoV-2 cases diagnosed before June 1, 2020  20-39 yrs.: Reference  40-49 yrs.: 3.19 (2.06-4.93)  50-59 yrs.: 10.84 (7.34-16.01)  60-69 yrs.: 24.87 (16.67-37.11)  ≥70 yrs.: 38.32 (25.47-57.64) |
| Burn  2020  (177) | Autoimmune condition, chronic kidney disease, COPD, dementia, heart disease, hyperlipidemia, hypertension, malignant neoplasm, obesity, Type 2 diabetes, Charlson Index | Cox proportional hazards model using a non-linear relationship with age | From diagnosed with COVID-19 to death  N= 102,002  Deaths: 2581  >18 yrs.: <5/4536  18–39 yrs.: 5/30,530  40-59 yrs.: 63/44,493  60-69 yrs.: 131/10,305  70-79 yrs.: 406/6120  ≥80 yrs.: 1976/6018 | Population based mortality HR (95% CIs)  From diagnosed with COVID-19 to death (Age only model):  20-24 yrs.: 0.0074 (0.0023 to 0.024)  25-29 yrs.: 0.0089 (0.0038 to 0.021)  30-34 yrs.: 0.01 (0.0044 to 0.022)  35-39 yrs.: 0.0097 (0.0044 to 0.021)  40-44 yrs.: 0.0081 (0.0046 to 0.014)  45-49 yrs.: 0.009 (0.006 to 0.014)  50-54 yrs.: 0.023 (0.016 to 0.033)  55-59 yrs.: 0.095 (0.077 to 0.12)  60-64 yrs.: 0.35 (0.33 to 0.38)  65-69 yrs.: Reference  70-74 yrs.: 2.30 (2.21 to 2.39)  75-79 yrs.: 4.56 (4.30 to 4.82)  80-84 yrs.: 8.29 (7.76 to 8.85)  85-89 yrs.: 14.69 (13.58 to 15.89)  90-94 yrs.: 26.00 (23.62 to 28.62)  95+ yrs.: 46.03 (40.97 to 51.71) | Population based mortality HR (95% CIs)  From diagnosed with COVID-19 to death (Age and comorbidity model):  20-24 yrs.: 0.0083 (0.0024 to 0.03)  25-29 yrs.: 0.0099 (0.004 to 0.025)  30-34 yrs.: 0.011 (0.0047 to 0.027)  35-39 yrs.: 0.012 (0.005 to 0.027)  40-44 yrs.: 0.011 (0.006 to 0.02)  45-49 yrs.: 0.013 (0.0088 to 0.02)  50-54 yrs.: 0.033 (0.023 to 0.047)  55-59 yrs.: 0.12 (0.099 to 0.15)  60-64 yrs.: 0.40 (0.37 to 0.43)  65-69 yrs.: Reference  70-74 yrs.: 2.05 (1.96 to 2.13)  75-79 yrs.: 3.62 (3.41 to 3.85)  80-84 yrs.: 5.91 (5.48 to 6.36)  85-89 yrs.: 9.38 (8.54 to 10.29)  90-94 yrs.: 14.88 (13.25 to 16.70)  95+ yrs.: 23.60 (20.52 to 27.16) |
| Carrillo-Vega†  2020  (180) | Sex, chronic kidney disease, COPD, diabetes & hypertension & obesity, diabetes & hypertension, diabetes & obesity, only hypertension, only obesity, only diabetes, pneumonia, pregnancy, immune-suppressed, hospitalized, intubated, ICU, health services | Logistic regression | N= 9946 (by outcome)  N= 10544 (in total)  Deaths: 963 (9.7%) (by outcome)  Deaths: 968 (9.2%) (in total)  <25 yrs.: 5/598  25-49 yrs.: 255/5640  50-74 yrs.: 573/3823  ≥75 yrs.: 135/483 | NA | Mortality (population based) OR (95% CI)  25-49 yrs.: Reference  50-74 yrs.: 1.96 (1.63-2.34)  ≥75 yrs.: 3.74 (2.80-4.98) |
| Gu  2020  (214) | Model 1  Sex, pandemic period, comorbidity score (considered coronary heart disease, hypertension, cardiac failure, cerebral infarction, chronic bronchitis, COPD, diabetes, renal failure, history of surgery)  Model 2  Sex, period of the pandemic, coronary heart disease, cerebral infarction, COPD, renal failure | Inverse probability-weighted Cox proportional hazard regression | N= 275  Deaths: 94 (21%)  Controls: 181 | Population-based mortality HR (95% CI)  Age (per year): 1.05 (1.0-1.1) | Population-based mortality HR (95% CI)  Model 1  Age (per year): 1.04 (1.02-1.07)  Model 2  Age (per year): 1.04 (1.02-1.06) |
| Gu  2020  (215) | Sex, BMI, smoking status, alcohol consumption, race/ethnicity, neighborhood socioeconomic disadvantage index, population density, comorbidity score, respiratory disease, circulatory disease, any cancer, type 2 diabetes, kidney disease, liver disease, autoimmune disease | Logistic regression analyses, firth bias-corrected estimates | N=1139 (Positive tests)  Deaths: 88 (7.7%)  <18 yrs.: 0/9  18- <35 yrs.: 2/203  35- <50 yrs.: 5/257  50- <65 yrs.: 16/349  65- <80 yrs.: 31/233  ≥80 yrs.: 34/88 | Mortality (population-based)  OR (95 % CI)  Age (per 10 years): 2.21 (1.87-2.62)  Age (per 1-year, own calculation): 1.08 (1.06-1.10)  <18 yrs.: -  18- <35 yrs.: Reference  35- <50 yrs.: 1.76 (0.39-7.95)  50- <65 yrs.: 3.99 (1.04-15.3)  65- <80 yrs.: 12.5 (3.40-46.3)  ≥80 yrs.: 51.0 (13.6-191) | Mortality (population-based)  OR (95 % CI)  Age (per 10 years): 2.31 (1.78-3)  Age (per 1-year, own calculation): 1.09 (1.06-1.12)  <18 yrs.: -  18- <35 yrs.: Reference  35- <50 yrs.: 1.22 (0.166-9.01)  50- <65 yrs.: 1.91 (0.33-11)  65- <80 yrs.: 6.22 (1.14-33.9)  ≥80 yrs.: 37.5 (6.71-209) |
| Harrison  2020  (219) | Sex, ethnicity, comorbidities within the Charlson comorbidity index (myocardial infarction, congestive heart failure, peripheral vascular disease, cerebrovascular disease, dementia, chronic pulmonary disease, rheumatic disease, peptic ulcer disease, mild liver disease, moderate/severe liver disease, diabetes mellitus, hemiplegia or paraplegia, renal disease, any malignancy, metastatic solid tumour, AIDS/HIV | Logistic regression | N=31,461  Deaths: 1,296 (4.1%)  <50 yrs.: 104/15,578  50-69 yrs.: 430/10,698  70-90 yrs.: 762/5,185 | Population-based mortality  OR (95% CI)  Whole population  Age (per year):  1.074 (1.069-1.078)  Adults aged <50 yrs.:  Age (per year): 1.06 (1.05-1.09)  Adults aged 50-69 yrs.:  Age (per year): 1.10 (1.08-1.12)  Adults aged 70-90 yrs.:  Age (per year): 1.03 (1.01-1.04) | Population-based mortality-  OR (95% CI)  Whole population  Age (per year): 1.063 (1.058-1.068)  Adults aged <50 yrs.  Age (per year): 1.03 (1.01-1.06)  Adults aged 50-69 yrs.  Age (per year): 1.07 (1.07-1.10)  Adults aged 70-90 yrs.  Age (per year): 1.02 (1.01-1.04) |
| Kabarriti  2020  (227) | Sex, socioeconomic status, ethnicity/race, BMI, hypertension, cardiovascular disease, diabetes, cancer, liver disease, dementia, chronic pulmonary disease, peptic ulcer, hemiplegia or paraplegia, kidney disease, HIV/AIDS | Cox proportional hazards models | N=5902  Deaths: 918 (15.5%)  ≤ 40 yrs.: 27/1255  41-60 yrs.: 133/2026  61-80 yrs.: 507/2002  >80 yrs.: 251/619 | Mortality (population based) OR (95% CI)  ≤ 40 yrs.: Reference  41-60 yrs.: 2.42 (1.60-3.66)  61-80 yrs.: 7.63 (5.18-11.2)  >80 yrs.: 14.9 (9.98-22.1) | Mortality (population based) OR (95% CI)  Model 1 (Complete case)  ≤ 40 yrs.: Reference  41-60 yrs.: 1.85 (1.17-2.94)  61-80 yrs.: 5.24 (3.37-8.16)  >80 yrs.: 9.48 (5.98-15.0)  Model 2 (Multiple imputation)  ≤ 40 yrs.: Reference  41-60 yrs.: 2.04 (1.34-3.10)  61-80 yrs.: 5.47 (3.66-8.18)  >80 yrs.: 10.3 (6.75-15.6) |
| Lee  2020  (235) | Sex, hypertension, diabetes, coronary artery disease, stroke, COPD, any cancer, chronic kidney disease, medication (angiotensin receptor blocker, angiotensin converting enzyme) | Generalized linear mixed-effect regression models, including squared term for age | N= 8,266  Deaths: 112 (1.4%) | NA | Population-based mortality within 60 days HR (95% CI)  Age per year: 1.372 (1.118-1.683)  Age^2^: 0.999 (0.997-1.000) |
| Reilev  2020  (269) | Number of comorbidities (0,1,2,3,4+ of chronic lung disease, hypertension, ischaemic heart disease, heart failure, atrial fibrillation, stroke, diabetes, dementia, cancer, chronic liver disease, hospital-diagnosed kidney disease, alcohol abuse, substance abuse, major psychiatric disorders, organ transplantation, medical overweight and obesity, rheumatoid arthritis/connective-tissue disease), sex | Logistic regression | N= 11,122 (positive cases, for hospitalization, death within 30 days)  Deaths within 30 days of positive test: 577 (5.2%)  0-9 yrs.: 0/251  10-19 yrs.: 0/495  20-29 yrs.: 0/1523  30-39 yrs.: NA/1588  40-49 yrs.: NA/1970  50-59 yrs.: NA/2035  30-59: 16/5593  60-69 yrs.: 56/1305  70-79 yrs.: 165/950  80-89 yrs.: 220/762  90+ yrs.: 120/243 | Death within 30 days among positive cases OR (95% CI)  Unadjusted  0-9 yrs.: NA  10-19 yrs.: NA  20-29 yrs.: NA  30-39 yrs.: NA  40-49 yrs.: NA  50-59 yrs.: Reference  60-69 yrs.: 6.0 (3.4-10.7)  70-79 yrs.: 28.3 (16.6-48.3)  80-89 yrs.: 54.7 (32.1-93.0)  90+ yrs.: 131.4 (74.5-231.6) | Death within 30 days among positive cases OR (95% CI)  Adjusted  0-9 yrs.: NA  10-19 yrs.: NA  20-29 yrs.: NA  30-39 yrs.: NA  40-49 yrs.: NA  50-59 yrs.: Reference  60-69 yrs.: 4.4 (2.5-7.9)  70-79 yrs.: 15.2 (8.7-26.3)  80-89 yrs.: 29.9 (17.2-51.9)  90+ yrs.: 90.2 (50.2-162.2) |
| Rossi  2020  (274) | Sex, calendar period, time from symptoms to diagnosis, place of birth, Charlson Comorbidity Index | Cox proportional hazard regression models | N= 2653 COVID-19 cases  Deaths (COVID-19 cases): 217 (8.2%)  <51 yrs.: 2/696  51-60 yrs.: 7/528  61-70 yrs.: 23/413  71-80 yrs.: 54/420  ≥81 yrs.: 131/596 | NA | Population-based mortality  HR (95% CI)  <51 yrs.: Reference  51-60 yrs.: 1.5 (0.5-4.2)  61-70 yrs.: 3.8 (1.6-9.4)  71-80 yrs.: 9.1 (4.0-20.6)  ≥81 yrs.: 27.8 (12.5-61.7) |
| Solís  2020  (294) | Sex, medical unit, week, comorbidities (none or one, two, three or more), hypertension, obesity, cardiovascular disease, diabetes, immunosuppression, COPD, asthma, chronic kidney disease, smoking, other, institution | Piecewise exponential hazards regression model | N=7497  Deaths: 650 (8.67%)  0-24 yrs.: NA  25-29 yrs.: NA  30-34 yrs.: NA  35-39 yrs.: NA  40-44 yrs.: NA  45-49 yrs.: NA  50-54 yrs.: NA  55-59 yrs.: NA  60-64 yrs.: NA  65-69 yrs.: NA  70+ yrs.: NA | NA | Population-based mortality at 35 days  HR (95% CI)  0-24 yrs.: 0.038 (0.005-0.270)  25-29 yrs.: 0.202 (0.104-0.392)  30-34 yrs.: 0.218 (0.121-0.394)  35-39 yrs.: 0.357 (0.225-0.566)  40-44 yrs.: 0.756 (0.530-1.078  45-49 yrs.: Reference  50-54 yrs.: 1.171 (0.854-1.605)  55-59 yrs.: 1.411 (1.035-1.923)  60-64 yrs.: 1.304 (0.926-1.836)  65-69 yrs.: 2.016 (1.458-2.787)  70+ yrs.: 2.608 (1.931-3.522) |
| Sousa  2020  (295) | Sex, cardiovascular disease, diabetes, hematologic disease, neurologic disease, obesity, pneumopathies, renal disease | Robust Poisson regression  Cox regression | N=2070  Deaths: 131 (6.33%)  <60 yrs.: 32/1573  ≥60 yrs.: 99/497 | NA | Population-based mortality IRR (95% CI)  <60 yrs.: Reference  ≥60 yrs.: 3.1 (1.9-5.0)  Population-based mortality HR (95% CI)  <60 yrs.: Reference  ≥60 yrs.: 3.6 (2.3-5.8) |
| Tartof  2020  (300) | BMI, sex, race/ethnicity, smoking, comorbidities: metastatic tumor/cancer, hyperlipidemia, myocardial infarction, other immune condition, organ transplant, congestive heart failure, peripheral vascular disease, cerebrovascular disease, chronic pulmonary disease, renal disease, hypertension, asthma, diabetes mellitus, time | Poisson regression with multiple imputation for missing data. | N=6916  Deaths: 206 (3,0%)  0-40 yrs.: 7/2227  41-50 yrs.: 10/1422  51-60 yrs.: 20/1545  61-70 yrs.: 41/1045  71-80 yrs.: 53/427  >80 yrs.: 75/250 | NA | Population-based mortality RR (95% CI)  0-40 yrs.: Reference  41-50 yrs.: 1.88 (0.71–4.95)  51-60 yrs.: 3.14 (1.31–7.57)  61-70 yrs.: 7.18 (3.08–16.78)  71-80 yrs.: 16.08 (6.72–38.52)  >80 yrs.: 43.21 (17.80–104.92) |
| Wang  2020  (302) | Residential area, smoking status, vital signs, administered medication, asthma, chronic kidney disease, number of comorbidities (0, 1-3, 4+)  Most prevalent; hypertension, diabetes, chronic kidney disease, obesity, cancer, asthma, COPD. | Logistic regression | N=7592 (total)  N=6764 (analysis)  Deaths: 828 (10.9%)  <40 yrs.: 10/1590  40-49 yrs.: 33/940  50-59 yrs.: 80/1306  60-69 yrs.: 167/1583  70-79 yrs.: 217/1137  80+ yrs.: 321/1036 | Population-based mortality OR (95% CI)  <40 yrs.: Reference  40-49 yrs.: 5.75 (2.82-11.72)  50-59 yrs.: 10.31 (5.32-19.98)  60-69 yrs.: 18.63 (9.81-35.41)  70-79 yrs.: 37.27 (19.67-70.61)  80+ yrs.: 70.93 (37.57-133.93) | Population-based mortality OR (95% CI)  <40 yrs.: Reference  40-49 yrs.: 3.48 (1.48-8.17)  50-59 yrs.: 5.53 (2.51-12.19)  60-69 yrs.: 8.44 (3.90-18.26)  70-79 yrs.: 18.29 (8.46-39.51)  80+ yrs.: 33.77 (15.66-72.84) |
| †: same study population  ICU: intensive care unit; COPD: chronic obstructive pulmonary disease; BMI: body mass index; yrs.: years; SOFA score: sequential organ failure assessment score; OR: odds ratio; HR: hazards ratio; CI: confidence intervals; NA: not available | | | | | |

**Table S6. Risk of bias summary for studies investigating case mortality**

| **Study** | **Major domains** | | | | | **Minor domains** | | | **OVERALL** |
| --- | --- | --- | --- | --- | --- | --- | --- | --- | --- |
|  | **Recruitment procedure** | **Exposure assessment** | **Outcome source and validation** | **Confounding** | **Analysis method** | **Chronology** | **Funding** | **Conflict of interest** |  |
| Bello-Chavolla et al. 2020  (170) |  |  |  |  |  |  |  |  |  |
| Boulle et al. 2020  (176) |  |  |  |  |  |  |  |  |  |
| Burn et al. 2020  (177) |  |  |  |  |  |  |  |  |  |
| Carrillo-Vega et al. 2020  (180) |  |  |  |  |  |  |  |  |  |
| Gu et al. 2020  (214) |  |  |  |  |  |  |  |  |  |
| Gu et al. 2020  (215) |  |  |  |  |  |  |  |  |  |
| Harrison et al. 2020  (219) |  |  |  |  |  |  |  |  |  |
| Kabarriti et al. 2020  (227) |  |  |  |  |  |  |  |  |  |
| Lee et al. 2020  (235) |  |  |  |  |  |  |  |  |  |
| Rossi et al. 2020  (274) |  |  |  |  |  |  |  |  |  |
| Solís et al. 2020  (294) |  |  |  |  |  |  |  |  |  |
| Sousa et al. 2020  (295) |  |  |  |  |  |  |  |  |  |
| Tartof et al. 2020  (300) |  |  |  |  |  |  |  |  |  |
| Wang et al. 2020  (302) |  |  |  |  |  |  |  |  |  |
| = low risk of bias = unclear risk of bias = high risk of bias | | | | | | | | | |

**Figure S6. Relationship between median age and log relative risk among studies using age as a categorical value (linear and cubic models, variance weighted): case mortality**

**Figure S7. Funnel plot of studies investigating case mortality**

**Table S7. Results of studies investigating risk of hospitalization**

| **Author,**  **Year**  **(Ref)** | **Confounders and**  **age-dependent risk factors used in the model** | **Type of analysis** | **N in analysis**  **Total hospitalized**  **In age group: # cases/total** | **Results**  **Unadjusted effect estimates (if available)** | **Results**  **Adjusted effect estimates** |
| --- | --- | --- | --- | --- | --- |
| Azar  2020  (167) | Adjusted Model 2:  Race/ethnicity, sex, homeless status, smoking status, type 2 diabetes, hypertension, depression, congestive heart failure, cardiovascular disease, cancer, chronic obstructive pulmonary disease, asthma  Adjusted Model 3:  Confounders in adjusted model 2 plus insurance status and income | Logistic regression Models built stepwise that incrementally included more covariates. | N= 1052  Admitted to hospital: 256 (24.3%)  18-29 yrs. 2/115  30-39 yrs. 19/170  18-39 yrs. 21/285  40-49 yrs. 25/174  50-59 yrs. 41/201  60-69 yrs. 62/184  70-79 yrs. 47/114  80+ yrs. 60/94 | Hospital admission  Unadjusted Model  18-39 yrs. Reference  40-49 yrs.: OR= 2.11 (1.14, 3.90)  50-59 yrs.: OR= 3.22 (1.84, 5.65)  60-69 yrs.: OR= 6.39 (3.72, 10.96)  70-79 yrs.: OR= 8.82 (4.94, 15.75)  80+ yrs.: OR= 22.18 (12.03, 40.91) | Hospital admission  Model 2  18-39 yrs. Reference  40-49 yrs.: OR= 1.91 (95% CI: 1.00-3.65)  50-59 yrs.: OR= 2.04 (95% CI: 1.10-3.78)  60-69 yrs.: OR= 3.88 (95% CI: 2.10-7.17)  70-79 yrs.: OR= 5.20 (95% CI: 2.61-10.34)  80+ yrs.: OR= 12.94 (95% CI: 6.14-27.26)  Model 3  18-39 yrs. Reference  40-49 yrs.: OR= 2.24 (95% CI: 1.13-4.43)  50-59 yrs.: OR= 2.62 (95% CI: 1.37-4.99)  60-69 yrs.: OR= 4.62 (95% CI: 2.39-9.95)  70-79 yrs.: OR= 5.68 (95% CI: 2.60-12.38)  80+ yrs.: OR= 19.08 (95% CI: 7.86-46.32) |
| Burn  2020  (177) | Autoimmune condition, chronic kidney disease, COPD, dementia, heart disease, hyperlipidemia, hypertension, malignant neoplasm, obesity, Type 2 diabetes, Charlson Index | Cox proportional hazards model using a non-linear relationship with age | Hospitalized with COVID-19 from general population: 8,582  Hospitalized from outpatient diagnosis: 9,437  From outpatient diagnosis:  20-24 yrs.  25-29 yrs.  30-34 yrs.  35-39 yrs.  40-44 yrs.  45-49 yrs.  50-54 yrs.  55-59 yrs.  60-64 yrs.  65-69 yrs.  70-74 yrs.  75-79 yrs.  80-84 yrs.  85-89 yrs.  90-94 yrs.  95+ yrs. | Hazard ratios (95% CIs)  Hospitalized with COVID-19 from general population  Age only model:  20-24 yrs.: 0.037 (0.032 to 0.042)  25-29 yrs.: 0.058 (0.052 to 0.064)  30-34 yrs.: 0.089 (0.082 to 0.096)  35-39 yrs.: 0.13 (0.13 to 0.14)  40-44 yrs.: 0.20 (0.19 to 0.21)  45-49 yrs.: 0.28 (0.27 to 0.29)  50-54 yrs.: 0.40 (0.39 to 0.41)  55-59 yrs.: 0.56 (0.55 to 0.56)  60-64 yrs.: 0.75 (0.75 to 0.76)  65-69 yrs.: Reference  70-74 yrs.: 1.30 (1.29 to 1.31)  75-79 yrs.: 1.65 (1.63 to 1.68)  80-84 yrs.: 2.06 (2.01 to 2.11)  85-89 yrs.: 2.51 (2.42 to 2.61)  90-94 yrs.: 3.00 (2.84 to 3.16)  95+ yrs.: 3.51 (3.27 to 3.76)  Hazard ratios (95% CIs)  Hospitalized from outpatient diagnosis  Age only model:  20-24 yrs.: 0.076 (0.067 to 0.086)  25-29 yrs.: 0.099 (0.091 to 0.11)  30-34 yrs.: 0.13 (0.12 to 0.14)  35-39 yrs.: 0.16 (0.15 to 0.17)  40-44 yrs.: 0.19 (0.18 to 0.20)  45-49 yrs.: 0.24 (0.22 to 0.25)  50-54 yrs.: 0.34 (0.32 to 0.36)  55-59 yrs.: 0.53 (0.51 to 0.55)  60-64 yrs.: 0.79 (0.78 to 0.80)  65-69 yrs.: Reference  70-74 yrs.: 1.10 (1.09 to 1.11)  75-79 yrs.: 1.07 (1.05 to 1.09)  80-84 yrs.: 0.97 (0.94 to 1.00)  85-89 yrs.: 0.83 (0.79 to 0.87)  90-94 yrs.: 0.70 (0.65 to 0.74)  95+ yrs.: 0.58 (0.54 to 0.64) | Hazard ratios (95% CIs)  Hospitalized with COVID-19 from general population  Age and comorbidity model:  20-24 yrs.: 0.063 (0.055 to 0.072)  25-29 yrs.: 0.094 (0.085 to 0.11)  30-34 yrs.: 0.14 (0.13 to 0.15)  35-39 yrs.: 0.20 (0.19 to 0.21)  40-44 yrs.: 0.28 (0.26 to 0.29)  45-49 yrs.: 0.38 (0.36 to 0.39)  50-54 yrs.: 0.50 (0.49 to 0.51)  55-59 yrs.: 0.64 (0.63 to 0.66)  60-64 yrs.: 0.81 (0.81 to 0.82)  65-69 yrs.: Reference  70-74 yrs.: 1.20 (1.19 to 1.21)  75-79 yrs.: 1.40 (1.38 to 1.43)  80-84 yrs.: 1.60 (1.55 to 1.65)  85-89 yrs.: 1.78 (1.70 to 1.87)  90-94 yrs.: 1.93 (1.81 to 2.06)  95+ yrs.: 2.05 (1.88 to 2.23)  Hazard ratios (95% CIs)  Hospitalized from outpatient diagnosis  Age and comorbidity model:  20-24 yrs.: 0.092 (0.08 to 0.10)  25-29 yrs.: 0.12 (0.11 to 0.13)  30-34 yrs.: 0.15 (0.14 to 0.16)  35-39 yrs.: 0.19 (0.18 to 0.20)  40-44 yrs.: 0.23 (0.21 to 0.24)  45-49 yrs.: 0.28 (0.26 to 0.30)  50-54 yrs.: 0.39 (0.36 to 0.41)  55-59 yrs.: 0.58 (0.55 to 0.60)  60-64 yrs.: 0.81 (0.80 to 0.83)  65-69 yrs.: Reference  70-74 yrs.: 1.09 (1.08 to 1.10)  75-79 yrs.: 1.07 (1.05 to 1.10)  80-84 yrs.: 0.99 (0.95 to 1.03)  85-89 yrs.: 0.87 (0.82 to 0.92)  90-94 yrs.: 0.75 (0.70 to 0.82)  95+ yrs.: 0.65 (0.59 to 0.72) |
| Carrillo-Vega†  2020  (180) | Sex, chronic kidney disease, COPD, diabetes & hypertension & obesity, diabetes & hypertension, diabetes & obesity, only hypertension, only obesity, only diabetes, pneumonia, health services | Logistic regression | N= 9946  Hospitalization:  3922 (39.4%)  25-49 yrs. 1532/5640  50-74 yrs. 2040/3823  ≥75 yrs. 350/483 | NA | Risk of hospitalization in COVID-19 positive cases OR (95% CI)  25-49 yrs.: Reference  50-74 yrs.: 2.05 (1.81-2.32)  ≥75 yrs.: 3.84 (2.90-5.10) |
| Cchiba and Patel  2020  (186) | Sex, asthma, race, smoking, obesity, hypertension, diabetes, obstructive sleep apnea, coronary artery disease, COPD, allergic rhinitis, rhinosinusitis, immunodeficiency | Poisson regression | N= 1526  Hospitalized: 853 (55.9%)  <40 yrs.: NA/414  40-69 yrs. NA/844  ≥70 yrs.: NA/268 | NA | Risk of hospitalization RR (95% CI)  <40 yrs.: 0.50 (0.38-0.64)  40-69 yrs. 0.76 (0.64-0.91)  ≥70 yrs.: Reference |
| Ebinger  2020  (201) | Sex, race, ethnicity, obesity, hypertension, diabetes mellitus, Elixhauser comorbidity score, prior myocardial infarction or heart failure, prior COPD or asthma, ACE inhibitor use, angiotensin receptor blocker use | Ordinal logistic regression | N= 442  Not Admitted:  228 (51.58%)  Admitted, Non-ICU:  137 (31%) | Population-based severe illness (Hospitalization) OR (95% CI)  *(Three interesting categories not together, difficult to compare for M.A.)*  Age (per 10 years):  1.68 (1.52-1.87) | Population-based severe illness (Hospitalization) OR (95% CI)  *(Three interesting categories not together, difficult to compare for M.A.)*  Age (per 10 years): 1.49 (1.30-1.70) |
| Giannouchos  2020  (210) | Gender, Mexican, smoker, chronical renal disease, diabetes, immunosuppression, COPD, obese, hypertension, cardiovascular disease, asthma, medical unit is a monitoring health unit for respiratory diseases (USMER), type of facility | Logistic regression analyses | N = 89,756  Hospitalizations:  31,271 (34.8%)  0-17 yrs.: 344/1885  18-44 yrs.: 7505/41,288  45-64 yrs.: 15,073/34,377  ≥65 yrs.: 8349/12,207 | NA | Hospitalization OR (95% CI)  0-17 yrs.: 1.73 (1.39-2.14)  18-44 yrs.: Reference  45-64 yrs.: 2.93 (2.76-3.12)  ≥65 yrs.: 7.24 (6.22-8.42) |
| Gu  2020  (215) | Sex, BMI, smoking status, alcohol consumption, race/ethnicity, neighborhood socioeconomic disadvantage index, population density, comorbidity score, respiratory disease, circulatory disease, any cancer, type 2 diabetes, kidney disease, liver disease, autoimmune disease | Logistic regression analyses, firth bias-corrected estimates | N= 765  Hospitalized: 270  <18 yrs.: 4/9  18- <35 yrs.: 38/203  35- <50 yrs.: 83/257  50- <65 yrs.: 171/349  65- <80 yrs.: 154/233  ≥80 yrs.: 73/88 | Hospitalization OR (95% CI)  Age (per 10 years):  1.69 (1.56-1.84)  Age (per year):  1.054 (1.045-1.063)  <18 yrs.: 3.52 (0.90-13.7)  18- <35 yrs.: Reference  35- <50 yrs.: 2.06 (1.33-3.19)  50- <65 yrs.: 4.13 (2.74-6.22)  65- <80 yrs.: 8.68 (5.56-13.6)  ≥80 yrs.: 25.3 (12.6-50.8) | Hospitalization OR (95% CI)  Age (per 10 years): 1.72 (1.53-1.93)  Age (per year): 1.056 (1.043-1.068)  <18 yrs.: 1.60 (0.21-12.4)  18- <35 yrs.: Reference  35- <50 yrs.: 1.70 (0.90-3.22)  50- <65 yrs.: 3.65 (2.01-6.61)  65- <80 yrs.: 6.61 (3.49-12.6)  ≥80 yrs.: 31.6 (12.7-78.5) |
| Killerby  2020  (230) | Sex, race, obesity, smoking, insurance status, hypertension, diabetes, chronic kidney disease, cardiovascular disease, chronic respiratory disease | Multivariable logistic regression | N=531  Hospitalized: 220 (41.4%)  18-44 yrs. 54/205  45-65 yrs. 76/196  ≥65 yrs. 90/130 | NA | Hospitalization OR 95% CIs  18-44 yrs.: Reference  45-65 yrs.: 1.0 (CIs not provided)  ≥65 yrs.: 3.4 (1.6-7.4) |
| Merzon  2020  (249) | Low vitamin D level, sex, SES, smoking, depression/anxiety, schizophrenia, dementia, diabetes, hypertension, cardiovascular disease, chronic lung disorders, BMI | Logistic regression | N= 782  Hospitalizations: NA  Age distribution: NA | Hospitalization OR (95% CI)  ≤50 years: Reference  >50 years: 2.51 (1.21–4.89) | Hospitalization OR (95% CI)  ≤50 years: Reference  >50 years: 2.71 (1.55–4.78) |
| Petrilli  2020  (263) | Week, race/ethnicity, smoking status, BMI, coronary artery disease, heart failure, hyperlipidemia, hypertension, diabetes, asthma or COPD, chronic kidney disease, cancer, sex | Mixed effects logistic regression models for admission to hospital | N= 5279  Hospitalization:  2741 (51.9%)  19-44 yrs.: 436/1846  45-54 yrs.: 410/902  55-64 yrs.: 605/1021  65-74 yrs.: 621/797  ≥75 yrs.: 668/713 | Hospital admission: OR (95% CIs)  Unadjusted  19-44 yrs.: Reference  45-54 yrs.: 2.69 (2.27-3.18)  55-64 yrs.: 4.69 (3.98-5.53)  65-74 yrs.: 11.38 (9.33-13.88)  ≥75 yrs.: 47.84 (34.73-65.91) | Hospital admission: OR (95% CIs)  Adjusted  19-44 yrs.: Reference  45-54 yrs.: 2.14 (1.76-2.59)  55-64 yrs.: 3.67 (3.01-4.48)  65-74 yrs.: 8.7 (6.77-11.22)  ≥75 yrs.: 37.87 (26.1-56.03) |
| Price-Haywood  2020  (266) | Race, sex, Charlson Comorbidity Index score, low-income residency, insurance, obesity | Hospitalization: logistic regression | N=3481  Admitted to hospital:  1382 (39.7%) | Hospitalization: OR (95% CI)  Adjusted for race and sex:  Per 5-yr. unit increase:  1.34 (1.30-1.37)  Per yr. increase: 1.06(1.05-1.06) | Hospitalization: OR (95% CI)  Fully adjusted:  Per 5-yr. unit increase: 1.29 (1.25-1.33)  Per yr. increase: 1.052(1.045-1.059) |
| Reilev  2020  (269) | Number of comorbidities (0,1,2,3,4+ of chronic lung disease, hypertension, ischaemic heart disease, heart failure, atrial fibrillation, stroke, diabetes, dementia, cancer, chronic liver disease, hospital-diagnosed kidney disease, alcohol abuse, substance abuse, major psychiatric disorders, organ transplantation, medical overweight and obesity, rheumatoid arthritis/connective-tissue disease), sex | Logistic regression | N= 11,122  Hospitalization:  2254 (20%)  0-9 yrs.: 11/251  10-19 yrs.: 14/495  20-29 yrs.: 55/1523  30-39 yrs.: 90/1588  40-49 yrs.: 192/1970  50-59 yrs.: 337/2035  60-69 yrs.: 374/1305  70-79 yrs.: 578/950  80-89 yrs.: 476/762  90+ yrs.: 127/243 | Hospitalization among positive cases OR (95% CI)  Unadjusted  0-9 yrs.: 0.2 (0.1-0.4)  10-19 yrs.: 0.1 (0.1-0.3)  20-29 yrs.: 0.2 (0.1-0.3)  30-39 yrs.: 0.3 (0.2-0.4)  40-49 yrs.: 0.5 (0.5-0.7)  50-59 yrs.: Reference  60-69 yrs.: 2.0 (1.7-2.4)  70-79 yrs.: 7.8 (6.6-9.3)  80-89 yrs.: 8.4 (7.0-10.1)  90+ yrs.: 5.5 (4.2-7.3) | Hospitalization among positive cases OR  (95% CI)  Adjusted  0-9 yrs.: 0.3 (0.2-0.6)  10-19 yrs.: 0.2 (0.1-0.3)  20-29 yrs.: 0.2 (0.2-0.3)  30-39 yrs.: 0.4 (0.3-0.5)  40-49 yrs.: 0.6 (0.5-0.8)  50-59 yrs.: Reference  60-69 yrs.: 1.6 (1.3-1.9)  70-79 yrs.: 4.7 (3.9-5.7)  80-89 yrs.: 4.8 (3.9-5.8)  90+ yrs.: 3.5 (2.6-4.7) |
| Rentsch  2020  (270) | Adjusted model 1:  Race, baseline comorbidities (chronic kidney disease, COPD, diabetes, hypertension, vascular disease), Medication history in year prior to test date (angiotensin converting enzyme inhibitor or angiotensin II receptor blocker, nonsteroidal anti-inflammatory drug), vital signs (systolic blood pressure, oxygen saturation, pulse, temperature), laboratory findings (albumin, eGFR, FIB-4, Hemoglobin, White blood cell count, Lymphocyte count)  Adjusted model 2:  Race, baseline comorbidities (chronic kidney disease, COPD, diabetes, hypertension, vascular disease), Medication history in year prior to test date (angiotensin converting enzyme inhibitor or angiotensin II receptor blocker, nonsteroidal anti-inflammatory drug), vital signs (systolic blood pressure, oxygen saturation, pulse, temperature), VACS Index score | Logistic regression | N= 585  Hospitalization: 297 (50.8%)  54-59 yrs.: 58/135  60-64 yrs.: 62/135  65-69 yrs.: 62/120  70-75 yrs.: 115/195 | Hospitalization: OR (95% CI)  Per 5-yr. increase: 1.26 (1.10-1.44)  Per yr. increase: 1.04 (1.02-1.08) | Hospitalization: OR (95% CI)  Model 1:  Per 5-yr. increase: 0.87 (0.71-1.05)  Per yr. increase: 0.97 (0.93-1.01)  Model 2:  Per 5-yr. increase: 0.64 (0.51-0.80)  Per yr. increase: 0.91 (0.87-0.96) |
| Rossi  2020  (274) | Sex, calendar period, time from symptoms to diagnosis, place of birth, Charlson Comorbidity Index | Cox proportional hazard regression models | N=2653  Hospitalized: 1075 (40.5%)  <51 yrs.: 107/696  51-60 yrs.: 128/528  61-70 yrs.: 205/413  71-80 yrs.: 291/420  ≥81 yrs.: 344/596 | NA | Hospitalization HR (95% CIs)  <51 yrs.: Reference  51-60 yrs.: 1.3 (1.0-1.8)  71-70 yrs.: 3.2 (2.4-4.1)  71-80 yrs.: 5.9 (4.5-7.6)  ≥81 yrs.: 7.1 (5.4-9.3) |
| Soares  2020  (293) | Sex, race, cardiovascular disease, diabetes, kidney disease, obesity, pulmonary disease, smoking, fever, headache, runny nose, shortness of breath, sore throat | Logistic regression | N=10,713  Hospitalized: 1152 (10.8%)  <60 yrs.: 546/8676  ≥60 yrs.: 606/2037 | Hospitalization due to COVID-19  OR (95% CIs)  <60 yrs.: Reference  ≥60 yrs.: 6.31 (5.55-7.17) | Hospitalization due to COVID-19  OR (95% CIs)  <60 yrs.: Reference  ≥60 yrs.: 3.40 (2.91-3.96) |
| van Gerwen  2020  (301) | Sex, race, smoking, BMI, hypertension, coronary artery disease, atrial fibrillation, congestive heart failure, peripheral vascular disease, cerebrovascular accident/transient ischemic attack, dementia, diabetes, hypothyroidism, chronic kidney disease, malignancy, asthma, COPD, prior venous thromboembolism | Logistic regression | N= 3703  Hospitalized: 2015 (54.4%)  18-40 yrs.: 207/861  40-60 yrs.: 539/1173  >60 yrs.: 1269/1669 | Hospitalization OR (95% CI)  18-40 yrs.: Reference  40-60 yrs.: 2.69 (2.21‐3.26)  >60 yrs.: 10.02 (8.27‐12.15) | Hospitalization OR (95% CI)  18-40 yrs.: Reference  40-60 yrs.: 2.02 (1.62‐2.50)  >60 yrs.: 5.47 (4.29‐6.96) |
| ICU: intensive care unit; COPD: chronic obstructive pulmonary disease; BMI: body mass index; yrs.: years; SOFA score: sequential organ failure assessment score; OR: odds ratio; HR: hazards ratio; CI: confidence intervals; NA: not available | | | | | |

**Table S8. Risk of bias summary for studies investigating hospitalization**

| **Study** | **Major domains** | | | | | **Minor domains** | | | **OVERALL** |
| --- | --- | --- | --- | --- | --- | --- | --- | --- | --- |
|  | **Recruitment procedure** | **Exposure assessment** | **Outcome source and validation** | **Confounding** | **Analysis method** | **Chronology** | **Funding** | **Conflict of interest** |  |
| Azar et al. 2020  (167) |  |  |  |  |  |  |  |  |  |
| Burn et al. 2020  (177) |  |  |  |  |  |  |  |  |  |
| Carrillo-Vega et al. 2020  (180) |  |  |  |  |  |  |  |  |  |
| Cchiba and Patel 2020  (186) |  |  |  |  |  |  |  |  |  |
| Ebinger et al. 2020  (201) |  |  |  |  |  |  |  |  |  |
| Giannouchos et al. 2020  (210) |  |  |  |  |  |  |  |  |  |
| Gu et al. 2020  (215) |  |  |  |  |  |  |  |  |  |
| Killerby et al. 2020  (230) |  |  |  |  |  |  |  |  |  |
| Merzon et al. 2020  (249) |  |  |  |  |  |  |  |  |  |
| Petrilli et al. 2020  (263) |  |  |  |  |  |  |  |  |  |
| Price-Haywood et al. 2020  (266) |  |  |  |  |  |  |  |  |  |
| Reiley et al. 2020  (269) |  |  |  |  |  |  |  |  |  |
| Rentsch et al. 2020  (270) |  |  |  |  |  |  |  |  |  |
| Rossi et al. 2020  (274) |  |  |  |  |  |  |  |  |  |
| Soares et al. 2020  (293) |  |  |  |  |  |  |  |  |  |
| van Gerwen et al. 2020  (301) |  |  |  |  |  |  |  |  |  |
| = low risk of bias = unclear risk of bias = high risk of bias | | | | | | | | | |

**Figure S8. Relationship between median age and log of relative risk among studies using age as a categorical value (linear and cubic models, variance weighted): hospitalization**

**Figure S9. Funnel plot for studies investigating risk of hospitalization**

**Table S9. Results of studies investigating admission to ICU**

| **Author,**  **Year**  **(Ref)** | **Confounders and**  **age-dependent risk factors used in the model** | **Type of analysis** | **N in analysis**  **Total ICU admissions**  **In age group: # cases/total** | **Results**  **Unadjusted effect estimates (if available)** | **Results**  **Adjusted effect estimates** |
| --- | --- | --- | --- | --- | --- |
| Argenziano  2020  (166) | Sex, BMI, smoking, coronary artery disease, congestive heart failure, history of stroke, diabetes mellitus, hypertension, cirrhosis, HIV, inflammatory bowel disease, pulmonary disease, renal disease, viral hepatitis, active malignancy, transplant history, rheumatological disease, immunosuppressed state, no comorbidities | Cox proportional hazards analysis only with complete data | N= 841  ICU patients:  236 (24%) | NA | ICU HR (95% CI)  Age (per year): 0.996 (0.985-1.01) |
| Bi, Hong, and Meng  2020  (173) | Sex, hypertension, diabetes, coronary heart disease, chronic lung disease, cerebrovascular disease, total comorbidity, fever, cough, shortness of breath, muscle soreness, fatigue, PaO2/FiO2, lymphocyte count, platelet count, C-reactive protein, D-dimer | Competing risk regressions | N= 420  ICU admissions: 19  <39yrs.: 1/163  40-59 yrs.: 4/149  60+ yrs.: 14/108 | NA | Risk of ICU admission HR (95% CI)  0-39 yrs.: Reference  40-59 yrs.: 4.4 (0.5-39.6)  60+ yrs.: 22.4 (3.0-168.2) |
| Du  2020  (199) | Number of comorbidities (0, 1, ≥2), highest temperature, lymphocyte Count, detection of SARS-CoV-2  Comorbidities considered in number of comorbidities: hypertension, diabetes, cardiovascular/cerebrovascular disease, chronic digestive disease, pre-existing pulmonary tuberculosis, chronic liver/kidney disease, chronic obstructive lung disease, peripheral artery disease, malignancy) | Ordinal logit regression | N= 179  Mild-to-moderate: 79 (44.2%)  Severe: 57 (31.8%)  Critically ill (ICU admission): 43 (24%) | NA | Disease severity (*calculated ORs don’t allow us to compare to other studies on ICU admission*)  OR (95% CI)  <60 yrs.: Reference  60-69 yrs.: 1.58 (1.06-2.37)  ≥70 yrs.: 1.69 (1.06-2.68) |
| Gu  2020  (215) | Sex, BMI, smoking status, alcohol consumption, race/ethnicity, neighborhood socioeconomic disadvantage index, population density, comorbidity score, respiratory disease, circulatory disease, any cancer, type 2 diabetes, kidney disease, liver disease, autoimmune disease | Logistic regression analyses, firth bias-corrected estimates | N= 756  ICU: 141  <18 yrs.: 3/9  18- <35 yrs.: 26/203  35- <50 yrs.: 36/257  50- <65 yrs.: 96/346  65- <80 yrs.: 91/233  ≥80 yrs.: 31/88 | ICU OR (95% CI)  Age (per 10 years): 1.37 (1.26-1.49)  Per yr. increase (own calculations):  1.03 (1.02-1.04)  <18 yrs.: 3.61 (0.86-15.1)  18- <35 yrs.: Reference  35- <50 yrs.: 1.10 (0.64-1.89)  50- <65 yrs.: 2.55 (1.59-4.09)  65- <80 yrs.: 4.39 (2.7-7.15)  ≥80 yrs.: 3.87 (2.12-7.07) | Population-based ICU OR (95% CI)  Age (per 10 years): 1.45 (1.27-1.65)  Per yr. increase (own calculations)  1.04 (1.02-1.05)  <18 yrs.: 3.67 (0.45-30)  18- <35 yrs.: Reference  35- <50 yrs.: 1.33 (0.57-3.11)  50- <65 yrs.: 2.96 (1.39-6.33)  65- <80 yrs.: 4.44 (2.01-9.82)  ≥80 yrs.: 7.70 (3.10-19.1) |
| Hashemi  2020  (220) | Chronic liver disease, obesity, sex, cardiac diseases, hypertension, diabetes, hyperlipidaemia, pulmonary disorders | Logistic regression | N= 363  ICU: NA | NA | In-hospital ICU admission  OR (95% CI)  Age (per year): 1.01 (0.99-1.02) |
| Kalligeros  2020  (228) | Ethnicity/race, sex, BMI, diabetes, hypertension, heart disease, lung disease | Logistic regression | N= 103  ICU admission: 41 (39.8%) | In-hospital ICU-admission  OR (95% CI)  Age (per year): 1.02 (1.00-1.05) | In-hospital ICU-admission  OR (95% CI)  Age (per year): 1.03 (1.00-1.07) |
| Kim  2020  (231) | Sex, race, ethnicity, smoker, hypertension, obesity, diabetes, chronic lung disease, cardiovascular disease, neurologic, renal, immunosuppression, gastrointestinal or liver, hematologic, rheumatologic or autoimmune, outpatient ACE-inhibitor use, angiotensin receptor blocker use prior to hospitalization | Log-linked Poisson generalized estimating equation regressions with an exchangeable correlation matrix | N=2490  ICU admissions:  798 (32.0%)  18-39 yrs.: 8/302  40-49 yrs.: 91/318  50-64 yrs.: 32/744  65-74 yrs.: 23/478  75-84 yrs.: 19/397  ≥85 yrs.: 8/251 | ICU RR (95% CI)  18-39 yrs.: Reference  40-49 yrs.: 1.38 (1.08-1.75)  50-64 yrs.: 1.64 (1.36-1.99)  65-74 yrs.: 1.80 (1.50-2.16)  75-84 yrs.: 1.80 (1.52-2.14)  ≥85 yrs.: 1.16 (0.87-1.54) | ICU RR (95% CI)  18-39 yrs.: Reference  40-49 yrs.: 1.22 (0.96-1.56)  50-64 yrs.: 1.53 (1.28-1.83)  65-74 yrs.: 1.65 (1.34-2.03)  75-84 yrs.: 1.84 (1.60-2.11)  ≥85 yrs.: 1.43 (1.00-2.04) |
| Price-Haywood  2020  (266) | Race, sex, Charlson Index, obesity | In-hospital ICU: Cox proportional-hazards models  For variables for which less than 25% of data missing, values were imputed | N= 1382  ICU:  474 (34.3%) | ICU: OR (95% CI)  Adjusted for race and sex:  Per 5-yr. unit increase:  0.99 (0.96-1.02) | ICU: OR (95% CI)  Fully adjusted:  Per 5-yr. unit increase: 1.01 (0.98-1.05)  Per yr.- unit increase (own calculations):  1.002 (0.996-1.010) |
| Rentsch  2020  (270) | Adjusted model 1:  Race, baseline comorbidities (chronic kidney disease, COPD, diabetes, hypertension, vascular disease), Medication history in year prior to test date (angiotensin converting enzyme inhibitor or angiotensin II receptor blocker, nonsteroidal anti-inflammatory drug), vital signs (systolic blood pressure, oxygen saturation, pulse, temperature), laboratory findings (albumin, eGFR, FIB-4, Hemoglobin, White blood cell count, Lymphocyte count)  Adjusted model 2:  Race, baseline comorbidities (chronic kidney disease, COPD, diabetes, hypertension, vascular disease), Medication history in year prior to test date (angiotensin converting enzyme inhibitor or angiotensin II receptor blocker, nonsteroidal anti-inflammatory drug), vital signs (systolic blood pressure, oxygen saturation, pulse, temperature), VACS Index score | Logistic regression | N= 585  ICU: 122 (20,9%)  54-59 yrs.: 11/135  60-64 yrs.: 24/135  65-69 yrs.: 28/120  70-75 yrs.: 59/195 | Hospitalization: OR (95% CI)  Per 5-yr. increase: 1.55 (1.30-1.86)  Per yr. increase: 1.09 (1.05-1.13) | Hospitalization: OR (95% CI)  Model 1:  Per 5-yr. increase: 1.31 (1.03-1.66)  Per yr. increase: 1.06 (1.01-1.11)  Model 2:  Per 5-yr. increase: 0.98 (0.76-1.26)  Per yr. increase: 1.0 (0.95-1.05) |
| Seiglie  2020  USA  (280) | BMI, sex, race/ethnicity, diabetes, coronary artery disease or myocardial infarction, chronic heart failure, hypertension, COPD/asthma, active cancer, liver disease, renal disease | Logistic regression | N = 436  ICU: 156 (34.7%)  <50 yrs.: NA/104  50-59 yrs.: NA/81  60-69 yrs.: NA/82  ≥70 yrs.: NA /181 | 14-day ICU admission OR (95% CIs)  <50 yrs.: Reference  50-59 yrs.: 1.37 (0.75-2.49)  60-69 yrs.: 1.05 (0.57-1.93)  ≥70 yrs.: 0.87 (0.89-1.96) | 14-day ICU admission OR (95% CIs)  <50 yrs.: Reference  50-59 yrs.: 1.13 (0.57-2.24)  60-69 yrs. 1.22 (0.60-2.49)  ≥70 yrs.: 1.41 (1.20-3.88) |
| Suleyman  2020  (296) | For ICU:  Sex, race, severe obesity, chronic kidney disease, cancer, diabetes, hypertension, coronary artery disease | Logistic regression | N= 463  ICU: 141 (39.7%)  ≤60 yrs.: 49/245  >60 yrs.: 92/218 | NA | ICU OR (95% CI)  ≤60 yrs.: Reference  >60 yrs.: 1.6 (1.0Bi-2.7) |
| Tai  2020  (299) | Model 2:  Cardiovascular conditions, sex, chest tightness, diabetes mellitus, lung diseases | Logistic regression | N= 332  ICU: 58 (17.5%) | ICU OR (95% CI)  Age (per year): 1.006 (0.983-1.030) | ICU OR (95% CI)  Model 2  Age (per year): 0.993 (0.966-1.020) |
| ICU: intensive care unit; COPD: chronic obstructive pulmonary disease; BMI: body mass index; yrs.: years; SOFA score: sequential organ failure assessment score; OR: odds ratio; HR: hazards ratio; CI: confidence intervals; NA: not available | | | | | |

**Table S10. Risk of bias summary for studies investigating risk of admission to ICU for hospitalized patients**

| **Study** | **Major domains** | | | | | **Minor domains** | | | **OVERALL** |
| --- | --- | --- | --- | --- | --- | --- | --- | --- | --- |
|  | **Recruitment procedure** | **Exposure assessment** | **Outcome source and validation** | **Confounding** | **Analysis method** | **Chronology** | **Funding** | **Conflict of interest** |  |
| Argenziano et al. 2020  (166) |  |  |  |  |  |  |  |  |  |
| Bi, Hong, and Meng et al. 2020  (173) |  |  |  |  |  |  |  |  |  |
| Du et al. 2020  (199) |  |  |  |  |  |  |  |  |  |
| Gu et al. 2020*  (215) |  |  |  |  |  |  |  |  |  |
| Hashemi et al. 2020  (220) |  |  |  |  |  |  |  |  |  |
| Kalligeros et al. 2020  (228) |  |  |  |  |  |  |  |  |  |
| Kim et al. 2020  (231) |  |  |  |  |  |  |  |  |  |
| Price-Haywood et al. 2020  (266) |  |  |  |  |  |  |  |  |  |
| Rentsch et al. 2020  (270) |  |  |  |  |  |  |  |  |  |
| Sieglie et al. 2020  (280) |  |  |  |  |  |  |  |  |  |
| Suleyman et al. 2020  (296) |  |  |  |  |  |  |  |  |  |
| Tai et al. 2020  (299) |  |  |  |  |  |  |  |  |  |
| *Positive test-based ICU = low risk of bias = unclear risk of bias = high risk of bias | | | | | | | | | |

**Table S11. Results of studies investigating risk of mechanical ventilation**

| **Author,**  **Year**  **(Ref)** | **Confounders and**  **age-dependent risk factors used in the model** | **Type of analysis**  **# cases / # non-cases** | **N in analysis**  **Total mechanical ventilation**  **In age group: # cases/total** | **Results**  **Unadjusted effect estimates (if available)** | **Results**  **Adjusted effect estimates** |
| --- | --- | --- | --- | --- | --- |
| Costa Monteiro  2020  (191) | Sex, race, obesity, diabetes, hypertension, coronary artery disease, chronic kidney disease, smoking | Multivariable logistic regression  Past medical history with cohort prevalence of >15% used in the model | N=112  Mechanical ventilation: 28 (25%) | NA | Mechanical ventilation OR (95% CI)  Per year: 0.99 (0.96-1.03) |
| Hashemi  2020  (220) | Chronic liver disease, obesity, sex, cardiac diseases, hypertension, diabetes, hyperlipidaemia, pulmonary disorders | Logistic regression | N=363  Mechanical ventilation: NA | NA | In-hospital mechanical ventilation OR (95% CI)  Age (per year): 1.01 (0.99-1.03) |
| Kalligeros  2020  (228) | Ethnicity/race, sex, BMI, diabetes, hypertension, heart disease, lung disease | Logistic regression | N= 103  Mechanical ventilation: 29 (28.2%) | Mechanical ventilation OR (95% CI)  Age (per year): 1.01 (0.98-1.04) | Mechanical ventilation OR (95% CI)  Age (per year): 1.02 (0.98-1.06) |
| Patel‡  2020  (261) | Therapeutic anticoagulation, home antiplatelet, race, BMI, Charlson score, glucose level | Poisson regression model and logistic regression | N= 1716  Mechanical ventilation= 254 (14.8%)  18-44 yrs.: NA/379  45- 59 yrs.: NA/508  60-69 yrs.: NA/371  70-79 yrs.: NA/242  >80 yrs.: NA/216 | NA | Mechanical ventilation  18-44 yrs.: OR= 1.16 (95% CI: 0.71-1.92)  45- 59 yrs.: Reference  60-69 yrs.: OR= 1.24 (95% CI: 0.79-1.94)  70-79 yrs.: OR= 0.89 (95% CI: 0.51-1.54)  >80 yrs.: OR= 0.42 (95% CI: 0.21-0.84) |
| Seiglie  2020  (280) | BMI, sex, race/ethnicity, diabetes, coronary artery disease or myocardial infarction, chronic heart failure, hypertension, COPD/asthma, active cancer, liver disease, renal disease | Logistic regression | N= 436  Intubations: 129 (29.7%)  <50 yrs.: NA/104  50-59 yrs.: NA/82  60-69 yrs.: NA/82  ≥70 yrs.: 181 | 14-day mechanical ventilation  OR (95% CIs)  <50 yrs.: Reference  50-59 yrs.: 1.11 (0.59-2.09)  60-69 yrs.: 0.99 (0.53-1.87)  ≥70 yrs.: 0.85 (0.50-1.45) | 14-day mechanical ventilation  OR (95% CIs)  <50 yrs.: Reference  50-59 yrs.: 1.02 (0.50-2.08)  60-69 yrs.: 1.20 (0.57-2.54)  ≥70 yrs.: 1.72 (0.82-3.61) |
| Suleyman  2020  (296) | For mechanical ventilation:  Sex, race, severe obesity, chronic kidney disease, cancer, diabetes, hypertension, coronary artery disease, congestive heart failure, tobacco use | Logistic regression | N= 355  Mechanical ventilation: 114 (32.1%)  ≤60 yrs.: NA/153  >60 yrs.: NA/202 | NA | Mechanical ventilation OR (95% CI)  ≤60 yrs.: Reference  >60 yrs.: 3.5 (1.9-6.4) |
| van Gerwen  2020  (301) | Sex, race, smoking, BMI, hypertension, coronary artery disease, atrial fibrillation, congestive heart failure, peripheral vascular disease, cerebrovascular accident/transient ischemic attack, dementia, diabetes, hypothyroidism, chronic kidney disease, malignancy, asthma, COPD, prior venous thromboembolism | Logistic regression | N=2015  Mechanical ventilation: 525 (26% of hospitalized)  18-40 yrs.: 29/207  40-60 yrs.: 135/539  >60 yrs.: 361/1269 | Mechanical ventilation OR (95% CI)  18-40 yrs.: Reference  40-60 yrs.: 2.05 (1.32‐3.18)  >60 yrs.: 2.44 (1.62-3.68) | Mechanical ventilation OR (95% CI)  18-40 yrs.: Reference  40-60 yrs.: 2.12 (1.35‐3.32)  >60 yrs.: 3.26 (2.08‐5.11) |
| ICU: intensive care unit; COPD: chronic obstructive pulmonary disease; BMI: body mass index; yrs.: years; SOFA score: sequential organ failure assessment score; OR: odds ratio; HR: hazards ratio; CI: confidence intervals; NA: not availableICU: intensive care unit; COPD: chronic obstructive pulmonary disease; BMI: body mass index; yrs.: years; SOFA score: sequential organ failure assessment score; OR: odds ratio; HR: hazards ratio; CI: confidence intervals; NA: not available | | | | | |

**Table S12. Risk of bias summary for studies investigating risk of mechanical ventilation**

| **Study** | **Major domains** | | | | | **Minor domains** | | | **OVERALL** |
| --- | --- | --- | --- | --- | --- | --- | --- | --- | --- |
|  | **Recruitment procedure** | **Exposure assessment** | **Outcome source and validation** | **Confounding** | **Analysis method** | **Chronology** | **Funding** | **Conflict of interest** |  |
| Costa Monteiro et al. 2020  (191) |  |  |  |  |  |  |  |  |  |
| Hashemi et al. 2020  (220) |  |  |  |  |  |  |  |  |  |
| Kalligeros et al. 2020  (228) |  |  |  |  |  |  |  |  |  |
| Patel et al. 2020  (261) |  |  |  |  |  |  |  |  |  |
| Seiglie et al. 2020  (280) |  |  |  |  |  |  |  |  |  |
| Suleyman et al. 2020  (296) |  |  |  |  |  |  |  |  |  |
| van Gerwen et al. 2020  (301) |  |  |  |  |  |  |  |  |  |
